# Establishing consensus on domains and items for Reporting intervention Fidelity in Non-Drug, non-surgical trials: the ReFiND Delphi study

**DOI:** 10.64898/2026.09.14.26362897

**Authors:** Fernando Sousa, Melanie K. Farlie, Terry Haines, Belinda Borrelli, Christopher Carroll, Catherine Mathews, Daniel C. Ribeiro, Julie M. Fritz, Martin Underwood, Nadine E. Foster, Sarah E. Lamb, Zila M. Sanchez, Peter Malliaras

## Abstract

**Objectives:** To develop consensus on core domains and items to inform a reporting guideline for intervention fidelity in non-drug, non-surgical trials (ReFiND).

**Design:** International, online three-round Delphi process.

**Methods:** We invited researchers with expertise in intervention fidelity, non-drug, non-surgical trials, trial reporting, and process evaluation methods to take part in a three-round Delphi study. They rated the importance of 7 intervention fidelity domains and 79 items using a 9-point scale (1 to 3= ‘not important’, 4 to 6= ‘important but not essential’, 7 to 9= ‘very important’). A comment box was provided along with each domain and item for panellists to provide the rationale for their ratings. Consensus was defined as more than 70% of panellists rating 1-3 (exclusion) or 7-9 (inclusion) after two voting rounds. A summary of ratings and panellists’ rationale from Rounds One and Two were provided to panellists in Round Two and Three, respectively. We conducted a thematic analysis to synthesise qualitative data.

**Results:** Sixty-one panellists completed Round One, 55 completed Round Two and 56 completed Round Three. Panellists had diverse backgrounds and were from 21 countries, including six low- and middle-income countries. After three rounds, five domains (Intervention Design, Intervention Fidelity Measurement Methods, Provider Training, Intervention Delivery by Providers, Intervention Receipt by Participants) and 20 items reached consensus for inclusion in the reporting guideline. Panellists’ rationale for their ratings was based on how specifically each domain and item related to intervention fidelity, how feasible it would be to implement in practice, and how broadly it could be applied across different contexts.

**Conclusions:** Using systematic methods, this study reached consensus on a core set of domains and items for inclusion in ReFiND. This core set provides a foundation for consistent and transparent reporting of intervention fidelity and helps researchers and clinicians better interpret and replicate interventions tested in non-drug, non-surgical trials.

**Key messages:** *What is already known on this topic:*

- Intervention fidelity is poorly reported across non-drug, non-surgical trials
- There is a lack of standardisation and conflicting information in intervention fidelity domains and reporting recommendations, making adequate reporting difficult

*What this study adds:*

- Using systematic methods, this study reached consensus on five domains and 20 items considered essential for fidelity reporting in non-drug, non-surgical trials

*How this study might affect research, practice or policy:*

- The findings of this study will inform a reporting guideline on intervention fidelity in non-drug, non-surgical trials (ReFiND), supporting greater transparency, replication, and reproducibility of such interventions.

## Introduction

Intervention fidelity, which refers to the extent to which an intervention is implemented as intended, can influence both the effectiveness of an intervention and its replication by researchers and implementation in practice.^1^ ^2^ Reporting intervention fidelity in a trial allows researchers and clinicians to assess how closely the delivered components aligned with what was intended, facilitating the interpretation of outcomes and the real-world implementation of the intervention.

A recent meta-epidemiological study found that intervention effects may be overestimated in randomised controlled trials where intervention fidelity is poor or not reported.^3^ Despite the importance of adequate reporting, intervention fidelity is often poorly and inconsistently reported in trials of non-drug, non-surgical interventions.^4–9^ This issue has not improved over time,^10^ even with research efforts aimed at addressing intervention fidelity. Several guidance documents providing recommendations on intervention fidelity exist;^1^ ^2^ ^9^ ^11–13^ however, they were not developed using systematic, consensus-based methods, and agreement across recommendations is limited. Reporting guidelines using consensus methods, such as TIDieR (Template for Intervention Description and Replication)^14^ and CONSORT (Consolidated Standards of Reporting Trials),^15^ include a few fidelity-related recommendations. However, because these are general guidelines and not specifically focused on intervention fidelity, many fidelity aspects discussed in the literature were not included in their Delphi processes.

Our scoping review of 73 guidance documents on intervention fidelity identified seven fidelity domains (i.e., a connected set of aspects related to fidelity), but only two were reported in more than half of the guidance documents.^12^ Across these domains, 81 fidelity recommendations (i.e., individual, actionable items) were identified in the review, yet only 14 appeared in more than half of the documents.^12^ Such a high volume of heterogeneous recommendations are not feasible to report in trials. Together, these findings, which reflect current evidence, highlight the timely need to reach consensus on an overarching core set of reporting guidelines for intervention fidelity.^16–18^

This Delphi study was conducted to identify the key domains and items essential for reporting intervention fidelity. The findings will inform the development of an international Reporting guideline for intervention Fidelity in Non-Drug, non-surgical trials (ReFiND),^19^ promoting greater transparency, interpretability, and reproducibility of evidence.

## Methods

### Registration

We registered our intention to develop ReFiND with the EQUATOR Network (July 2023)^20^ and published our protocol (July 2024) elsewhere.^19^ This study received approval from the Monash University Human Research Ethics Committee (ID 41579) and was conducted according to the EQUATOR Guidance for Developers of Health Research Reporting.^21^

The ReFiND project includes six stages of development: (1) formation of a steering committee, (2) developing a scoping review, (3) conducting a Delphi study, (4) running consensus meetings, (5) elaborating a guideline statement, and (6) implementing dissemination strategies.^19^ This paper reports on the Delphi study and the earlier stages that informed it. We used ACCORD (ACcurate COnsensus Reporting Document) to guide the reporting of this paper.^22^

### Steering Committee

We formed a guideline development group (GDG), which served as the steering committee for the project, including 13 researchers from various backgrounds, expertise, disciplines, and geographical locations. The process of selecting GDG members was described in our protocol.^19^

### Panellists

The eligibility criteria for the Delphi panel included having at least one publication related to intervention fidelity, trials of non-drug, non-surgical interventions, process evaluation, or trial reporting methods. We identified potential panellists from a list of authors of guidance documents on intervention fidelity included in a scoping review conducted by our GDG.^12^ Given the limited diversity (e.g., geographic location) of the documents included in the scoping review,^12^ we conducted searches on PubMed to identify authors of recent publications relevant to the eligibility criteria, aiming to better balance geographic representation. We aimed to invite a broad range of potential panellists to ensure diversity and relevant expertise. GDG members could participate as panellists.

As we had already identified a large number of potential eligible researchers from the scoping review and sought a targeted approach to balance geographic representation, we invited researchers directly to participate via email rather than advertising the study in professional and research networks, as outlined in our protocol.^19^ In the email, we explained the purpose and processes of the study, along with their expected contributions. We also stated that participating in the Delphi rounds and making significant contributions to the manuscript, in line with ICMJE (International Committee of Medical Journal Editors) recommendations, would result in an invitation to co-author the guideline statement.

### Preparatory research

We conducted a scoping review of existing guidance on intervention fidelity to identify domains and items for inclusion in the Delphi process.^12^ The review identified seven domains and 81 fidelity recommendations across 73 guidance documents. Two researchers (FS and PM) reviewed these domains and recommendation items to identify potential overlaps and improve clarity. They suggested merging certain items, splitting others, and rewording some domains and items, which were then reviewed by the GDG. This refinement process resulted in seven domains and 79 items for inclusion in the first Delphi survey. Each domain was accompanied by a brief description and examples. We stated in the survey that the domains and items were extracted from a scoping review. Initially, we had planned to put these domains and items in a survey for the GDG members to vote on which should be included in the first Delphi round. However, through internal discussions within the GDG, we opted to include all items in the Delphi to ensure input from the broad range of panellists.

Informed by the scoping review findings, the GDG developed a study-specific working definition of fidelity for use in the Delphi study, which was applied to support clarity and consistency across Delphi rounds. First, GDG members selected their preferred definition from a list of ten definitions extracted from the scoping review and provided comments on all of them. Second, based on these selections and the feedback received, the project leadership team drafted two potential working definitions. Third, these were then discussed among GDG members through online meetings and email correspondence over multiple iterations until consensus was reached. Finally, the group agreed on a working definition that was broad and not tied to specific fidelity domains: *‘Intervention fidelity refers to the extent to which an intervention is implemented as planned in the trial protocol’*.

### Delphi rounds

There were three Delphi rounds. All online surveys were distributed via Qualtrics © (Provo, UT), a survey management platform. We sent an email from an institutional Monash University account to panellists to reassure them that the invitation to participate sent via Qualtrics was legitimate and secure. Identifiable information (name and email address for follow-up in subsequent Delphi rounds) was collected in the Round One survey but stored separately from panellists’ survey responses. A unique ID, randomly generated by Qualtrics, was used to link responses across rounds while maintaining anonymity.

Each round included key instructions, a section with intervention fidelity domains, and another with the individual items. Given that the surveys were lengthy, items were presented to participants in random order using Qualtrics’ randomizer feature to minimise fatigue effects. The first Delphi round included the study aims, reasons for the invitation, expected contributions, details on the opportunity for co-authorship, an explanatory statement for consent, demographic data collection, and a set of open-ended questions where panellists could make suggestions related to domains and items (Appendix 1). Each survey was piloted by one health professional who delivers non-drug, non-surgical interventions and two researchers. We made slight changes (e.g., wording and formatting) based on their feedback. Given the length of the surveys, we did not include a specific question about applicability to different types of trials for each item, as originally stated in our protocol.^19^ Instead, we instructed panellists that items considered very important for inclusion should be applicable to nearly all types of trials (Appendix 1-3).

Rounds remained open for 3-4 weeks, with one email reminder sent each week. Round One was initially anticipated to start in January 2024, but because the scoping review to inform the Delphi was extensive and took longer than expected, Round One was conducted between October and November 2024. Round Two took place in December 2024, and Round Three was conducted between January and February 2025. All panellists who took part in Round One were invited to Rounds Two and Three, regardless of whether they participated in Round Two. In each round, we asked Delphi panellists to individually rate the importance of each domain and each item to be included in a reporting guideline for intervention fidelity using a 9-point scale (1 to 3= ‘not important’, 4 to 6= ‘important but not essential’, 7 to 9= ‘very important’). Each domain and item was accompanied by a comment box for panellists to provide their rationale and suggestions (open-ended responses). These responses informed changes to the domains and items (e.g., merging or improving wording) in the subsequent round. A summary of the numerical group ratings (e.g., median and percentage of the full panel) and the rationale behind those ratings (i.e., anonymous panellists’ comments) was provided to panellists in Rounds Two and Three.

Consensus was defined as more than 70% of panellists rating a domain or item as 1 to 3, leading to exclusion, and 7 to 9, leading to inclusion in the reporting guideline, after at least two voting rounds. In the absence of a universally accepted definition of consensus in Delphi studies, this threshold (i.e., 70%) was defined a priori,^19^ taking into account the study aims, the heterogeneity of the panel and consistency with previous Delphi studies conducted to inform the development of reporting guidelines, such as CONSORT-ROUTINE and CONSORT-SURROGATE.^23^ ^24^ Domains and items were re-rated in Round Two, even if they had reached the consensus threshold in Round One. This process aimed to allow panellists to reflect on the group summary and anonymous comments from Round One, supporting more informed ratings, as outlined in our protocol.^19^ Domains and items that underwent significant changes based on panellists’ suggestions between rounds (Appendix 4) were also re-rated. Round Three included the remaining domains and items that did not reach consensus (i.e., ratings 4 to 6) in Round Two. A list of domains and items that had already reached consensus was shared with panellists for information purposes. Domains and items that did not reach the consensus threshold in Round Three were not included in the guideline.

### Data analysis

We used descriptive statistics to summarise panellists’ characteristics and numerical ratings. We conducted a thematic analysis of the qualitative responses (e.g., rationale for ratings).^25^ In each round, all qualitative survey responses were extracted from Qualtrics and uploaded to Atlas.ti,^26^ a qualitative research software. One researcher reviewed responses for each domain and item, identifying units of meaning and assigning codes. For example, responses suggesting that items should be merged would be coded as ‘Merging’, while those recommending item splitting would be coded as ‘Splitting’. These initial codes were then grouped into broader themes by the same researcher. The codes and themes were shared with the GDG members for feedback, decision-making, and refinement, as needed. Any disagreements were resolved through discussion among the team until majority agreement was reached.

### Patient and public involvement

Patients or the public were not systematically involved in the design, conduct, reporting, or dissemination of this methodological study. The domains and items reflect the perspectives of the Delphi panellists, including clinicians and researchers with relevant experience.

## Results

One hundred and twenty-four researchers were invited via Qualtrics, but nine emails bounced, resulting in 115 successfully sent invitations. Sixty-one panellists completed Round One, 55 completed Round Two, and 56 completed Round Three. This represents a retention rate of 90% for Round Two and 92% for Round Three, based on those who completed Round One. Panellists had diverse backgrounds and were from 21 countries, including six low- and middle-income countries (LMICs) (Table 1).

**Table 1.** ReFiND Delphi panellist characteristics in Round One (n=61).

| **Age (years)** | **N** | **%** |
| --- | --- | --- |
| 25-34 | 7 | 11 |
| 35-44 | 16 | 26 |
| 45-54 | 20 | 33 |
| 55-64 | 14 | 23 |
| 65 and over | 4 | 7 |
| **Gender** | **N** | **%** |
| Woman | 36 | 59 |
| Man | 25 | 41 |
| **Country of residence** | **N** | **%** |
| Australia | 7 | 11 |
| Belgium | 1 | 2 |
| Brazil | 3 | 5 |
| Canada | 2 | 3 |
| China | 1 | 2 |
| Denmark | 2 | 3 |
| Ethiopia | 2 | 3 |
| India | 1 | 2 |
| Ireland | 2 | 3 |
| Italy | 1 | 2 |
| Malta | 1 | 2 |
| Netherlands | 2 | 3 |
| New Zealand | 2 | 3 |
| South Africa | 3 | 5 |
| South Korea | 1 | 2 |
| Spain | 1 | 2 |
| Sri Lanka | 1 | 2 |
| Sweden | 1 | 2 |
| Switzerland | 1 | 2 |
| United Kingdom of Great Britain and Northern Ireland | 17 | 28 |
| United States of America | 9 | 15 |
| **Education** | **N** | **%** |
| Master's or equivalent | 3 | 5 |
| Doctorate (DPhil/PhD) or equivalent | 56 | 92 |
| Other<sup>a</sup> | 2 | 3 |
| **Academic position** | **N** | **%** |
| Research assistant/PhD student | 3 | 5 |
| Postdoctoral Research Fellow | 3 | 5 |
| Lecturer/Assistant Professor | 8 | 13 |
| Senior Lecturer/Senior Research Fellow | 7 | 11 |
| Associate Professor/Reader/Principal Lecturer | 11 | 18 |
| Professor | 22 | 36 |
| Other<sup>b</sup> | 7 | 11 |

| **Research roles<sup>1</sup>** | **N** | **%** |
| --- | --- | --- |
| Trial investigator | 43 | 70 |
| Trial methodologist | 23 | 38 |
| Trial assistant | 4 | 7 |
| Intervention provider within a trial | 12 | 20 |
| Implementation scientist | 31 | 51 |
| Evidence synthesis specialist | 11 | 18 |
| Statistician | 6 | 10 |
| Epidemiologist | 7 | 11 |
| Health economist | 1 | 2 |
| Journal Editor or Associate Editor | 19 | 31 |
| Journal Editorial Board member | 18 | 30 |
| Research funding agency member | 4 | 7 |
| Research ethics committee member | 7 | 11 |
| Clinician/Health or Allied Health Professional/Social worker | 18 | 30 |
| Other<sup>c</sup> | 6 | 10 |

| **Areas of Research<sup>1</sup>** | **N** | **%** |
| --- | --- | --- |
| Meta-Research/Research methodology (transparency, reproducibility, credibility) | 22 | 36 |
| Clinical trials | 36 | 59 |
| Implementation science | 36 | 59 |
| Health Services | 28 | 46 |
| Health Policy and Systems | 9 | 15 |
| Health Education | 11 | 18 |
| Health Ethics | 1 | 2 |
| Health Informatics | 2 | 3 |
| Digital Health | 18 | 30 |
| Public, Environmental and Occupational Health | 16 | 26 |
| Behavioural research | 27 | 44 |
| Psychology | 17 | 28 |
| Nursing/Midwifery | 8 | 13 |
| Social Work | 1 | 2 |
| Clinical Medicine | 11 | 18 |
| Rehabilitation | 21 | 34 |
| Sports Sciences | 8 | 13 |
| Complementary and Integrative Health | 4 | 7 |
| Other<sup>d</sup> | 4 | 7 |

| **Trial involvement level<sup>1</sup>** | **N** | **%** |
| --- | --- | --- |
| Intervention design | 54 | 89 |
| Feasibility and pilot trials | 52 | 85 |
| Explanatory or efficacy trials | 35 | 57 |
| Pragmatic or effectiveness trials | 44 | 72 |
| Effectiveness-implementation hybrid trials | 27 | 44 |
| Implementation trials | 22 | 36 |
| Other<sup>e</sup> | 3 | 5 |

| **Publications<sup>1</sup>** | **N** | **%** |
| --- | --- | --- |
| Author of a peer-reviewed publication related to reporting methods | 39 | 64 |
| Author of a peer-reviewed publication related to trials of non-drug, non-surgical interventions | 51 | 84 |
| Author of a peer-reviewed publication related to the process evaluation of non-drug, non-surgical interventions | 40 | 66 |
| Author of a peer-reviewed publication related to the fidelity of non-drug, non-surgical interventions | 50 | 82 |

<sup>1</sup>Panellists could select multiple options, so totals may exceed 100%.
Responses as reported by panellists:
<sup>a</sup>PhD Candidate, Master in Philosophy (MPhil) part of the PhD programme
<sup>b</sup>Senior Research Scientist, PhD Student and Senior Lecturer, Adjunct Research Assistant Professor, Research Coordinator, Clinician, Senior Researcher and Associate Professor, Head of Medical Affairs
<sup>c</sup>Academic, Non-profit Leader, Programme Manager, Research Officer, Behavioural Scientist and Research Funding Panel Member, Qualitative Researcher
<sup>d</sup>Pain Research, Dental Clinic, Global Public Health and Maternal Health, Psychometrics
<sup>e</sup>Process evaluation, Evaluations of health service changes, Translational Research

In Round One, panellists rated seven domains and 79 individual items (Figure 1). They provided 1,083 comments explaining their ratings or offering suggestions: 53 on specific open-ended questions, 142 on individual domains, and 888 on individual items. Seventy-five percent of panellists provided at least one comment. Domain Four (intervention receipt) was split into 4a (actual receipt) and 4b (participant understanding) to accommodate panellists’ feedback. From the original 79 items, 11 were merged and 4 new items were added based on panellists’ suggestions, resulting in 72 items for Round Two (Appendix 4, Table A1). Sixteen items were modified based on panellists’ feedback (Appendix 4, Table A2).

**Figure 1.**
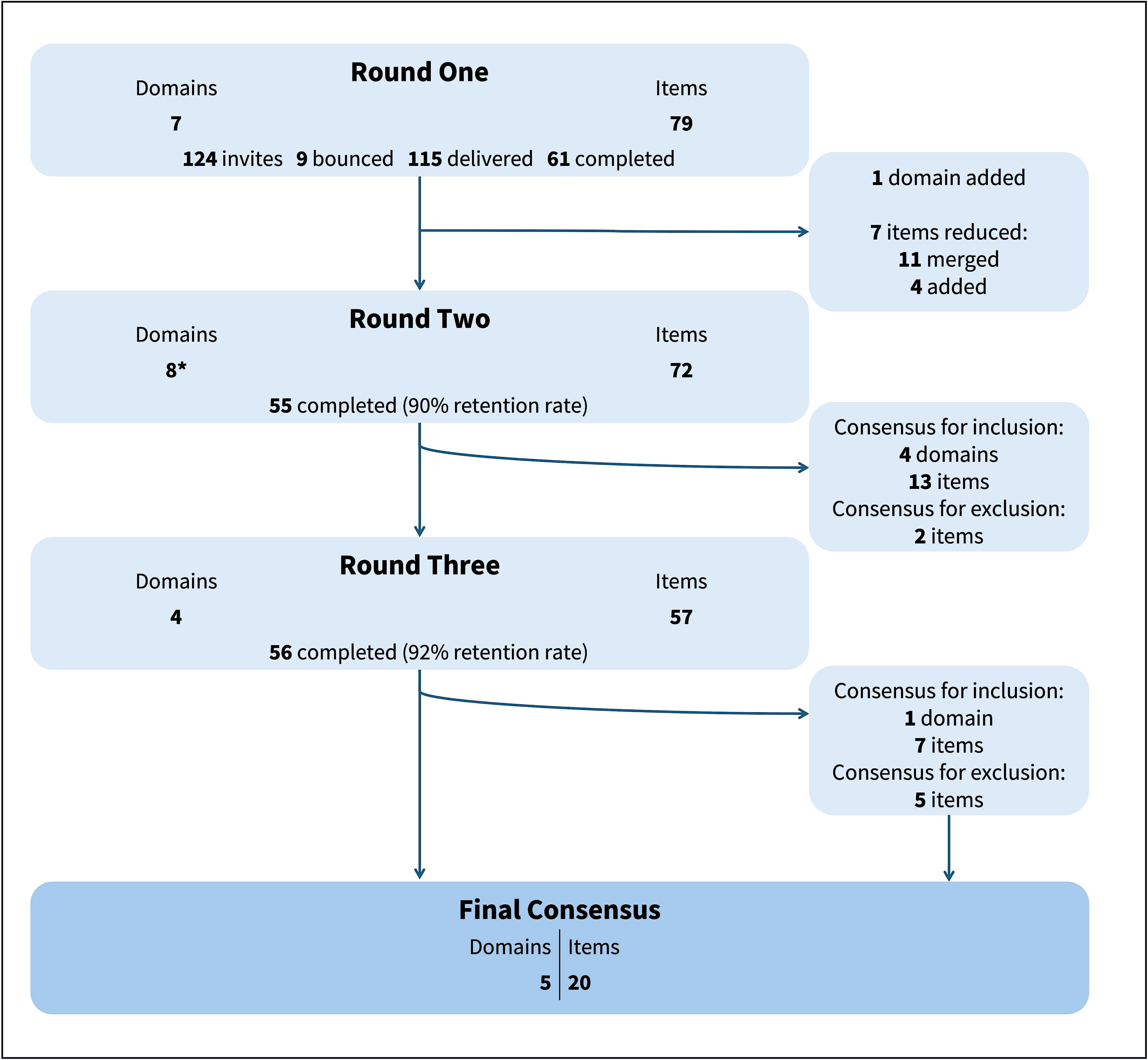
Flow diagram of the ReFiND Delphi process. *One domain (Intervention Receipt) was split into two after feedback from panellists in Round One, resulting in 8 domains instead of 7 in Round Two. Retention rate is based on participants completing Round One.

In Round Two, panellists rated eight domains and 72 items. Panellists provided 1,274 comments throughout the survey: 21 on specific open-ended questions, 162 on individual domains, and 1,091 on individual items. Over 80% of panellists provided at least one comment. After Round Two, four domains and 15 items achieved consensus: four domains and 13 items were included, while two items were excluded (Figure 1). The remaining four domains and 57 items that had not achieved consensus yet after Round Two were voted on in Round Three. Of these, one domain and seven items achieved consensus for inclusion, and five items for exclusion (Table 2 and 3). No significant changes were made to domains or items from Round Two to Round Three. We received 1,187 comments in Round Three. At the conclusion of the Delphi process, five domains and 20 items reached consensus for inclusion in ReFiND (Table 2 and 3). Domains and items that did not reach consensus are reported in the Appendix 5. Delphi surveys, including panellists’ comments from one round to another, are provided in Appendix 1-3.

**Table 2.** Ratings of intervention fidelity domains across Delphi rounds, ranked by percentage. Percentages meeting the consensus threshold (>70%) are shown in bold.

| Domain | Median (IQR) |  |  | % Ratings 1-3 |  |  | % Ratings 4-6 |  |  | % Ratings 7-9 |  |  | Consensus outcome |
| --- | --- | --- | --- | --- | --- | --- | --- | --- | --- | --- | --- | --- | --- |
|  | R1 | R2 | R3 | R1 | R2 | R3 | R1 | R2 | R3 | R1 | R2 | R3 |  |
| Intervention Delivery by Providers | 9 (8, 9) | 9 (9, 9) | C-R2* | 0.0 | 0.0 | C-R2 | 3.3 | 0.0 | C-R2 | 96.7 | **100.0** | C-R2 | INCLUDE |
| Intervention Fidelity Measurement Methods | 8 (7, 9) | 9 (8, 9) | C-R2 | 0.0 | 1.8 | C-R2 | 6.6 | 3.6 | C-R2 | 93.4 | **94.5** | C-R2 | INCLUDE |
| Intervention Design | 8 (7, 9) | 8 (6.5, 8) | C-R2 | 1.6 | 3.6 | C-R2 | 14.8 | 21.8 | C-R2 | 83.6 | **74.5** | C-R2 | INCLUDE |
| Provider Training | 8 (7, 9) | 8 (6, 8) | C-R2 | 1.6 | 1.8 | C-R2 | 21.3 | 25.5 | C-R2 | 77.0 | **72.7** | C-R2 | INCLUDE |
| Intervention Receipt by Participants - Actual receipt<sup>1</sup> | 8 (7, 9) | 8 (7, 9) | 8 (7, 9) | 3.3 | 7.3 | 5.4 | 9.8 | 16.4 | 8.9 | 86.9 | 76.4 | **85.7** | INCLUDE |
| Intervention Receipt by Participants - Understanding | NA | 7 (4, 8) | 5 (3, 7) | NA | 20.0 | 26.8 | NA | 27.3 | 41.1 | NA | 52.7 | 32.1 | NO CONSENSUS |
| Intervention Enactment by Participants | 7 (6, 8) | 5 (4, 7) | 5 (3, 7) | 6.6 | 23.6 | 35.7 | 29.5 | 45.5 | 37.5 | 63.9 | 30.9 | 26.8 | NO CONSENSUS |
| Moderators of Intervention Fidelity and Outcomes | 7 (5, 9) | 5 (3, 6) | 4 (2, 6) | 11.5 | 34.5 | 44.6 | 32.8 | 41.8 | 35.7 | 55.7 | 23.6 | 19.6 | NO CONSENSUS |

R1: Round One R2: Round Two R3: Round Three
Ratings 1-3: Not important Ratings 4-6: May be important but not essential Ratings 7-9: Very important
NA: Not Applicable – This domain was suggested by panelists in R1 and therefore included only in R2.
\*C-R2: Consensus reached in Round Two. Domains that achieved the consensus threshold (>70%) after being voted on R1 and R2 were not voted on again in R3.

**Table 3.**
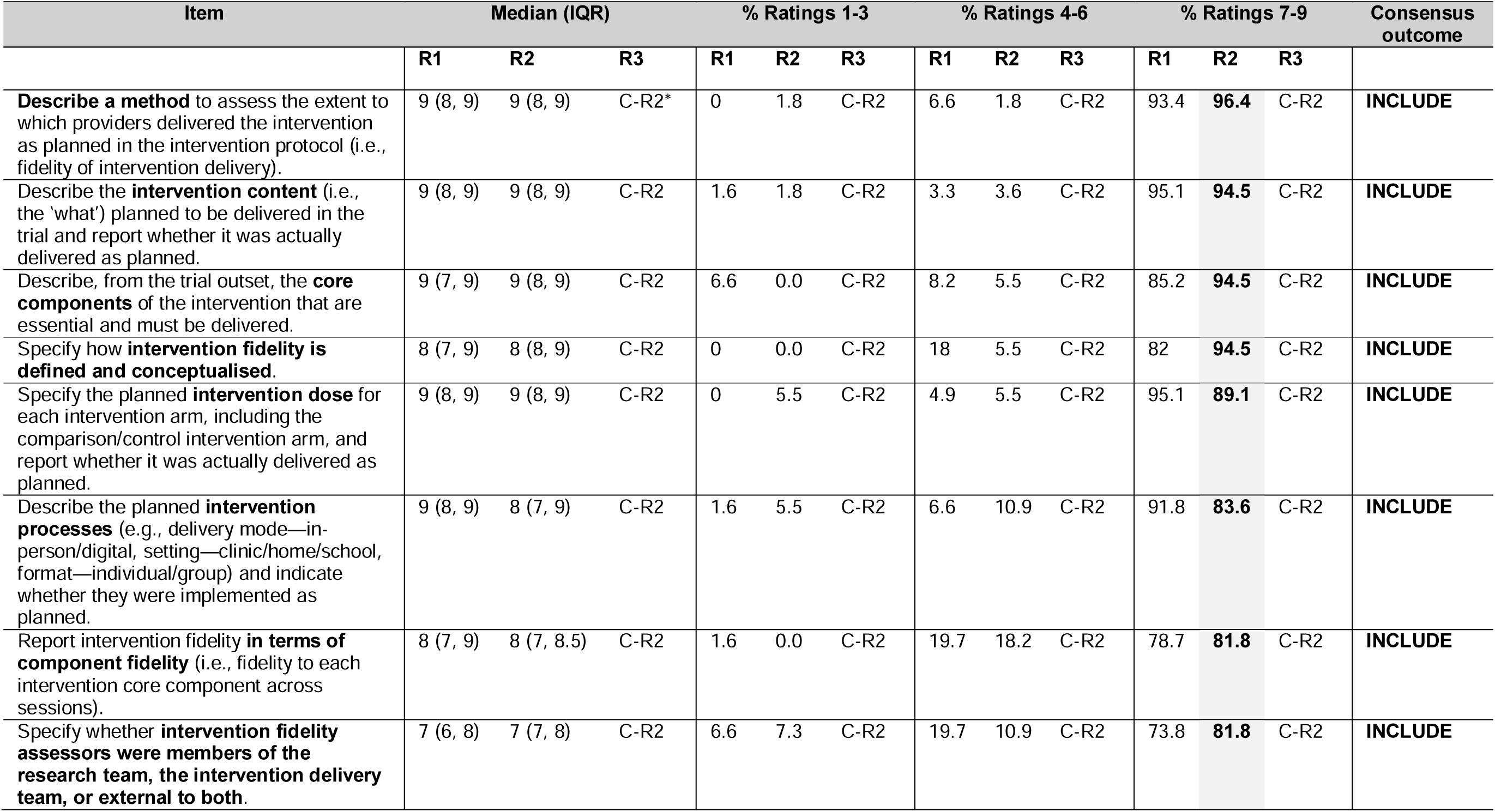
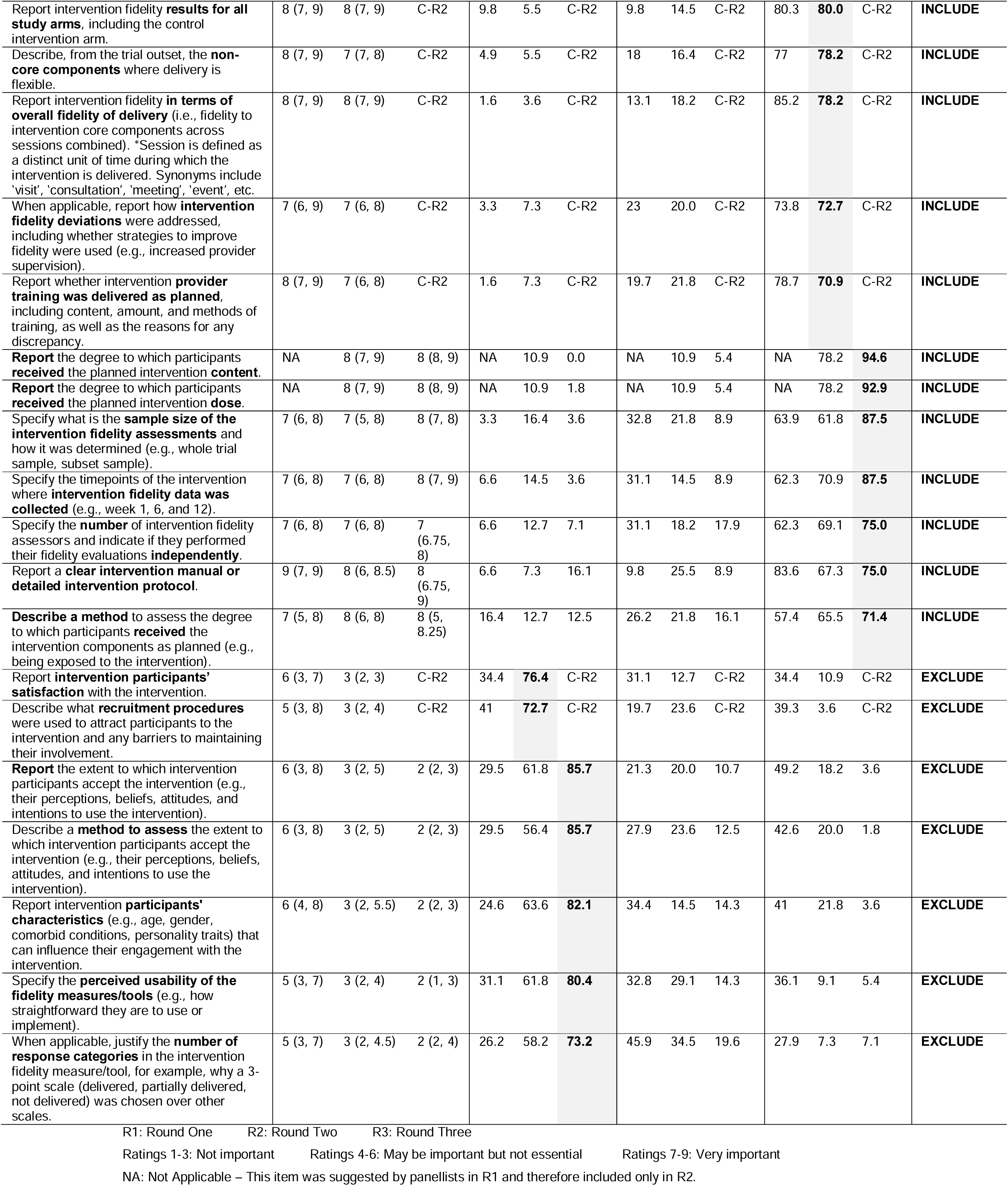
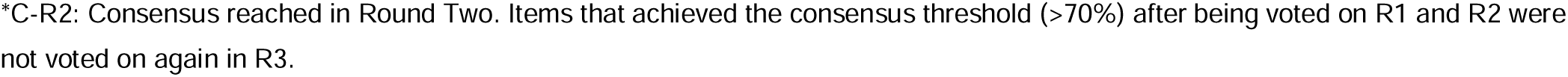
Ratings of intervention fidelity items meeting the >70% consensus threshold for inclusion (7-9) or exclusion (1-3) across Delphi rounds, ranked by percentage. Percentages meeting the consensus threshold (>70%) are shown in bold. See Appendix 5 for items that did not reach consensus.

In the qualitative analysis, we identified three themes: (1) suggestions for improvements in future rounds, (2) the need for clarifications about ReFiND scope and applicability, and (3) factors considered by panellists while rating the domains and items (Table 4). Suggestions from panellists (Theme 1) were discussed among the GDG members and implemented when deemed feasible and agreed upon. Examples of implemented suggestions include adding a ‘Not applicable’ option to some items and providing panellists with the full list of domains and items so they could cross-check information when making their individual ratings. Suggested clarifications identified in Theme 2, as well as factors influencing panellists’ decision-making (Theme 3), helped inform the elaboration of the ReFiND statement (currently under review).

**Table 4.**
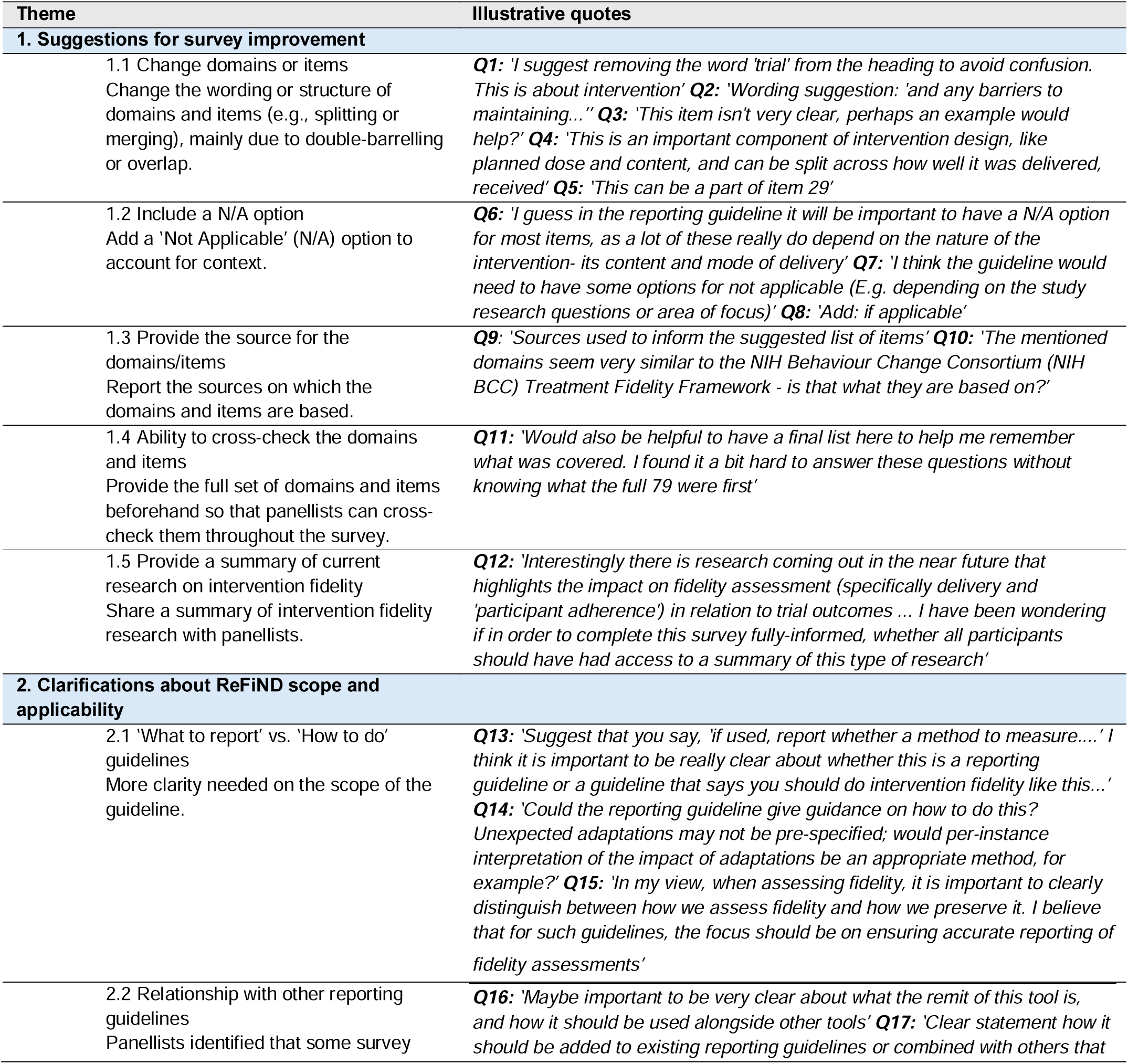
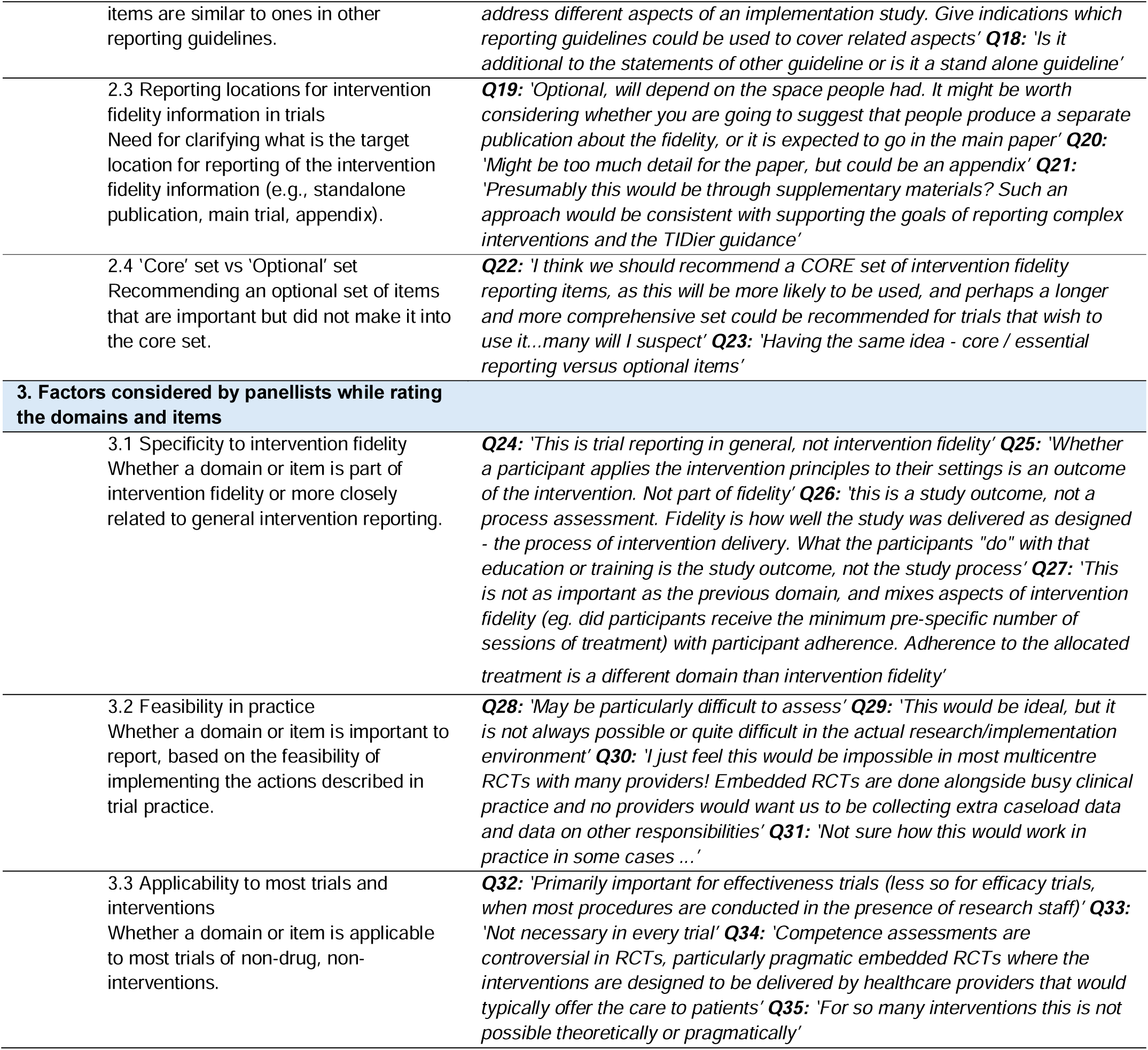
Themes identified from panellists’ comments across the Delphi rounds.

## Discussion

Through this Delphi study, international experts achieved consensus on five domains of intervention fidelity and 20 reporting items, which informs ReFiND, a reporting guideline for intervention fidelity in non-drug, non-surgical trials.^19^ Our findings highlight the importance of complete reporting of intervention design details, so that both the standard against which fidelity is assessed and the methods used to assess fidelity are transparent. In addition to fidelity of intervention delivery, the most commonly reported domain,^12^ panellists also reached consensus on the importance of reporting whether provider training was delivered as planned, and the extent to which participants received the planned intervention components and dose. While these findings reflect to some extent the multidimensional nature of intervention fidelity, extending beyond delivery, not all domains were widely accepted.

The domains of participant understanding (a component of intervention receipt) and enactment did not meet the consensus threshold. These domains are recommended by some frameworks, such as the National Institutes of Health/Behaviour Change Consortium (NIH/BCC) Fidelity Framework,^9^ ^11^ ^13^ but their inclusion under the intervention fidelity ‘umbrella’ remains debated.^2^ ^27^ During our Delphi process, many researchers argued that whether a participant understands the intervention concepts or applies them in daily life is better conceptualised as moderators or outcomes of the intervention, rather than as components of fidelity itself. Conversely, other researchers viewed enactment as an important domain of intervention fidelity, as it provides insights into the mechanisms through which outcomes are achieved. Although the importance of participant understanding and enactment were generally acknowledged, there was no consensus on whether these should be reported as part of intervention fidelity. These findings suggest that, at present, we cannot recommend the assessment and reporting of these components as essential standards for intervention fidelity, mainly due to the lack of a unified understanding. However, participant understanding and enactment remain important to trials more broadly, particularly those involving certain types of interventions such as behavioural ones. Researchers should consider reporting these components where relevant, while recognising that they may not fall within the scope of intervention fidelity. As the field evolves, this understanding may improve, and in the future a consensus may be reached.

Our qualitative findings indicate that panellists considered multiple factors when rating, such as the feasibility of implementing recommendations in practice and the relevance of domains and items to most non-drug, non-surgical trials. This helps explain why many items did not reach consensus for inclusion, as ratings often reflected a balance between perceived importance and broader concerns around feasibility and generalisability. Despite these considerations and the focus on only essential items for inclusion (the core set), 20 items achieved consensus. This number aligns with that found in other reporting guidelines^28^ ^29^ but may still be challenging to fully report in main trial reports due to word limits. One option is to report this information outside the main trial report, such as in appendices or publicly available repositories, with a clear reference in the main text indicating where the information can be accessed. Additionally, independent publications focusing on intervention fidelity findings have become increasingly common and can be used for full reporting.^30–32^

### Strengths, limitations, and considerations

We involved a diverse group of experts from various countries, including LMICs, and from different academic seniority levels and research roles, such as journal editors, members of research funding agencies, and intervention providers within trials, each playing a crucial role in intervention fidelity. Many panellists were identified through a list of authors of guidance documents on intervention fidelity.^12^ Given the heterogeneity of recommendations across these documents, inviting panellists from this list helped to ensure the inclusion of participants with differing views on intervention fidelity. This contributed to mitigating the risks of panel stacking^33^ (i.e., the disproportionate recruitment of panellists representing similar views) and fostered a broader debate. Interestingly, we noted a predominance of panellists from the UK, a trend also observed in other Delphi studies developed to inform reporting guidelines.^34^ ^35^ Additionally, we included nearly all recommendations from the intervention fidelity literature in the Delphi process, even those that, in our view, initially fell outside the core scope of fidelity (e.g., items related to recruitment). This inclusive approach allowed us, for the first time, to identify through a systematic consensus process which recommendations may be considered essential and which may be unrelated to intervention fidelity.

In addition to its diversity, the panel in this study was highly engaged. We received thousands of comments from panellists across the survey rounds. To promote informed ratings, we shared all panellists’ comments from the previous round in Rounds Two and Three, excluding only redundant comments. Their feedback supported item revisions between rounds and provided valuable context for their ratings. Panellists raised important considerations that helped inform the explanations and elaborations in the ReFiND statement, which will be published separately, and shaped its overall scope and usability. Other strengths of this study include its prospective registration and publication of the protocol in a peer-reviewed journal,^19^ as well as the high retention rates in the second and third rounds of the Delphi process (90% and 91%, respectively).

This study has some limitations. First, only about half of those invited participated in Round One. While other studies have reported similar rates throughout their processes,^36^ ^37^ one possible reason for this is that we sent direct invitations rather than pre-registering interest and inviting only those who had expressed willingness to participate. Additionally, we did not include partial or incomplete responses. Second, the surveys were lengthy, which could have introduced fatigue effects on panellists. However, we mitigated this by randomising the order of the survey questions. Third, initial coding of qualitative responses was done by a single assessor. Fourth, we are unable to determine whether the changes made to the published protocol, as reported in the Methods section, could have influenced the findings of this study. We are also unable to assess whether the consensus outcomes would remain stable under different consensus thresholds, as no sensitivity analyses were preplanned for this purpose.

The findings of this Delphi study were discussed in online consensus meetings with the panellists and the ReFiND guideline development group to conceptualise the guideline statement with explanations and elaboration document, which will be disseminated soon (currently under review). These discussions helped support that the guideline is clear, easy to read, relevant to different disciplines, and accessible to a range of users. Once published, ReFiND will be disseminated through multiple strategies, including the development of a dedicated website, inclusion in the EQUATOR Network library, and presentations in webinars and conferences.

## Conclusion

International experts reached consensus on five domains and 20 reporting items for inclusion in ReFiND, a reporting guideline for intervention fidelity in non-drug, non-surgical trials. These findings contribute to the transparent reporting of intervention fidelity, facilitating better replication of effective interventions by researchers and clinicians.

## Contributors

FS: conceptualisation, methodology, investigation, writing – original draft, writing – review & editing, project administration. MKF: conceptualisation, methodology, investigation, writing – review & editing, supervision. TH: conceptualisation, methodology, writing – review & editing, supervision. BB: conceptualisation, methodology, writing – review & editing. CC: conceptualisation, methodology, writing – review & editing. CM: conceptualisation, methodology, writing – review & editing. DCR: conceptualisation, methodology, writing – review & editing. JMF: conceptualisation, methodology, writing – review & editing. MU: conceptualisation, methodology, writing – review & editing. NEF: conceptualisation, methodology, writing – review & editing. SEL: conceptualisation, methodology, writing – review & editing. ZMS: conceptualisation, methodology, writing – review & editing. PM: conceptualisation, methodology, investigation, writing – review & editing, project administration, supervision. FS is responsible for the overall content as guarantor.

## Data Availability

All data produced in the present work are contained in the manuscript

