## Appendix 5 for "Establishing consensus on domains and items for Reporting intervention Fidelity in Non-Drug, non-surgical trials: the ReFiND Delphi study"

**Appendix 5: Items that did not reach consensus**

**Table A3.** Items that did not reach the Delphi consensus threshold (>70% agreement).

| **Item** | **Median (IQR)** | | | **% 1-3** | | | **% 4-6** | | | **% 7-9** | | | **Consensus Outcome** |
| --- | --- | --- | --- | --- | --- | --- | --- | --- | --- | --- | --- | --- | --- |
|  | **R1** | **R2** | **R3** | **R1** | **R2** | **R3** | **R1** | **R2** | **R3** | **R1** | **R2** | **R3** |  |
| **Report whether** unintended components (e.g., components that are unnecessary or unhelpful) were delivered. | 7 (5, 9) | 7 (5, 8) | 7 (5, 8) | 8.2 | 14.5 | 12.5 | 34.4 | 32.7 | 19.6 | 57.4 | 52.7 | 67.9 | NO CONSENSUS |
| Report whether **adaptations made** to the intervention are consistent with the flexibility defined in the protocol, if applicable. | 7 (7, 9) | 7 (5, 8) | 7 (5, 8) | 4.9 | 16.4 | 12.5 | 19.7 | 21.8 | 19.6 | 75.4 | 61.8 | 67.9 | NO CONSENSUS |
| If applicable, specify the rate of **missing intervention fidelity data** and how it was managed. | 7 (6, 8) | 7 (5.5, 8) | 7 (5, 8) | 6.6 | 7.3 | 8.9 | 27.9 | 32.7 | 25.0 | 65.6 | 60.0 | 66.1 | NO CONSENSUS |
| Describe a **specific** **intervention fidelity assessment and monitoring protocol** prior to trial initiation/trial analysis. | 7 (6, 9) | 7 (5, 8) | 7 (5, 8) | 4.9 | 3.6 | 7.1 | 37.7 | 40.0 | 32.1 | 57.4 | 56.4 | 60.7 | NO CONSENSUS |
| If not all intervention fidelity domains were assessed, describe the rationale for selecting the **assessed domains**. | 7 (6, 8) | 7 (5, 8) | 7 (5, 8) | 4.9 | 5.5 | 16.1 | 32.8 | 34.5 | 23.2 | 62.3 | 60.0 | 60.7 | NO CONSENSUS |
| Specify when **intervention fidelity data was analysed**, such as concurrently with the intervention or after the intervention period was completed. | 8 (6, 9) | 7 (4, 7) | 7 (5, 8) | 4.9 | 23.6 | 17.9 | 26.2 | 25.5 | 26.8 | 68.9 | 50.9 | 55.4 | NO CONSENSUS |
| Specify the **type of statistical analysis conducted** to obtain intervention fidelity results (e.g., descriptive, ANOVA, regression). | 7 (6, 9) | 6 (5, 8) | 7 (4.75, 8) | 3.3 | 10.9 | 17.9 | 29.5 | 40.0 | 30.4 | 67.2 | 49.1 | 51.8 | NO CONSENSUS |
| Report the degree to which the intervention delivered to intervention participants is **congruent with its underlying theory or logic model**. | 7 (5, 9) | 7 (5, 8) | 6.5 (3.75, 8) | 9.8 | 18.2 | 25.0 | 27.9 | 30.9 | 25.0 | 62.3 | 50.9 | 50.0 | NO CONSENSUS |
| Report the extent to which the **actual intervention providers in the trial met the provider selection criteria or expected characteristics** described in the protocol (e.g., training, experience, qualifications). | 5 (5, 7) | 7 (5, 8) | 6.5 (4, 8) | 23 | 20.0 | 19.6 | 49.2 | 20.0 | 30.4 | 27.9 | 60.0 | 50.0 | NO CONSENSUS |
| Specify an a priori threshold for **acceptable level of intervention fidelity** or cut-offs. | 7 (5, 8) | 7 (5, 8) | 6 (4, 8) | 14.8 | 9.1 | 19.6 | 19.7 | 38.2 | 32.1 | 65.6 | 52.7 | 48.2 | NO CONSENSUS |
| Describe any actions taken to minimise **selection bias when sampling a sub-set** of data points for intervention fidelity analysis. | 7 (6, 8) | 6 (5, 8) | 6 (5, 8) | 4.9 | 16.4 | 10.7 | 29.5 | 38.2 | 42.9 | 65.6 | 45.5 | 46.4 | NO CONSENSUS |
| Specify the **level of blinding** of intervention fidelity assessors at the time of intervention fidelity assessments (e.g., blinded to intervention outcomes or intervention provider identity). | 7 (6, 8) | 6 (5, 7) | 6 (4, 7) | 9.8 | 9.1 | 10.7 | 26.2 | 43.6 | 42.9 | 63.9 | 47.3 | 46.4 | NO CONSENSUS |
| Specify the **frameworks and guidance** used to inform intervention fidelity assessments. | 7 (6, 8) | 7 (5, 8) | 6 (4, 8) | 6.6 | 7.3 | 16.1 | 36.1 | 40.0 | 39.3 | 57.4 | 52.7 | 44.6 | NO CONSENSUS |
| When applicable, describe **contextual factors** assessed or observed by the research team that may have influenced intervention fidelity assessment or results (e.g., intervention complexity, intervention provider caseload, time spent by providers on small talk vs time delivering the intervention as intended, etc). | 6 (4, 8) | 7 (4.5, 8) | 6 (4, 8) | 19.7 | 23.6 | 19.6 | 32.8 | 20.0 | 37.5 | 47.5 | 56.4 | 42.9 | NO CONSENSUS |
| Describe an **intervention provider training plan** including training content, amount, and methods used to deliver the training (e.g, training manuals, didactic sessions, role modelling, supervision with feedback). | 7 (6, 9) | 7 (4.5, 8) | 6 (4, 7) | 3.3 | 21.8 | 21.4 | 24.6 | 21.8 | 39.3 | 72.1 | 56.4 | 39.3 | NO CONSENSUS |
| Report intervention fidelity **in terms of session fidelity** (i.e., fidelity to intervention core components within sessions). *Session is defined as a distinct unit of time during which the intervention is delivered. Synonyms include ‘visit’, ‘consultation’, ‘meeting’, ‘event’, etc. | 7 (5, 8) | 6 (5, 8) | 6 (5, 7) | 9.8 | 3.6 | 8.9 | 36.1 | 58.2 | 51.8 | 54.1 | 38.2 | 39.3 | NO CONSENSUS |
| **Report participant adherence** to the intervention (i.e., degree to which the participant **follows the intervention as instructed/required**). | 8 (7, 9) | 6 (2, 7) | 5 (2, 7.25) | 13.1 | 41.8 | 41.1 | 9.8 | 12.7 | 21.4 | 77 | 45.5 | 37.5 | NO CONSENSUS |
| **Report** provider competence and proficiency to deliver the intervention (e.g., comprehension of the intervention components, demonstration of relevant skills required to deliver the intervention, etc). | 7 (6, 8) | 5 (4.5, 8) | 5 (3.75, 7.25) | 4.9 | 23.6 | 25.0 | 29.5 | 30.9 | 37.5 | 65.6 | 45.5 | 37.5 | NO CONSENSUS |
| Describe how intervention fidelity **measures/tools were developed**. If existing measures/tools are used, specify them and provide their source. | 7 (6, 8) | 5 (3, 7) | 4.5 (2, 7) | 8.2 | 34.5 | 39.3 | 31.1 | 25.5 | 23.2 | 60.7 | 40.0 | 37.5 | NO CONSENSUS |
| Describe how **data from multiple intervention fidelity measurement methods**, when applicable, are integrated and triangulated to provide comprehensive fidelity results. For example, describe how quantitative data (e.g., from fidelity measures/tools) are combined with qualitative data (e.g., self-reports). | 7 (6, 9) | 6 (5, 8) | 5 (4, 7) | 8.2 | 9.1 | 19.6 | 31.1 | 47.3 | 42.9 | 60.7 | 43.6 | 37.5 | NO CONSENSUS |
| Describe a **method to assess** whether unintended components are delivered (e.g., components that are unnecessary or unhelpful). | 7 (5, 9) | 6 (5, 8) | 5 (4, 7) | 8.2 | 12.7 | 23.2 | 31.1 | 43.6 | 41.1 | 60.7 | 43.6 | 35.7 | NO CONSENSUS |
| Report whether **booster (i.e., additional) training sessions** were offered and delivered to intervention providers. | 7 (6, 8) | 6 (5, 7) | 5 (3, 7) | 4.9 | 14.5 | 26.8 | 27.9 | 36.4 | 41.1 | 67.2 | 49.1 | 32.1 | NO CONSENSUS |
| **Describe a method** **to assess** **participant adherence** to the intervention (i.e., degree to which the participant **follows the intervention as instructed/required**). | 8 (6, 9) | 5 (2, 7) | 4 (2, 7) | 13.1 | 45.5 | 46.4 | 14.8 | 16.4 | 23.2 | 72.1 | 38.2 | 30.4 | NO CONSENSUS |
| Report the level of training **received by intervention providers** (e.g., frequency of training, attendance to training sessions). | 7 (6, 8) | 5 (5, 7) | 5 (4, 7) | 6.6 | 10.9 | 21.4 | 39.3 | 52.7 | 50.0 | 54.1 | 36.4 | 28.6 | NO CONSENSUS |
| **Report** the degree to which participants **understood/comprehended** the intervention components as planned. | 7 (5, 8) | 5 (3, 7) | 3.5 (2, 6.25) | 14.8 | 34.5 | 50.0 | 26.2 | 27.3 | 25.0 | 59 | 38.2 | 25.0 | NO CONSENSUS |
| **Describe a method** to assess the degree to which intervention participants **apply the intervention** **in real-life settings** (also referred to as 'enactment’). | 6 (4, 8) | 4 (2, 6.5) | 3 (2, 6) | 21.3 | 47.3 | 55.4 | 36.1 | 27.3 | 23.2 | 42.6 | 25.5 | 21.4 | NO CONSENSUS |
| **Describe a method** to assess the degree to which participants **understood/comprehended** the intervention components as planned. | NA | 5 (3, 7) | 3 (2, 6) | NA | 40.0 | 51.8 | NA | 23.6 | 28.6 | NA | 36.4 | 19.6 | NO CONSENSUS |
| Describe how intervention fidelity **assessors were trained** to use the intervention fidelity measures/tools and conduct the fidelity assessments. | 7 (5, 8) | 5 (4.5, 7) | 5 (5, 6) | 8.2 | 18.2 | 10.7 | 37.7 | 50.9 | 69.6 | 54.1 | 30.9 | 19.6 | NO CONSENSUS |
| If applicable, describe the **rationale for any sensitivity/additional analysis** informed by intervention fidelity data (e.g., intention-to-treat versus removing low fidelity cases, mediation analysis based on fidelity to specific intervention components). | 6 (4, 8) | 5 (2, 7) | 4 (2, 5) | 21.3 | 30.9 | 44.6 | 29.5 | 36.4 | 35.7 | 49.2 | 32.7 | 19.6 | NO CONSENSUS |
| **Report** how well intervention participants **enacted** the intervention **content**, meaning the degree to which they **applied it in real-life settings**. | 6 (5, 8) | 3 (2, 6) | 3 (2, 5.25) | 18 | 50.9 | 58.9 | 36.1 | 29.1 | 23.2 | 45.9 | 20.0 | 17.9 | NO CONSENSUS |
| **Describe a method** to assess provider competence and proficiency to deliver the intervention (e.g., assessment of knowledge and skills). | 7 (5, 8) | 5 (4, 8) | 5 (3, 6) | 6.6 | 23.6 | 28.6 | 36.1 | 40.0 | 53.6 | 57.4 | 36.4 | 17.9 | NO CONSENSUS |
| Describe the **strengths and limitations** of the intervention fidelity assessment methods used. | 6 (4, 8) | 5 (3, 6) | 5 (2, 6) | 19.7 | 32.7 | 33.9 | 32.8 | 45.5 | 48.2 | 47.5 | 21.8 | 17.9 | NO CONSENSUS |
| Specify how the intervention fidelity **measures/tools align with the intervention content and activities**. | 7 (5, 8) | 6 (4, 7) | 5 (3, 6) | 14.8 | 12.7 | 26.8 | 29.5 | 50.9 | 57.1 | 55.7 | 36.4 | 16.1 | NO CONSENSUS |
| Specify whether the intervention fidelity **measures/tools were piloted**. | 6 (4, 7) | 4 (3, 5) | 3 (2, 5) | 18 | 40.0 | 53.6 | 47.5 | 45.5 | 32.1 | 34.4 | 14.5 | 14.3 | NO CONSENSUS |
| If **different weights were assigned to items** in an intervention fidelity measure/tool that has not been externally validated, report how these weights were determined (e.g., theory, empirical evidence, sensitivity analysis) and the rationale for this approach. | 6 (5, 8) | 5 (4, 6) | 4 (2, 5.25) | 13.1 | 21.8 | 42.9 | 42.6 | 60.0 | 42.9 | 44.3 | 18.2 | 14.3 | NO CONSENSUS |
| Specify how delivery contamination **between intervention arms** was avoided. | 7 (5, 9) | 5 (3, 7) | 3 (2, 4) | 16.4 | 41.8 | 66.1 | 16.4 | 29.1 | 21.4 | 67.2 | 29.1 | 12.5 | NO CONSENSUS |
| Report whether the **planned number of intervention providers** to deliver the intervention within the trial was achieved. | 6 (4, 7) | 5 (2.5, 7) | 3 (2, 5) | 23 | 43.6 | 57.1 | 34.4 | 29.1 | 30.4 | 42.6 | 27.3 | 12.5 | NO CONSENSUS |
| Describe any revisions to intervention fidelity assessments based on findings from the **intervention development process (e.g., piloting the intervention)**. | 5 (3, 7) | 5 (2, 7) | 4 (2, 5) | 29.5 | 36.4 | 44.6 | 36.1 | 36.4 | 42.9 | 34.4 | 27.3 | 12.5 | NO CONSENSUS |
| **Specify the** **psychometric properties** of the intervention fidelity measures/tools (e.g., reliability, validity, and sensitivity to change). | 7 (5, 8) | 5 (3, 5.5) | 4 (2, 6) | 11.5 | 32.7 | 41.1 | 36.1 | 47.3 | 46.4 | 52.5 | 20.0 | 12.5 | NO CONSENSUS |
| Specify the benchmarks used to **interpret the psychometrics** of the intervention fidelity measures/tools. For example, report what constitutes an acceptable level of inter-rater reliability. | 6 (3, 7) | 4 (3, 6) | 3 (2, 5) | 29.5 | 45.5 | 64.3 | 24.6 | 34.5 | 23.2 | 45.9 | 20.0 | 12.5 | NO CONSENSUS |
| Report whether the **cultural and linguistic aspects of the intervention** were delivered as planned, if applicable. | 6 (5, 8) | 5 (3, 7) | 4 (2, 5) | 19.7 | 27.3 | 48.2 | 31.1 | 45.5 | 39.3 | 49.2 | 27.3 | 12.5 | NO CONSENSUS |
| **Report** how well intervention participants **enacted** the intervention **dose**, meaning the degree to which they **applied it in real-life settings**. | NA | 2 (2, 5) | 2.5 (2, 4.25) | NA | 61.8 | 67.9 | NA | 29.1 | 25.0 | NA | 9.1 | 7.1 | NO CONSENSUS |
| **Describe each item** in the intervention fidelity measure/tool and the **process of their selection or development.** | 6 (3, 7) | 4 (3, 5) | 3 (2, 5) | 31.1 | 45.5 | 66.1 | 39.3 | 41.8 | 26.8 | 29.5 | 12.7 | 7.1 | NO CONSENSUS |
| Specify if there was any requirement for the **selection of intervention fidelity assessors** (e.g., be a health registered professional). | 6 (5, 7) | 5 (3, 6) | 5 (3, 5) | 11.5 | 34.5 | 35.7 | 42.6 | 47.3 | 57.1 | 45.9 | 18.2 | 7.1 | NO CONSENSUS |
| Describe how **stakeholders** were involved in the development of the intervention fidelity measures/tools. | 5 (4, 7) | 4 (2.5, 5) | 3 (2, 4) | 21.3 | 41.8 | 64.3 | 45.9 | 45.5 | 30.4 | 32.8 | 12.7 | 5.4 | NO CONSENSUS |
