## Appendix 4 for "Establishing consensus on domains and items for Reporting intervention Fidelity in Non-Drug, non-surgical trials: the ReFiND Delphi study"

**Appendix 4: Item changes between rounds**

From the original 79 items included in Round One, 11 were merged and 4 new items were added based on panellists’ suggestions (Table A1), resulting in 72 items for Round Two. The wording of 16 items was modified from Round One to Round Two based on panellists’ feedback (Table A2).

**Table A1.** Items that were merged or added based on panellists’ feedback.

| **Original item in Round One** | **Rationale for change based on panellists’ feedback** | **Outcome** |
| --- | --- | --- |
| Describe if the level of interactions that providers had with participants was consistent across trial arms and among all participants within each arm. | This item is about the level of attention providers gave to participants and could be included as an example of contextual factor rather than a single item | MERGED into the following existing item:  When applicable, describe contextual factors assessed or observed by the research team that may have influenced intervention fidelity assessment or results (e.g., intervention complexity, intervention provider caseload, time spent by providers on small talk vs time delivering the intervention as intended, etc). |
| Describe how intervention complexity was considered when planning and conducting fidelity assessments. | This item aligns with the existing item related to contextual factors | MERGED into the following existing item:  When applicable, describe contextual factors assessed or observed by the research team that may have influenced intervention fidelity assessment or results (e.g., intervention complexity, intervention provider caseload, time spent by providers on small talk vs time delivering the intervention as intended, etc). |
| Describe a method to determine whether adaptations made during intervention implementation fall within the allowed flexibility specified in the protocol or compromise fidelity. | This item overlaps with the item related to deviations | MERGED into the following existing item:  When applicable, report how intervention fidelity deviations were addressed, including whether strategies to improve fidelity were used (e.g., increased provider supervision). |
| At the trial outset, specify the minimum criteria for selecting trial providers. This can be based on their characteristics (e.g., motivation, flexibility, etc) and qualifications, including education, experience, and credentials. | This item is too detailed and overlaps with the previous item | MERGED into the following existing item:  Report the extent to which the actual intervention providers in the trial met the provider selection criteria or expected characteristics described in the protocol (e.g., training, experience, qualifications). |
| Report providers characteristics (e.g. years of experience, gender and race/ethnicity) and qualifications (i.e., education, experience, credentials). | This item is too similar to the previous item and race/ethnicity may not be important | MERGED into the following existing item:  Report the extent to which the actual intervention providers in the trial met the provider selection criteria or expected characteristics described in the protocol (e.g., training, experience, qualifications). |
| Report provider training according to a well-defined, a priori competence criterion (e.g., adherence to 80% of the intervention components). | This item can be confusing and is related to the criteria used to assess provider competence, which is covered in another item | MERGED into the following existing item:  Describe a method to assess provider competence and proficiency to deliver the intervention (e.g., assessment of knowledge and skills). |
| Describe a method to measure provider skill acquisition and maintenance after training, as well as their understanding of the treatment components, including the underlying rationale and theory. | The description of the item should be clearer, and it overlaps with a previous item | MERGED into the following existing item:  Describe a method to assess provider competence and proficiency to deliver the intervention (e.g., assessment of knowledge and skills). |
| Report provider skill acquisition and maintenance after training, including their understanding of the treatment components and the underlying rationale and theory. | This item overlaps with a previous item, which is simpler | MERGED into the following existing item:  Report provider competence and proficiency to deliver the intervention (e.g., comprehension of the intervention components, demonstration of relevant skills required to deliver the intervention, etc). |
| Specify the provider-participant ratios, caseloads and any other responsibilities that providers had in addition to their participant caseload that may impact the amount of time they can spend on intervention delivery. | This item is related to factors influencing intervention fidelity. It could be added to the item about context | MERGED into the following existing item:  When applicable, describe contextual factors assessed or observed by the research team that may have influenced intervention fidelity assessment or results (e.g., intervention complexity, intervention provider caseload, time spent by providers on small talk vs time delivering the intervention as intended, etc). |
| Specify if a component analysis was conducted (e.g., identification of which intervention components are critical for producing effects). | This item needs more clarity and overlaps with a previous item | MERGED into the following existing item:  If applicable, describe the rationale for any sensitivity/additional analysis informed by intervention fidelity data (e.g., intention-to-treat versus removing low fidelity cases, mediation analysis based on fidelity to specific intervention components). |
| Report any enhancement strategies employed to address low fidelity, if applicable. | This item is unclear and could be combined with the item related to deviations | MERGED into the following existing item:  When applicable, report how intervention fidelity deviations were addressed, including whether strategies to improve fidelity were used (e.g., increased provider supervision). |
| NA | Content and dose should be crossed with delivery, receipt, and enactment | NEW ITEM created:  Report the degree to which participants received the planned intervention content. |
| NA | Content and dose should be crossed with delivery, receipt, and enactment | NEW ITEM created:  Report the degree to which participants received the planned intervention dose. |
| NA | Actual receipt is a concept different from participant understanding/comprehension | NEW ITEM created:  Describe a method to assess the degree to which participants understood/comprehended the intervention components as planned. |
| NA | Content and dose should be crossed with delivery, receipt, and enactment | NEW ITEM created:  Report how well intervention participants enacted the intervention dose, meaning the degree to which they applied it in real-life settings. |

**Table A2.** Item modification from Round One to Round Two.

| **Original item in Round One** | **Revised item** | **Actions/Rationale for modifications** |
| --- | --- | --- |
| Describe a method to assess the degree to which participants received and understood the information/concepts/content provided in the intervention. | Describe a method to assess the degree to which participants received the intervention components as planned (e.g., being exposed to the intervention). | The item should focus on a single concept, which is actual receipt. Participant understanding is addressed in a separate item. Added ‘as planned’ for consistency throughout |
| Report the degree to which participants received and understood the information/concepts/content provided in the intervention. | Report the degree to which participants understood/comprehended the intervention components as planned. | The item should focus on a single concept, which is participant understanding. Actual receipt is addressed in a separate item (see above). Added ‘as planned’ for consistency throughout |
| Report the degree to which participants applied the intervention skills into their real life (also called 'enactment', 'engagement' or 'participant responsiveness'). | Report how well intervention participants enacted the intervention content, meaning the degree to which they applied it in real-life settings. | Used ‘how well’ to reflect both quality and quantity |
| Describe a method to assess participant adherence to the intervention. | Describe a method to assess participant adherence to the intervention (i.e., degree to which the participant follows the intervention as instructed/required). | Added a definition in brackets for clarity |
| Report participant adherence to the intervention. | Report participant adherence to the intervention (i.e., degree to which the  participant follows the intervention as instructed/required). | Added a definition in brackets for clarity |
| Describe the degree of flexibility permitted for different intervention aspects, such as in intervention manuals, provider training (e.g., adapting to learning styles and experience), intervention delivery (e.g., adapting to patient type and motivation), receipt (e.g., adapting to learning style and health literacy), and enactment (e.g., adapting to social and environmental context). | Report whether adaptations made to the intervention are consistent with the flexibility defined in the protocol, if applicable. | The item was simplified. Examples were removed as they added unnecessary complexity |
| When selecting trial providers, describe whether there is a good fit between the potential provider, the population (i.e., matching to key characteristics such as ethnicity) and the intervention (i.e., providers buy into the intervention theory and foundations). | Report the extent to which the actual intervention providers in the trial met the provider selection criteria or expected characteristics described in the  protocol (e.g., training, experience, qualifications). | The item was too specific for a broad guideline. Wording was adjusted to make the item broader |
| Report the number of providers available to be trained in a specific intervention, and the number of providers required to deliver the specific intervention. | Report whether the planned number of intervention providers to deliver the intervention within the trial was achieved. | Simplified the wording and aligned it with the structure of other items for consistency |
| Report moderators of intervention fidelity. | When applicable, describe contextual factors assessed or observed by the research team that may have influenced intervention fidelity assessment or results (e.g., intervention complexity, intervention provider caseload, time spent by providers on small talk vs time delivering the intervention as intended, etc). | Combined items related to contextual factors and provided greater clarity and examples |
| Specify how the intervention was piloted, and how the findings from the piloting process informed the fidelity assessments. | Describe any revisions to intervention fidelity assessments based on findings from the intervention development process (e.g., piloting the intervention). | Not only piloting but also other revisions could be important to report. Wording was adjusted to make the item broader |
| Specify whether the fidelity measures/tools were developed during the study or adapted from existing ones. | Describe how intervention fidelity measures/tools were developed. If  existing measures/tools are used, specify them and provide their source. | Used ‘how’ to reflect both quality and quantity |
| Specify when fidelity assessments were conducted, such as concurrently with the intervention or after the intervention period was completed. | Specify when intervention fidelity data was analysed, such as concurrently with the intervention or after the intervention period was completed. | Clarified that this item relates to when the data were analysed |
| Specify if the fidelity sample represents different stages of the intervention (e.g., early, middle, or late) and whether fidelity assessments covered full sessions or only specific segments. | Specify the timepoints of the intervention where intervention fidelity data was collected (e.g., week 1, 6, and 12). | Simplified the item and made it broader |
| Describe the rationale for any sensitivity analysis of fidelity data (e.g., intention-to-treat versus per protocol analysis, removing low fidelity cases). | If applicable, describe the rationale for any sensitivity/additional analysis informed by intervention fidelity data (e.g., intention-to-treat versus removing low fidelity cases, mediation analysis based on fidelity to specific intervention components). | Clarified the examples and added ‘if applicable’ |
| Specify how the sample size for fidelity assessment was determined (e.g., all sessions, random subset of sessions). If the fidelity sample was randomly selected, explain the randomisation process. | Specify what is the sample size of the intervention fidelity assessments and how it was determined (e.g., whole trial sample, subset sample). | Simplified the item and examples |
| Describe any actions taken to minimise selection bias when selecting sessions to be rated and how the fidelity sample is representative of the whole trial sample. | Describe any actions taken to minimise selection bias when sampling a sub-set of data points for intervention fidelity analysis. | Simplified the item and made it broader |
