## Appendix 3 for "Establishing consensus on domains and items for Reporting intervention Fidelity in Non-Drug, non-surgical trials: the ReFiND Delphi study"

**ReFiND Consent**

### Welcome to the ReFiND Delphi Study Round 3

Dear ReFiND Collaborators,

Thank you for completing the previous rounds of the ReFiND Delphi survey. We appreciate your time and contributions.

Below, you will find important clarifications and guidance for our final round, Round 3. Please review this information carefully before proceeding with the survey (tick the boxes once you have read the information).

**From Round 2 to Round 3**

In Round 2, four out of seven domains met the consensus threshold for inclusion in ReFiND (>70% of panellists rating 7–9). Two out of 72 items met the threshold for exclusion (>70% of panellists rating 1–3), while 13 items met the threshold for inclusion (>70% of panellists rating 7–9). These domains (n=4) and items (n=15) that achieved consensus for either inclusion or exclusion were removed from the Delphi survey and will not be subject to voting in Round 3.

Therefore, the Round 3 survey includes three domains and 57 items that did not achieve consensus in the previous rounds. A summary of ratings and comments from Round 2, providing the rationale for the ratings, is presented alongside the domains and items in Round 3.

Comments suggesting changes to a domain or item (e.g., rewording, merging, etc.) or clarifications about the survey in general were thoughtfully reviewed and carefully considered during our analysis and are not displayed in the survey. Most suggestions were addressed; however, not all could be incorporated due to occasional differences in the feedback. For example, in Round 1, some panellists recommended splitting items with multiple embedded components, while in Round 2, others suggested merging similar items. Additionally, some panellists suggested adding content to an item, but the proposed content overlapped with information already included in other items. While not every suggestion could be implemented, all feedback is highly valued and will play an important role in shaping the consensus meeting and informing the interpretation of our findings.

Every domain and item in the Delphi process must be voted on at least twice to achieve consenus, ensuring panellists take feedback from previous round into account when providing ratings. Domains and items introduced in Round 2 (not voted on in Round 1) or extensively revised between Rounds 1 and 2 are considered new and will be subject to voting in Round 3. For this reason, you may notice that a few items in this survey already met the inclusion criteria (>70% of panellists rating 7–9) in Round 2 but are included again for voting in Round 3.

☐ **I have read the information above.**

**Rating process in Round 3**

The rating process in Round 3 is identical to that in previous rounds. You will rate the importance of domains and items for inclusion in the reporting guideline using a 9-point scale (1 = not important, 9 = very important). As in previous rounds, our goal is to develop a **CORE SET** of reporting recommendations that includes only the **MINIMUM** and **ESSENTIAL** domains and items for intervention fidelity reporting in non-drug, non-surgical trials.

As previously mentioned, 13 essential items have already been included in ReFiND based on voting in earlier rounds. Therefore, please ensure that you assign 7–9 ratings only to items you believe are essential and should be included in this core set.

Items that do not achieve consensus in Round 3 will not be included in ReFiND. However, we have received suggestions to include an optional set of items alongside the ReFiND core set. Therefore, items rated as important but not essential in Round 3 may be discussed in the consensus meeting following Round 3 to determine whether they should be reported as optional items outside the core set (as a separate set).

☐ **I have read the information above.**

**Scope**

As highlighted in Rounds 1 and 2, the original recommendations from the literature, from which domains and items were extracted, were rarely developed using consensus methods and often blend general trial or intervention reporting with intervention fidelity. As a result, some survey items may appear broader than intervention fidelity. We included these items in the survey to ensure the decision—whether to include or exclude them—is based on your ratings (i.e., international consensus), not the judgment of a select group of authors.

If you believe an item is more related to general trial or intervention reporting rather than intervention fidelity, please vote to exclude it, as it falls outside the scope of ReFiND.

☐ **I have read the information above.**

**Relationship between ReFiND and other reporting guidelines**

ReFiND will serve as a standalone reporting guideline for intervention fidelity. We will encourage researchers to use CONSORT for general trial reporting, TIDieR for general intervention reporting, and ReFiND for intervention fidelity reporting. TIDieR authors suggest using TIDieR alongside CONSORT for general intervention reporting items, and we will similarly recommend that researchers using TIDieR refer to ReFiND for intervention fidelity reporting items.

While ReFiND focuses on intervention fidelity in trial reporting, the agreed domains and items may also apply to intervention fidelity in protocol reporting. During the consensus meeting, we will discuss their relevance for protocols, and if consensus is reached, we will encourage their incorporation into protocol reporting (specifically for intervention fidelity), alongside SPIRIT. For now, please rate the domains and items based on their relevance for intervention fidelity reporting, not on whether they are published in the trial results paper (what happened) or in in the trial protocol paper (what was intended to happen).

☐ **I have read the information above.**

**Applicability to different trials and interventions**

We understand that it is nearly impossible for an item to be applicable to all types of trials and non-drug, non-surgical interventions. However, some items will be more relevant than others. We ask you to use your judgment to decide whether certain domains and items should be included based on their perceived applicability. For example, some items about intervention providers may not apply to self-management digital interventions. However, considering the broader range of interventions, you may still find it worthwhile to include those items, as they may be applicable to many other types of intervention. As with other reporting guidelines, once we have agreed on a core set, we will encourage authors to use only those items that are applicable to their specific context and disregard those that are not.

☐ I have read the information above.

**Word/space limitations**

Some comments highlighted concerns about word and space limitations in trial reporting, which we acknowledge and appreciate. For this Delphi survey, we kindly ask that the rating of **essential** domains and items for intervention fidelity reporting focus on their importance, rather than on these word/space limitations. Recommendations on how and where to best report items deemed essential will be addressed after Round 3.

☐ I have read the information above.

#### Instructions - ROUND 3

This Round 3 survey will take approximately 35 to 65 minutes to complete, though the duration may vary. The purpose of this study is to achieve consensus on a core set of domains and items to be included in a reporting guideline for intervention fidelity. For this study, *intervention fidelity* refers to the extent to which an intervention is implemented as planned in the trial protocol.

You can pause the survey at any time by closing your browser and resume later, as long as you use the same browser where you started. Please note, however, that you may not be able to return to previous pages once you have submitted your responses.

Round 3 will remain open until **6 February 2025**. We will contact you regarding the online consensus meeting in March 2025. The success of this reporting guideline in driving positive changes in intervention fidelity reporting relies on your participation, so we kindly ask that you complete this final round by the deadline. Thank you again for your collaboration.

The Round 3 survey begins on the following page.

☐ I have read the information above.

*This study received approval from the Monash University Human Research Ethics Committee (ID 41579/2024).*

*Monash University values the privacy of every individual's personal information and is committed to the protection of that information from unauthorised use and disclosure except where permitted by law. For more information about Data Protection and Privacy at Monash University please see our Data Protection and Privacy Procedure.*

*If you have any questions about how Monash University is collecting and handling your personal information, please contact our Data Protection and Privacy Office at.*

#### SECTION 1 - Domains

##### SECTION 1 – Rating Domains

In this section, we present intervention fidelity domains with descriptions and examples and ask you to rate their importance for intervention fidelity evaluation and/or reporting. You will use a 9-point scale (1 = 'not important' to 9 = 'very important'). You can also provide comments in the optional free-text box accompanying each domain.

**Rating 1 to 3:** indicates the domain is not important for intervention fidelity evaluation and/or reporting.
**Rating 4 to 6:** indicates the domain may be important but is not essential for intervention fidelity evaluation and/or reporting.
**Rating 7 to 9:** indicates the domain is very important for intervention fidelity evaluation and/or reporting.

**Key information to consider while rating domains (Please READ CAREFULLY)**

- For this study, **intervention fidelity** refers to the extent to which an intervention is implemented as planned in the trial protocol. This is a working definition and will be updated after the Delphi study to ensure it aligns with the domains and items achieving consensus after the third survey round. Acknowledging differing perspectives on what constitutes fidelity, we **seek your expert opinion** on the importance of each domain presented.
- We aim to achieve consensus on **domains specific to intervention fidelity**. Therefore, when rating a domain, please focus on its importance to intervention fidelity evaluation and reporting, rather than its relevance to general intervention reporting.
- For this study, a **domain** refers to a specific and connected set of aspects related to intervention fidelity that should be assessed and reported in trials of non-drug, non-surgical interventions.

- Please review and consider the **summary of ratings and verbatim comments from Round 2** presented alongside each domain before providing your ratings. Note that the comments represent the justification for ratings from only some panellists, as not all panellists provided comments in Round 2.
- We recognise that intervention fidelity is a complex topic with mixed views. This Delphi process aims to identify intervention fidelity domains and items that are agreed as **essential** to most researchers in the field.
- Please note that the domain numbers in this survey align with those used in Round 2. Since some domains that reached consensus in Round 2 are not included from Round 3, the **numbering may appear inconsistent**. For example, the survey may start with Domain 4 instead of Domain 1, as the earlier domains have already achieved consensus. Kindly disregard the numbering and focus on the domain content.

Please rate the importance of the domains below for intervention fidelity evaluation and/or reporting:

**Domain 4: Intervention Receipt by Participants**
Description: This domain focuses on two concepts: how intervention participants (a) received (actual receipt) and (b) understood the intervention (understanding).

(4a) Actual receipt: It may assess the extent to which participants received the intervention as planned. For example, in a group intervention where health professionals deliver weekly lectures on diabetes management, it assesses whether participants attended lectures (i.e., received the intervention).

(4b) Understanding: It may assess whether participants understood the instructions, concepts, or content of the intervention as outlined by the intervention's principles. This may include checking whether they can accurately repeat what was taught/delivered.

ATTENTION: Based on feedback from previous rounds, this domain has two components (4a and 4b), which are rated independently. If you do NOT agree that Domain 4, 'Intervention Receipt by Participants', should be included in the reporting guideline, you should give a low score to both components (4a and 4b). If either component meets the threshold for inclusion consensus (>70% of ratings 7-9), Domain 4 will be recommended for inclusion in the guideline.

Ratings from Round 2:

| | **Median (IQR)** | % of panellists rating this item as **1-3** | % of panellists rating this item as **4-6** | % of panellists rating this item as **7-9** |
|---|---|---|---|---|
| **(4a) Actual Receipt** | 8 (7, 9) | 7.3 | 16.4 | **76.4** |
| **(4b) Understanding** | 7 (4, 8) | 20.0 | 27.3 | 52.7 |

Verbatim comments from panellists on the ratings in Round 2:

- *‘‘4a is measuring what happened whilst 4b is measuring the effect of what happened - which is arguably a trail outcome. So 4a high priority and 4b low priority’*
- *‘4b principally represents an outcome (understanding) - and might reflect a poorly-designed (rather than a poorly delivered or implemented) intervention - which then itself acts as a potential moderator of an intervention's intended outcomes’*
- *‘Relates to participant comprehension of the intervention, not the fidelity of the intervention’*
- *‘These can be very difficult to measure and therefore to report (especially 4b), particularly in complex behavioural trials. Setting and context also plays a significant role in how this can be reported. Both have been recognised as important aspects of fidelity, but 4b may more closely evaluate intervention feasibility/ acceptability/ participant buy-in, as opposed to deviation from protocol (fidelity)’*
- *‘Fidelity is about what is given - how people understand it may be a function of more than fidelity’*

- *‘Marketing literature already distinguishes reach for effective reach. The latter means that you may have received a letter (reach or you were present during the lecture), but did you open the letter and read its content (were you really listening to the lecturer). Whether the content of the lecture was understood is a design issue and is the outcome of what the lecture should have achieved. It is important to know, but this is not an element of the extent to which the intervention is implemented as planned. I have seen interventions that were too complicated for their low-educated audience. That is a design issue. Not implementation. Again the facts, did it happen vs Quality and Result’*
- *‘In my opinion, receipt is not strictly part of fidelity but is more closely related to adherence. An intervention may be delivered poorly (resulting in poor receipt), but fidelity primarily concerns whether the intervention is delivered as intended —that is, the degree of exactness with which it adheres to the original design or protocol’*
- *‘For me, 4b (understanding) relates to understanding intervention mechanisms, not so much to fidelity (if an intervention was implemented as planned)’*
- *‘In an ideal situation, with a properly developed intervention, 4b should be redundant as understanding by the intended audience should have been covered as part of the intervention development’*
- *‘No critical to evaluate and understand fidelity intervention’*
- *‘Actual receipt measures a different domain (reach) and should not be part of intervention fidelity measurement. The extent to which the intervention was adequately received by target recipients is however important and could be measured as part of intervention fidelity, although not necessarily a prerequisite of this domain’*
- *‘Participants can receive an intervention (e.g. by attending the lectures) but do not necessarily need to understand the information to enact the intervention. What matters most is that they understand the instructions provided’*
- *‘Methods of instruction vary and what is better for one participant may not hold for others’*
- *‘I feel engagement or participant responsiveness/relationship with providers is missing from this as defined above’*
- *‘I rated this domain as a 6 because, while participant receipt and understanding are valuable for evaluating some interventions (especially behavioural or educational), they are not universally essential for intervention fidelity reporting. Their importance depends on the nature of the intervention and the extent to which participant engagement directly influences fidelity’*
- *‘Understanding is important, but is difficult to measure’*

- *‘Whilst I think it is important to know whether patients understood the intervention or not, in different trial designs this will matter to a greater or lesser extent. Not understanding would be a mechanism by which interventions did not work’*
- *‘Reporting of physical receipt is important because if delivery fidelity was high and outcomes poor but only 50% of the participants were present, that may explain outcomes. Reporting of understanding is important because if delivery fidelity was high and outcomes poor, it helps point to where problems may be with the intervention. Perhaps the language is too complicated or the demands of the intervention too high. Further to physical receipt and being able verbally reflect the intervention protocol, I think receipt is about participant acceptance and intention to implement. I think intention should be captured in this domain as well’*
- *‘You don't know what you don't know or understand. And you certainly can't teach it’*
- *‘Reporting of understanding is important because if delivery fidelity was high and outcomes poor, it helps point to where problems may be with the intervention. Perhaps the language is too complicated or the demands of the intervention too high. Further to physical receipt and being able verbally reflect the intervention protocol, I think receipt is about participant acceptance and intention to implement. I think intention should be captured in this domain as well’*
- *‘The actual receipt example above is linked to adherence and intervention delivery. This would be more important than understanding’*
- *'It is a good idea to add receipt as we felt something is missing under receipt domain’*
- *‘Both are important’*
- *‘So often ignored in research, but the intervention will be ineffective if participants do not attend or understand the intervention’*
- *‘I think these are both important things to know in understanding the effectiveness of an intervention but whether 4b constitutes fidelity I'm less sure'*
- *‘This is critical - the extent to which participants attend (and receive) planned intervention components where the intervention is delivered needs to be reported’*

Please note that Concept 4a (Actual Receipt) met the inclusion criteria in Round 2 (>70% of panellists rating 7-9) but remains subject to voting in Round 3. This is because Domain 4 was significantly revised between Rounds 1 and 2 (i.e., split into two concepts) and considered new, meaning no feedback from panellists was provided in Round 2. According to Delphi methodology principles, panellists must receive feedback from the previous round to ensure informed consensus.

| | Not important | | | | | | Very important | | |
|---|---|---|---|---|---|---|---|---|---|
| | 1 | 2 | 3 | 4 | 5 | 6 | 7 | 8 | 9 |
| 4a: Actual receipt | | | | | | | | | |
| 4b: Understanding | | | | | | | | | |

Please provide any comments or suggestions you have about Domain 4.

**Domain 5: Intervention Enactment by Participants**
Description: This domain focuses on how intervention participants enacted the intervention. It may assess the extent to which participants integrate what they learned from the intervention into their routine behaviours. For example, it may assess whether intervention participants who received a lecture on preparing low-carbohydrate meals for diabetes management incorporated these preparations into their daily routines.

Ratings from Round 2:

| **Median (IQR)** | % of panellists rating this item as **1-3** | % of panellists rating this item as **4-6** | % of panellists rating this item as **7-9** |
|---|---|---|---|
| 5 (4, 7) | 23.6 | 45.5 | 30.9 |

Verbatim comments from panellists on the ratings in Round 2:

- *‘I agree with the comments suggesting that this item might not be part of fidelity. It could be more related to other factors and, in any case, would be part of the trial results’*

- *‘Agree with many comments that this is not about fidelity per se, even though in NIH BCC framework’*
- *‘Taking the comments above into account, I would consider this as a primary or secondary intervention outcome, rather than a measure of intervention fidelity’*
- *‘As 4b, this principally represents an outcome (are participants doing what the intervention seeks to get them to do?) and is affected by a whole range of factors, even if the intervention is delivered with perfect fidelity by providers, and with perfect receipt by participants’*
- *‘Enactment should be at least partially determined before the intervention is delivered in a trial ie as part of a feasibility study or other vanguard study. While for some it could be conceived as adherence within a trial, I see it as more of an outcome. Not important to fidelity’*
- *‘In my opinion, enactment is not strictly part of fidelity but is more closely related to adherence. An intervention may be delivered poorly (resulting in poor enactment), but fidelity primarily concerns whether the intervention is delivered as intended—that is, the degree of exactness with which it adheres to the original design or protocol’*
- *‘I still feel that intervention fidelity could be high whilst enactment is low suggesting that it is a somewhat separate element from fidelity’*
- *‘I rated this domain as a 6 because, based on my experience using the NIH-BCC and acceptability frameworks for evaluating a pilot study, I observed significant overlap between this construct and acceptability or implementation evaluation. Enactment aligns more closely with participants’ acceptance and integration of the intervention into their routines, which is a core aspect of acceptability and implementation outcomes, rather than fidelity. Fidelity evaluation focuses more on whether the intervention was delivered and received as planned, whereas enactment extends beyond this into behavioural adoption, which fits better under acceptability or outcome evaluation’*
- *‘Fidelity has multiple levels, including this one. It overlaps with the concept of patient adherence’*
- *‘Enactment is more an outcome rather than a measure of fidelity*

- *'I was very surprised (and more than a little bit disappointed) to see the enactment domain receive such low ratings (fewer than 2/3s of respondents rated this domain as essential)! While fidelity enactment or other aspects of fidelity are considered "implementation outcomes" this does not mean it is not a central aspect of fidelity and essential to good fidelity reporting. Admittedly there will be some trials such as hybrid implementation-effectiveness trials where fidelity may be an important outcome, but the fidelity with which an intervention is enacted remains an essential part of fidelity. A fidelity guideline that doesn't' include enactment would itself have low fidelity as it would not reflect the well regarded and simple definition of fidelity as reflecting the extent to which an intervention is implemented as intended. It is about way more than delivery, despite the fact that most trial only report on delivery'*
- *'If the intervention has elements that the target group should take notice of than fidelity is related to whether they were exposed to these elements (and subsequent steps within them). The impact is an outcome. Sure, this is informative, but it is not about implementing as planned. Information on fidelity gives us reasons why there might not be an effect, as steps taken were missing, but that is something different. Then you have to do new research to find out why the current setup was not working for your target group. That is why pilot testing is so important before you even expose people to your intervention'*
- *'The significant overlap of this domain with study outcomes and participant adherence was noted in Round 1, and is a domain weakness. Despite the complexity of measuring this domain, I agree that it can provide insights into where fidelity to the protocol may break down - is it with the provider? is it with the receipt? or is it what the participant does with the received intervention? This can provide insights into mechanisms of action and how (lack of) fidelity impacted these mechanisms/ intervention effect. However, this is only the case if authors are able to report useful, meaningful evaluations of intervention enactment. I therefore think it will be important to consider what we would expect researchers to report for this domain'*
- *'I think this may be more appropriate for longer term studies. Also, in orthopedic interventions, this may not be as important as say, aerobic exercise'*
- *'This is really important to understand the longer term impacts that an intervention has, and therefore whether the intervention had the intended effects'*

- *‘I equate enactment and adherence. An RCT is evaluating whether an intervention protocol works or how much it works. It's the "ideal world" version of the intervention. In reporting intervention fidelity, therefore, I think you need to know whether the intervention was implemented/enacted/adhered to by participants. Again, it helps you see where the strengths or breakdown points of an intervention are. If it was delivered as intended, there was understanding and intent to implement but poor implementation, researchers can then target understanding the gap between intention and implementation. If you only look at delivery fidelity, it doesn't give you the full picture between intervention protocol and outcome and potentially effective interventions may be discarded or interventions that were effective but for reasons other than enactment of the protocol itself may be accepted but unable to be reproduced by others because the reason for their effectiveness has not been identified’*
- *‘I would argue that reporting on fidelity enactment is crucial for fidelity reporting - it helps shed light on the mechanism(s) of impact. The fact that it is difficult to measure and report on fidelity enactment does not make it any less important. There are creative ways to measure and report on this key aspect of fidelity. While fidelity will be an important outcome in some trials (eg implementation trials), it is still essential to report on fidelity enactment in those trials. And in effectiveness trials, enactment is critical to measure and report in order to give meaning and enhance understanding regarding effectiveness results (eg did a trial fail because the intervention is poor or because it wasn't enacted at all or as intended)’*
- *‘While some overlap with the study outcomes are inevitable, I still believe that this should be included. This domain does not focus on the end outcome per se, but rather how the participants/patients arrived to the expected outcome and whether this aligned with the hypothesised mechanisms. Some participants might have changed (e.g. improved their diabetes management / diet plan) but not necessarily as was intended’*

- *‘Enactment is important to measure as it provides a way of understanding the mechanism through which intervention outcomes may occur. E.g. if take the example of a medicine trial as an example of a simple intervention - if we do not know whether the participants take the medicine after attending the intervention, how do we know whether the medicine works to improve their health outcomes or not (e.g. improvements in condition symptoms). This also relates to complex interventions that have multiple components and multiple asks of participants in terms of behaviour change. I note comments above that talk about this being an outcome - I agree that this is an outcome but not necessarily a study outcome. I believe in many cases the behaviour measured through enactment are more process outcomes rather than intervention outcomes. If not measured within fidelity the risk is that this wouldn't be measured, and from the fidelity measures would only be able to infer if the intervention was delivered and received, but not whether participants actually used those skills in their daily life. I agree also that it has some overlap with study outcomes, but I think an important perspective for authors of fidelity research is to specify how they are defining the fidelity outcomes (e.g. fidelity of delivery, receipt, enactment) and study outcomes that the intervention aims to change’*
- *‘This is crucial for behavioral interventions’*
- *‘I disagree that this is linked to outcomes as the outcome may not be closely aligned with the intervention and target of choice’*
- *‘Reporting enactment is essential in research trials although there are some misunderstanding between this and trial outcome’*

| | Not important | | | | | | Very important | | |
|---|---|---|---|---|---|---|---|---|---|
| | 1 | 2 | 3 | 4 | 5 | 6 | 7 | 8 | 9 |
| D5: Intervention Enactment by Participants | | | | | | | | | |

Please provide any comments or suggestions you have about Domain 5.

**Domain 6: Moderators of intervention fidelity and outcomes**

Description: This domain focuses on aspects that may moderate intervention fidelity and the relationship between intervention fidelity and trial outcomes. It may assess whether contextual elements, such as the provider-participant relationship or organisational factors (e.g., provider caseload), impact intervention fidelity. It may also assess whether intervention fidelity itself influences trial outcomes, and how the outcomes of an intervention may be interpreted in the context of its fidelity results.

Ratings from Round 2:

| **Median (IQR)** | % of panellists rating this item as **1-3** | % of panellists rating this item as **4-6** | % of panellists rating this item as **7-9** |
|---|---|---|---|
| 5 (3, 6) | 34.5 | 41.8 | 23.6 |

Verbatim comments from panellists on the ratings in Round 2:

- *‘Important, but more about understanding how interventions lead to outcomes rather than reporting of fidelity itself’*
- *‘Agreed that it is an important construct in evaluating an intervention's efficacy or effectiveness, but not the fidelity with which it was implemented as intended’*
- *‘This is important, but it is difficult to tease apart what is fidelity and what is an outcome. For example, many interventions are changing the healthcare setting and this would be an outcome’*
- *‘More like a secondary analysis of treatment effects. Not important’*
- *‘Although context is critical to assessing fidelity, and perhaps could form a part of both "receipt" and "delivery" domains, using the term moderators may be too complex/unclear. Intervention effect moderators in and of themselves are distinct from fidelity’*
- *‘Knowing what 'external factors' impact fidelity is important to report eg contexts, provider issues etc. Not sure the term moderators is the right one here (moderators are variables that cannot, or are unlikely, to be amenable to change..)’*
- *‘I agree that the concept of moderators may not be clear’*

- *‘I think author reflections on the moderators or potential moderators of intervention fidelity on outcomes is an important aspect of fidelity reporting. I don't think they can be measured necessarily, they are not well defined. However, that is the point. This is the "real world" aspect of an intervention and author insights can provide valuable context and understanding. It should be included in fidelity reporting’*
- *‘I think moderators are very important to address but I think it's not clear enough yet on what they are’*
- *‘I am not sure if this item is directly related to fidelity. Additionally, moderators can be numerous and not all of them fully known’*
- *‘Many moderating factors, such as caseload or organisational culture, are external to the intervention itself and may be challenging to account for systematically. These factors are better addressed in implementation or contextual analyses rather than fidelity evaluation’*
- *‘Needs to measured to ensure barriers are mitigated for equitable implementation but not part of fidelity’*
- *‘As stated, you have quantity, did it happen, and quality. You may want to know both. Quality is related to the design of the study. Even if providers do not do what you want them to do or similar for patients, you were not able to design an intervention that can work for them. as the item already states moderators of fidelity. So it is not about fidelity itself. But these are thing you would like to investigate to know what can be improved. But, that is a different question’*
- *‘Although important, on second thought I think this should not be part of standard intervention fidelity measurement. It speaks to the underlying assumptions of how a program is anticipated to work and would typically be part of e.g., a realist evaluation of how a program works, for whom and under which circumstances - but this is another aspect of evaluation all together and should not be part of routine intervention fidelity measurement’*
- *‘Relevant to large trials with a large number of providers/participants’*
- *‘Critical to understand deviation’*
- *‘Important to consider as it will inform how scalable the intervention is in the real world’*
- *‘Likely to be underpowered’*
- *‘If purely report fidelity then I'd say this is less important but if about how to understand fidelity and impact of fidelity then this becomes more important’*
- *‘Less important- we can consider these as sub-factors of main fidelity domains’*

- *‘Gathering data on intervention fidelity is already labour intensive and tricky. It is good to mention or to consider moderators and an author may describe these in a little detail but to make it an essential criteria is nor sufficient’*
- *‘I doubt this will be picked up during the assessment of fidelity - It is more likely to be identified at the end of the main trial’*

| | Not important | | | | | | | Very important | |
|---|---|---|---|---|---|---|---|---|---|
| | 1 | 2 | 3 | 4 | 5 | 6 | 7 | 8 | 9 |
| D6: Moderators of intervention fidelity and outcomes | | | | | | | | | |

Please provide any comments or suggestions you have about Domain 6.

If you any further comments or suggestions about the domains listed in this survey, please enter them here.

#### SECTION 2 - Instructions - items

##### SECTION 2 – Rating Individual Items

In this section, we present intervention fidelity items and ask you to **please rate their importance for inclusion in the reporting guideline for intervention fidelity**. You will use a 9-point scale (1 = ‘not important’ to 9 = ‘very important’). You can also provide comments in the optional free-text box accompanying each domain.

**Rating 1 to 3:** indicates the item is not important for inclusion in the reporting guideline.
**Rating 4 to 6:** indicates the item may be important but is not essential for inclusion in the reporting guideline.
**Rating 7 to 9:** indicates the item is very important for inclusion in the reporting guideline.

**Key information to consider while rating items (Please READ CAREFULLY)**

- For this study, **intervention fidelity** refers to the extent to which an intervention is implemented as planned in the trial protocol. This is a working definition and will be updated after the Delphi study to ensure it aligns with the items and domains achieving consensus after the third survey round. Acknowledging differing perspectives on what constitutes intervention fidelity, we will present items that vary in use in the literature and ask for your expert opinion on their importance for inclusion in the reporting guideline.
- For this study, a **reporting guideline** is a set of recommendations on essential information that should be included in academic papers, facilitating understanding, replication, usability, and further research. The reporting guideline is overarching, meaning that the items should be broad enough to be applicable to nearly every type of non-drug, non-surgical intervention and research trial.
- The items in this survey were identified from several intervention fidelity guidance documents, each representing different perspectives. You may find that some items are not directly related to intervention fidelity. Therefore, when rating an item, please remember that we are seeking consensus on those that are **specific to intervention fidelity** reporting, rather than general intervention reporting.
- Items in this survey are displayed in a **RANDOM ORDER** to reduce the effects of response fatigue. This is intentional, so please do not worry if things seem out of sequence.
- Please note that the item numbers in this survey align with those used in Round 2. Since some items that reached consensus in Round 2 are not included in Round 3, the **numbering may appear inconsistent**. For example, you may see item number 71 in the survey, but this survey includes only 57 items in total. Kindly disregard the numbering and focus on the item content.
- Our goal is to develop a **CORE SET** of reporting recommendations that includes only the **MINIMUM** and **ESSENTIAL** items for intervention fidelity reporting in non-drug, non-surgical trials. As previously mentioned, 13 essential items have already been included in ReFiND based on voting in earlier rounds. Therefore, please ensure that you assign 7–9 ratings only to items you believe are essential and should be included in this core set.
- Please review and consider the **summary of ratings and verbatim comments** from Round 2 presented alongside each item before providing your ratings. Note that the comments represent the justification for ratings from only some panellists, as not all panellists provided comments in Round 2.

#### ITEM 1-8

Please rate the importance of including the items below in a reporting guideline for intervention fidelity in non-drug, non-surgical trials.

Note: Some items are closely worded but their main substance is unique (highlighted in **bold**). Comments from Round 2 are reported verbatim, with some repeated across items as provided by the panellists.

**ITEM 5:** Report the degree to which the intervention delivered to intervention participants is **congruent with its underlying theory or logic model**.

Ratings from Round 2:

| **Median (IQR)** | % of panellists rating this item as **1-3** | % of panellists rating this item as **4-6** | % of panellists rating this item as **7-9** |
|---|---|---|---|
| 7 (5, 8) | 18.2 | 30.9 | 50.9 |

Verbatim comments from Round 2:

- ‘*While this seems to be a mandatory item for improving the replication and outcome evaluation, this is a not a must have for a fidelity reporting as long as the intervention components are reported in detail’*
- *‘Congruence to underlying theory is about the intervention development more than about fidelity’*
- *‘Surely this is part of the intervention development process & should be reported in the intervention development paper’*
- *‘Often this is described as process evaluation. Unravelling the black box of change. The fidelity of the theory. Indirectly this is about the fidelity of the intervention as well. Important, but a bit different from assessing of what happened.’*
- *‘Helpful but not exactly intervention fidelity reporting. Intervention should be clearly defined and reported. Underlying theory important but not essential as previously discussed’*
- *‘Not sure about this... this would be a limitation (if things didnt work) or reported in a paper or report anyway. Is it necessary to stipulate it?’*
- *‘This is fidelity in intervention design, not implementation’*
- *‘Not sure this belongs into a fidelity reporting guideline, although very important’*

- *‘In specific physiotherapy interventions, it can be challenging to clearly identify the underlying theory or logic model’*
- *‘Not as important’*
- *‘Agree that cannot assess fidelity without this’*
- *‘Essential’*
- *‘If intervention is based on theory then this is important’*
- *‘Fidelity to the logic mode/underlying theory is surely the definition of fidelity? Otherwise what are we assessing delivery against?’*
- *‘This is important to assess whether the intervention retains its fidelity to the theory underlying the intervention (are we still testing the underlying theory we think we're testing?) which is an important part of fidelity’*
- *‘These need to be congruent even if only reporting to a prespecified number of components or the main component of an underlying theory’*
- *‘This is all too often overlooked and should be part of the core recommendations for fidelity reporting’*

| | Not important | | | | | Very important | | | |
|---|---|---|---|---|---|---|---|---|---|
| | 1 | 2 | 3 | 4 | 5 | 6 | 7 | 8 | 9 |
| ITEM 5 | | | | | | | | | |

Please provide any comments or suggestions you have about ITEM 5.

**ITEM 6:** Describe a **method to assess** whether unintended components are delivered (e.g., components not planned or explicitly specified to be avoided in the original design).

Ratings from Round 2:

| **Median (IQR)** | % of panellists rating this item as **1-3** | % of panellists rating this item as **4-6** | % of panellists rating this item as **7-9** |
|---|---|---|---|
| 6 (5, 8) | 12.7 | 43.6 | 43.6 |

Verbatim comments from Round 2:

- *‘Nice to have rather than essential - to me fidelity is more about whether the planned components were delivered’*
- *‘Unintended effects can be positive or negative, This you already assess or are supposed to assess in a study. Is not a core fidelity issue’*
- *‘If i have 4000 words I'd rather write about the good stuff’*
- *‘Could be added in the comment to other item for researchers to think about whether to report this’*
- *‘Part of the development of the fidelity measure. This is useful to report, but could be addressed by including a copy of the fidelity measure(s) as Supplemental Materials’*
- *‘I wonder if this is more about reporting adaptations’*
- *‘Assessing unintended components may align more closely with understanding intervention outcomes than with core fidelity evaluation. It helps contextualise results rather than directly evaluating fidelity’*
- *‘Useful’*
- *‘These unintended components could undermine the intervention impact’*
- *‘Off-protocol events are important consideration’*
- *‘Feels essential to try and capture this (but hard!)’*
- *‘Important part of delivery of the intervention (by providers) - for example, if providers have personal opinions against an aspect of an intervention and give participants reasons to avoid carrying it out, this undermines intervention fidelity’*
- *‘Important and known factors that might influence the outcome as co-interventions should be described and compared between groups. Unnecessary components could be hard to be defined a priori’*
- *‘Fidelity goes both ways, include what should be there & do not include that which should not be there’*

| | Not important | | | | | Very important | | | |
|---|---|---|---|---|---|---|---|---|---|
| | 1 | 2 | 3 | 4 | 5 | 6 | 7 | 8 | 9 |
| ITEM 6 | | | | | | | | | |

Please provide any comments or suggestions you have about ITEM 6.

**ITEM 7: Report whether** unintended components (e.g., components not planned or explicitly specified to be avoided in the original design) were delivered.

Ratings from Round 2:

| **Median (IQR)** | % of panellists rating this item as **1-3** | % of panellists rating this item as **4-6** | % of panellists rating this item as **7-9** |
|---|---|---|---|
| 7 (5, 8) | 14.5 | 32.7 | 52.7 |

Verbatim comments from Round 2:

- *'I rated this item as a 5 because, while reporting unintended components can provide useful context for understanding trial outcomes, it is not essential for core fidelity evaluation. Its importance depends on the complexity and nature of the intervention. This item may be particularly relevant for a cRCT, or similar designs with multiple providers or settings'*
- *'It is an essential element for any study. This is about research ethics'*
- *'Critical when there might have been a very "active" component for the behavior change that was delivered but was not formally part of the intervention package'*
- *'Important but not critical'*
- *'The judgement of helpful or not is irrelevant and is ultimately based on participant response'*
- *'Unless these someone undermine what was supposed to be delivered in the intervention, this is not essential'*
- *'Nice to have rather than essential - to me fidelity is more about whether the planned components were delivered'*
- *'Could be interesting but not critical - to understand possible lack of fidelity'*
- *'Important part of delivery of the intervention (by providers) - for example, if providers have personal opinions against an aspect of an intervention and give participants reasons to avoid carrying it out, this undermines intervention fidelity'*

- *‘Yes, I think this is part of "delivered as intended". Has implications for fidelity, replication and outcome’*
- *‘Include extra helpful components that were not intended’*
- *‘Very essential although it was very rarely followed in existing studies’*
- *‘Fidelity goes both ways, include what should be there & do not include that which should not be there’*

| | Not important | | | | | Very important | | | |
|---|---|---|---|---|---|---|---|---|---|
| | 1 | 2 | 3 | 4 | 5 | 6 | 7 | 8 | 9 |
| ITEM 7 | | | | | | | | | |

Please provide any comments or suggestions you have about ITEM 7.

**ITEM 8:** Specify how delivery contamination **between intervention arms** was avoided.

Ratings from Round 2:

| **Median (IQR)** | % of panellists rating this item as **1-3** | % of panellists rating this item as **4-6** | % of panellists rating this item as **7-9** |
|---|---|---|---|
| 5 (3, 7) | 41.8 | 29.1 | 29.1 |

Verbatim comments from Round 2:

- *‘Critical for trial design, not necessarily a component of fidelity’*
- *‘Important construct but not part of fidelity assessment’*
- *‘This does not belong to a fidelity reporting guideline’*
- *‘Agree with comments that this is more trial design. The fidelity reporting part would be whether contamination occurred or not, based on evaluation of which components were delivered’*
- *‘By reporting fidelity, this should be self-evident and does not need to be reported separately’*
- *‘Important for replying intervention but not essential to fidelity assessment’*

- *‘The concept that you need to know exactly what was delivered to each intervention arm is clearly important but whether they need to specify how avoided contamination I'm unsure. The importance of this item will be dependent on the other items present’*

| | Not important | | | | | Very important | | | |
|---|---|---|---|---|---|---|---|---|---|
| | 1 | 2 | 3 | 4 | 5 | 6 | 7 | 8 | 9 |
| ITEM 8 | | | | | | | | | |

Please provide any comments or suggestions you have about ITEM 8.

**ITEM 9-18**

Please rate the importance of including the items below in a reporting guideline for intervention fidelity in non-drug, non-surgical trials.

Note: Some items are closely worded but their main substance is unique (highlighted in **bold**). Comments from Round 2 are reported verbatim, with some repeated across items as provided by the panellists.

**ITEM 9: Describe a method** to assess the degree to which participants **received** the intervention components as planned (e.g., being exposed to the intervention).

Ratings from Round 2:

| **Median (IQR)** | % of panellists rating this item as **1-3** | % of panellists rating this item as **4-6** | % of panellists rating this item as **7-9** |
|---|---|---|---|
| 8 (6, 8) | 12.7 | 21.8 | 65.5 |

Verbatim comments from Round 2:

- *‘Intervention receipt is not intervention fidelity’*

- *'It provides essential insights into whether the intervention reached its intended audience. But not same as intervention receipt or enactment'*
- *'Isn't this more related to adherence rather than fidelity?'*
- *'This is not fidelity, but rather reach? Similar comments up to item 18: they all don't focus on fidelity but other aspects of the trial that are important, but do not belong in this guideline'*
- *'Whether they received what they should have is more important than exactly how it is measured'*
- *'The method is less important than the actual extent to which they received the intervention components (regarding reporting fidelity-- the method should just be part of the method of the trial)'*
- *'This is really core'*
- *'This is the core of fidelity assessment'*
- *'Yes, until there is a standardised measure'*
- *'Essential element of fidelity'*
- *'This aspect is critical for evaluating fidelity in non-pharmacological interventions, specifically in physiotherapy'*
- *'I think this is fidelity'*

| | Not important | | | | | Very important | | | |
|---|---|---|---|---|---|---|---|---|---|
| | 1 | 2 | 3 | 4 | 5 | 6 | 7 | 8 | 9 |
| ITEM 9 | | | | | | | | | |

Please provide any comments or suggestions you have about ITEM 9.

**ITEM 10: Report** the degree to which participants **received** the planned intervention **content/components**.

Ratings from Round 2:

| **Median (IQR)** | % of panellists rating this item as **1-3** | % of panellists rating this item as **4-6** | % of panellists rating this item as **7-9** |
|---|---|---|---|

| | | | |
|---|---|---|---|
| 8 (7, 9) | 10.9 | 10.9 | **78.2** |

Verbatim comments from Round 2:

- *'It provides essential insights into whether the intervention reached its intended audience. But not same as intervention receipt or enactment'*
- *'Important for an intention to treat and per-protocol analyses, but less about fidelity'*
- *'If this is the inverse to whether it was delivered as planned, then it is important'*
- *'This is really core'*
- *'This could be a one line reporting standard for fidelity assessment'*
- *'This item is also important in interventions that require the active and continuous participation of the patient'*

| | Not important | | | | | Very important | | | |
|---|---|---|---|---|---|---|---|---|---|
| | 1 | 2 | 3 | 4 | 5 | 6 | 7 | 8 | 9 |
| ITEM 10 | | | | | | | | | |

Please provide any comments or suggestions you have about ITEM 10.

**ITEM 11: Report** the degree to which participants **received** the planned intervention **dose**.

Ratings from Round 2:

| **Median (IQR)** | % of panellists rating this item as **1-3** | % of panellists rating this item as **4-6** | % of panellists rating this item as **7-9** |
|---|---|---|---|
| 8 (7, 9) | 10.9 | 10.9 | **78.2** |

Verbatim comments from Round 2:

- *'It provides essential insights into whether the intervention reached its intended audience. But not same as intervention receipt or enactment'*

- *‘Fidelity is more about delivery than receipt of intervention’*
- *‘If this is the inverse to whether it was delivered as planned, then it is important’*
- *‘Sometimes relevant, sometimes "dose" is difficult concept in non-drug studies’*
- *‘More important in some studies than others (i.e., those with a single dose)’*
- *‘This is really core’*
- *‘Dosage is definitely important alongside content and/or components’*
- *‘Yes helpful to distinguish content and dose - you probably have it later but this implies a need to report minimal dose and content received for effectiveness too’*

| | Not important | | | | | Very important | | | |
|---|---|---|---|---|---|---|---|---|---|
| | 1 | 2 | 3 | 4 | 5 | 6 | 7 | 8 | 9 |
| ITEM 11 | | | | | | | | | |

Please provide any comments or suggestions you have about ITEM 11.

**ITEM 12: Describe a method** to assess the degree to which participants **understood/comprehended** the intervention components as planned.

Ratings from Round 2:

| **Median (IQR)** | % of panellists rating this item as **1-3** | % of panellists rating this item as **4-6** | % of panellists rating this item as **7-9** |
|---|---|---|---|
| 5 (3, 7) | 40.0 | 23.6 | 36.4 |

Verbatim comments from Round 2:

- *‘Not about fidelity, more about participant health literacy’*
- *‘This is the quality of the reception’*
- *‘Outcome’*
- *‘Part of feasibility/acceptability testing’*

- *‘Participant understanding/comprehension is part of measuring if the intervention worked, not measuring if the intervention was delivered as planned’*
- *‘I think if the education is well done AND there are boosters, this is not needed’*
- *‘I think this is important, but is very difficult to measure in reality’*
- *‘Difficult to ascertain in many circumstances’*
- *‘May be difficult to meaningfully measure’*
- *‘Depends on the intervention and the stage of the trial’*
- *‘This aspect is critical for evaluating fidelity in interventions where active patient participation is required’*

| | Not important | | | | | Very important | | | |
|---|---|---|---|---|---|---|---|---|---|
| | 1 | 2 | 3 | 4 | 5 | 6 | 7 | 8 | 9 |
| ITEM 12 | | | | | | | | | |

Please provide any comments or suggestions you have about ITEM 12.

**ITEM 13: Report** the degree to which participants **understood/comprehended** the intervention components as planned.

Ratings from Round 2:

| **Median (IQR)** | % of panellists rating this item as **1-3** | % of panellists rating this item as **4-6** | % of panellists rating this item as **7-9** |
|---|---|---|---|
| 5 (3, 7) | 34.5 | 27.3 | 38.2 |

Verbatim comments from Round 2:

- *‘Not about fidelity, more about participant health literacy’*
- *‘This is not a direct measure of fidelity’*
- *‘Outcome’*

- *‘Participant understanding/comprehension is part of measuring if the intervention worked, not measuring if the intervention was delivered as planned’*
- *‘More important to report the degree (rather than method)’*
- *‘That is why you need to pilot test your intervention. It is important but should be checked before the trial. Not after’*
- *‘Important in feasibility / acceptability testing but not core in fidelity reporting’*
- *‘I think this is important, but is very difficult to measure in reality’*

| | Not important | | | | | Very important | | | |
|---|---|---|---|---|---|---|---|---|---|
| | 1 | 2 | 3 | 4 | 5 | 6 | 7 | 8 | 9 |
| ITEM 13 | | | | | | | | | |

Please provide any comments or suggestions you have about ITEM 13.

**ITEM 14: Describe a method** to assess the degree to which intervention participants **apply the intervention in real-life settings** (also referred to as 'enactment’).

Ratings from Round 2:

| **Median (IQR)** | % of panellists rating this item as **1-3** | % of panellists rating this item as **4-6** | % of panellists rating this item as **7-9** |
|---|---|---|---|
| 4 (2, 6.5) | 47.3 | 27.3 | 25.5 |

Verbatim comments from Round 2:

- *‘Enactment doesn't feel part of fidelity to me, despite what NIH BCC says’*
- *‘Outcome not fidelity’*
- *‘This is an outcome not a measure of fidelity’*
- *‘This is real life application, not fidelity of the intervention within the trial’*
- *‘I see these as a component of the intervention (e.g., was the assigned intervention homework completed, if any)’*

- *‘This overlaps a lot with intervention outcomes and adherence’*
- *‘Not a component of treatment fidelity’*
- *‘I think enactment is a component of fidelity, but is again very difficult to measure objectively’*
- *‘Relevant for some behavioral interventions’*
- *‘Absolutely crucial to report on this essential aspect of fidelity’*
- *‘Method to assess enactment needs to be standardised. In the meantime report method used for assessment’*
- *‘Apply means finding out what they do and you need to know what happened’*
- *‘It looks like from the above quotes that they did not understand the definition of enactment as described in Belg, Borrelli or Borrelli et al. it is important to ascertain that the person not only has received the intervention, understands it, but is also physically and mentally capable of performing the intervention. Otherwise, non-adherence could be misconstrued as an ineffective intervention, when in effect, the person couldn't perform it’*
- *‘Essential’*

| | Not important | | | | | Very important | | | |
|---|---|---|---|---|---|---|---|---|---|
| | 1 | 2 | 3 | 4 | 5 | 6 | 7 | 8 | 9 |
| ITEM 14 | | | | | | | | | |

Please provide any comments or suggestions you have about ITEM 14.

**ITEM 15: Report** how well intervention participants applied the intervention **content**, meaning the degree to which they **used it in real-life settings.**

Ratings from Round 2:

| **Median (IQR)** | % of panellists rating this item as **1-3** | % of panellists rating this item as **4-6** | % of panellists rating this item as **7-9** |
|---|---|---|---|
| 3 (2, 6) | 50.9 | 29.1 | 20.0 |

Verbatim comments from Round 2:

- *‘An outcome, not IF’*
- *‘Study outcomes - translating into practice’*
- *‘Important as part of outcomes evaluation, not fidelity reporting’*
- *‘Enactment is an outcome not a measure of fidelity’*
- *‘May be critical for some kinds of studies (i.e., those in which enactment is critical to participants experiencing the IV). However, probably better described as "adherence"-- depends on whether you are lumping fidelity and adherence into a similar/same construct’*
- *‘This overlaps a lot with intervention outcomes/effect; while it can provide information about what was done by participants with the intervention received, I am not fully convinced that this always a described component of a "planned protocol", especially in a pragmatic trial, and therefore may not be fidelity. This is a larger question of whether enactment should be considered part of fidelity as suggested by some current guidelines, and, if so, how it can be measured in a meaningful way that is separate from intervention effect’*
- *‘I think this moves fidelity too much away from what it really is, and in other fidelity studies we have reviewed, enactment is rarely done and when it is, it speaks more to the feasibility of the intervention’*
- *‘Often call uptake in practice acording to Rogers’*
- *‘No, fidelity is about the experimental situation’*
- *‘I think it is to different thing. Fidelity is one thing and uptake is another form of study’*

| | Not important | | | | | Very important | | | |
|---|---|---|---|---|---|---|---|---|---|
| | 1 | 2 | 3 | 4 | 5 | 6 | 7 | 8 | 9 |
| ITEM 15 | | | | | | | | | |

Please provide any comments or suggestions you have about ITEM 15.

**ITEM 16: Report** how well intervention participants **applied** the intervention **dose**, meaning the degree to which they **used it in real-life settings**.

Ratings from Round 2:

| **Median (IQR)** | % of panellists rating this item as **1-3** | % of panellists rating this item as **4-6** | % of panellists rating this item as **7-9** |
|---|---|---|---|
| 2 (2, 5) | 61.8 | 29.1 | 9.1 |

Verbatim comments from Round 2:

- *'An outcome, not IF'*
- *'Important as part of outcomes evaluation, not fidelity reporting'*
- *'Enactment is an outcome not a measure of fidelity'*
- *'May be critical for some kinds of studies (i.e., those in which enactment is critical to participants experiencing the IV). However, probably better described as "adherence"-- depends on whether you are lumping fidelity and adherence into a similar/same construct'*
- *'Not fidelity'*
- *'Too specific and too hard to measure'*
- *'Have all the enactment ones as optional for reporting'*
- *'Enactment having a dose feels a little 'meta' - this is one I could see dropping as not critical'*

| | Not important | | | | | Very important | | |
|---|---|---|---|---|---|---|---|---|
| | 1 | 2 | 3 | 4 | 5 | 6 | 7 | 8 | 9 |
| ITEM 16 | | | | | | | | | |

Please provide any comments or suggestions you have about ITEM 16.

**ITEM 17: Describe a method to assess participant adherence** to the intervention (i.e., degree to which the participant **follows the intervention as instructed/required**).

Ratings from Round 2:

| Median (IQR) | % of panellists rating this item as **1-3** | % of panellists rating this item as **4-6** | % of panellists rating this item as **7-9** |
|---|---|---|---|
| 5 (2, 7) | 45.5 | 16.4 | 38.2 |

Verbatim comments from Round 2:

- *'An outcome, not IF'*
- *'Intervention adherence is a different concept to intervention fidelity'*
- *'This item and the next one are outcomes'*
- *'Important as part of outcomes evaluation, not fidelity reporting'*
- *'This part of understanding why intervention did / did not work. Not part of assessment of its delivery'*
- *'Method is part of method, not fidelity reporting'*
- *'How can you not report this'*
- *'I am on the fence about whether adherence is a part of fidelity; It could be considered part of intervention receipt (did participants attend sessions, etc)'*
- *'This will need to be clearer and better differentiated from items 14-15 if included; adherence and fidelity enactment are not well differentiated as constructs yet in the literature (I find enactment a clearer construct)'*
- *'With this definition of adherence, think this overlaps with receipt and enactment. Adherence is a tricky concept to use as could refer to lots of different things, so i personally prefer enactment under the umbrella of engagement (to capture things like attendance, receipt, engagement) - therefore have rated this one low'*
- *'Important concept for many interventions'*
- *'Yes, in the study itself'*

| | Not important | | | | | Very important | | | |
|---|---|---|---|---|---|---|---|---|---|
| | 1 | 2 | 3 | 4 | 5 | 6 | 7 | 8 | 9 |
| ITEM 17 | | | | | | | | | |

Please provide any comments or suggestions you have about ITEM 17.

**ITEM 18: Report participant adherence** to the intervention (i.e., degree to which the participant **follows the intervention as instructed/required**).

Ratings from Round 2:

| **Median (IQR)** | % of panellists rating this item as **1-3** | % of panellists rating this item as **4-6** | % of panellists rating this item as **7-9** |
|---|---|---|---|
| 6 (2, 7) | 41.8 | 12.7 | 45.5 |

Verbatim comments from Round 2:

- *‘An outcome, not IF’*
- *‘Intervention adherence is a different concept to intervention fidelity’*
- *‘This item and the next one are outcomes’*
- *‘Important as part of outcomes evaluation, not fidelity reporting’*
- *‘This part of understanding why intervention did / did not work. Not part of assessment of its delivery’*
- *‘Not fidelity’*
- *‘How can you not report this’*
- *‘I am on the fence about whether adherence is a part of fidelity; It could be considered part of intervention receipt (did participants attend sessions, etc)’*
- *‘This will need to be clearer and better differentiated from items 14-15 if included; adherence and fidelity enactment are not well differentiated as constructs yet in the literature (I find enactment a clearer construct)’*
- *‘With this definition of adherence, think this overlaps with receipt and enactment. Adherence is a tricky concept to use as could refer to lots of different things, so i personally prefer enactment under the umbrella of engagement (to capture things like attendance, receipt, engagement) - therefore have rated this one low’*
- *‘Adherence and enactment seem intertwined to me (differ based on study, but similar foundational construct)’*
- *‘This is conceptually nice but I wonder how much studies can really talk about adherence with psychosocial interventions. They might come to x studies but whether they engage fully in exercises is a different matter’*
- *‘Relevant to many non-drug interventions’*

| | Not important | | | | | Very important | | | |
|---|---|---|---|---|---|---|---|---|---|
| | 1 | 2 | 3 | 4 | 5 | 6 | 7 | 8 | 9 |
| ITEM 18 | | | | | | | | | |

Please provide any comments or suggestions you have about ITEM 18.

**ITEM 19-22**

Please rate the importance of including the items below in a reporting guideline for intervention fidelity in non-drug, non-surgical trials.

Note: Some items are closely worded but their main substance is unique (highlighted in **bold**). Comments from Round 2 are reported verbatim, with some repeated across items as provided by the panellists.

**ITEM 19:** Describe a **method to assess** the extent to which intervention participants **accept** the intervention (e.g., their perceptions, beliefs, attitudes, and intentions to use the intervention).

Ratings from Round 2:

| **Median (IQR)** | % of panellists rating this item as **1-3** | % of panellists rating this item as **4-6** | % of panellists rating this item as **7-9** |
|---|---|---|---|
| 3 (2, 5) | 56.4 | 23.6 | 20.0 |

Verbatim comments from Round 2:

- *'This is a acceptability study, not a fidelity study'*
- *'As I look at this I don't think it is critical'*
- *'Not fidelity'*
- *'Important influential factor but separate from fidelity outcomes being measured here'*
- *'I think acceptability is distinct from fidelity - but is important'*

- *‘Acceptability is not fidelity - important for implementation but not important for assessing fidelity’*
- *‘Not a priority for most studies’*
- *‘Important implementation outcome, but not essential to fidelity reporting’*
- *‘important and related but not necessarily core to fidelity’*
- *‘I dont see this as critical to fidelity - acceptance is an outcome’*
- *‘Is about the methodology of assessment’*
- *‘I think it is important for the field to come to consensus on whether intervention acceptability is part of fidelity or not. I vote that it should be as, in theory, you design and intervention to be acceptable and you can measure whether or not it was (i.e., fidelity to intended intervention acceptability). Seems analogous to the intervention enactment discussions’*
- *‘Yes, for consistency and also to enable interpretation of results. Perhaps in future this needs to be a standardised measure of its own’*

| | Not important | | | | | Very important | | | |
|---|---|---|---|---|---|---|---|---|---|
| | 1 | 2 | 3 | 4 | 5 | 6 | 7 | 8 | 9 |
| ITEM 19 | | | | | | | | | |

Please provide any comments or suggestions you have about ITEM 19.

**ITEM 20: Report** the extent to which intervention participants **accept** the intervention (e.g., their perceptions, beliefs, attitudes, and intentions to use the intervention).

Ratings from Round 2:

| **Median (IQR)** | % of panellists rating this item as **1-3** | % of panellists rating this item as **4-6** | % of panellists rating this item as **7-9** |
|---|---|---|---|
| 3 (2, 5) | 61.8 | 20.0 | 18.2 |

Verbatim comments from Round 2:

- *‘Not fidelity assessment but acceptability’*

- *'Important influential factor but separate from fidelity outcomes being measured here'*
- *'Important implementation outcome, but not essential to fidelity reporting'*
- *'Part of readiness for change not fidelity'*
- *'As in ITEM 19, part of acceptability testing but not fidelity reporting'*
- *'Acceptance is more of an outcome of the intervention not a measure of fidelity'*
- *'Yes. Could be likert scale. Doesn't need to be more detailed if methods are reported'*
- *'Belief may be placebo? But might be important to discuss'*

| | Not important | | | | | | | | Very important |
|---|---|---|---|---|---|---|---|---|---|
| | 1 | 2 | 3 | 4 | 5 | 6 | 7 | 8 | 9 |
| ITEM 20 | | | | | | | | | |

Please provide any comments or suggestions you have about ITEM 20.

**ITEM 21:** Report intervention **participants' characteristics** (e.g., age, gender, comorbid conditions, personality traits) that can influence their engagement with the intervention.

Ratings from Round 2:

| **Median (IQR)** | % of panellists rating this item as **1-3** | % of panellists rating this item as **4-6** | % of panellists rating this item as **7-9** |
|---|---|---|---|
| 3 (2, 5.5) | 63.6 | 14.5 | 21.8 |

Verbatim comments from Round 2:

- *'Not fidelity assessment but for general description about intervention'*
- *'Not core to fidelity'*
- *'Part of inclusion, not fidelity'*
- *'This is more of a secondary analysis rather than the fidelity assessment itself'*

- *'General reporting, indeed'*
- *'Part of overall project reporting'*
- *'General reporting rather than fidelity reporting'*
- *'As before, I suggest not overdoing it. This is a critical reporting item from CONSORT, not a fidelity guideline. I am concerned this exercise is trying to overdo things'*
- *'Helpful for a mediational analysis but see this a different to fidelity checks / reporting - which might provide variables for the mediational analysis'*
- *'If fidelity is considered delivery, receipt and enactment, then yes, needs to be reported. It impacts evaluation of the intervention being tested. Not necessary for fidelity of delivery only. Is necessary for evaluating whether or not it was the intervention that achieved the outcome'*
- *'I think fidelity should perhaps report if it's been designed for individual characteristics and if delivered to individuals outside of those characteristics it will be important but not relevant this way'*
- *'Important consideration for many studies'*

| | Not important | | | | | Very important | | | |
|---|---|---|---|---|---|---|---|---|---|
| | 1 | 2 | 3 | 4 | 5 | 6 | 7 | 8 | 9 |
| ITEM 21 | | | | | | | | | |

Please provide any comments or suggestions you have about ITEM 21.

**ITEM 23-26**

**ITEM 25:** Report whether **adaptations made** to the intervention are consistent with the flexibility defined in the protocol, if applicable.

Ratings from Round 2:

| **Median (IQR)** | % of panellists rating this item as **1-3** | % of panellists rating this item as **4-6** | % of panellists rating this item as **7-9** |
|---|---|---|---|

| | | | |
|---|---|---|---|
| 7 (5, 8) | 16.4 | 21.8 | 61.8 |

Verbatim comments from Round 2:

- *'Part of general reporting'*
- *'Trial reporting, fidelity reporting is about whether the permitted adaptations were then delivered ??'*
- *'This is a adaptation study'*
- *'Not important as adaptations could simply be translation into another language'*
- *'It is important to describe changes or adaptations in non-pharmacological interventions, particularly in therapeutic exercise interventions'*
- *'Agree useful to describe adaptations - whether consistent with protocol is a more subjective judgement'*
- *'This is a useful component that invites transparency in fidelity reporting, and can build off of items 23 and 24'*
- *'Very important since it changes the meaning of fidelity ratings'*
- *'This is the anthesis of fidelity. Adaptations compromise the fidelity. However, they should be reported as such'*
- *'If protocol allows adaptations then it is being followed. If not allowed then not part of protocol. Does not need a separate heading'*

| | Not important | | | | | Very important | | | |
|---|---|---|---|---|---|---|---|---|---|
| | 1 | 2 | 3 | 4 | 5 | 6 | 7 | 8 | 9 |
| ITEM 25 | | | | | | | | | |

Please provide any comments or suggestions you have about ITEM 25.

**ITEM 27-34**

Please rate the importance of including the items below in a reporting guideline for intervention fidelity in non-drug, non-surgical trials.

Note: Some items are closely worded but their main substance is unique (highlighted in **bold**). Comments from Round 2 are reported verbatim, with some repeated across items as provided by the panellists.

**ITEM 27:** Report the extent to which the **actual intervention providers in the trial met the provider selection criteria or expected characteristics** described in the protocol (e.g., training, experience, qualifications).

Ratings from Round 2:

| **Median (IQR)** | % of panellists rating this item as **1-3** | % of panellists rating this item as **4-6** | % of panellists rating this item as **7-9** |
|---|---|---|---|
| 7 (5, 8) | 20.0 | 20.0 | 60.0 |

Verbatim comments from Round 2:

- *'Important, but maybe not fidelity?'*
- *'This is process evaluation rather than fidelity. Here we are interested in whether the intervention was delivered as planned. This may help us understand why fidelity was / was not achieved but not if it was achieved'*
- *'Important for trial reporting, not fidelity reporting. Fidelity reporting should focus on whether or not the intervention was delivered, received, enacted as intended. Not the training. Training is methods and trial reporting'*
- *'Not relevant to fidelity'*
- *'It feels unlikely that this would happen, so not a core component'*
- *'This is one that overlaps between intervention methods and fidelity'*
- *'Has overlap with intervention reporting. While this is strongly linked to "whether implemented as planned:, ie fidelity, I suggest it's not essential to understand fidelity'*
- *'Agree where specified, but often not specified'*
- *'This is a prerequisite for provider recruitment therefore not necessary reporting'*
- *'Why would use a provider who didn't meet requirements?'*
- *'This is not essential. If they did not meet the criteria, they should not be there. However, this is somewhat similar to interventions, where we train people to deliver something, but it doesn't always mean they deliver it as intended'*
- *'This is arbitrary. Reporting the characteristics of people delivering the intervention is sufficient'*

- *‘If it is specified in the intervention, then it is relevant to its fidelity’*
- *‘This is essential in complex interventions’*
- *‘This is not onerous and is a single sentence potentially if you have a single person delivering an intervention. If you have more than one, this is important to understanding therapist drift’*
- *‘This would be essential to drawing conclusions regarding fidelity delivery. - if you are including whether deliverers were trained (another item we rate), it would be crucial to also report the proportion of underwent the training (if it wasn't 100% as a requirement for inclusion)’*
- *‘This is important to help understand factors that are common to influencing intervention effectiveness’*
- *‘Important, but not always applicable. E.g. the selection criterion might just have been that it is a person in a leadership position in an organization and otherwise, there would be not participation’*
- *‘Ensuring that intervention providers meet the pre-specified selection criteria is essential for fidelity. Deviations in provider characteristics can lead to variability in intervention delivery and may compromise the validity of the trial’*

| | Not important | | | | | Very important | | | |
|---|---|---|---|---|---|---|---|---|---|
| | 1 | 2 | 3 | 4 | 5 | 6 | 7 | 8 | 9 |
| ITEM 27 | | | | | | | | | |

Please provide any comments or suggestions you have about ITEM 27.

**ITEM 28: Describe a method** to assess **provider competence and proficiency** to deliver the intervention (e.g., assessment of provider knowledge and skills).

Ratings from Round 2:

| **Median (IQR)** | % of panellists rating this item as **1-3** | % of panellists rating this item as **4-6** | % of panellists rating this item as **7-9** |
|---|---|---|---|
| 5 (4, 8) | 23.6 | 40.0 | 36.4 |

Verbatim comments from Round 2:

- *‘This is not important aspect as this is a sub component of assessment of training for providers’*
- *‘This is one that overlaps between intervention methods and fidelity’*
- *‘As stated the training is an intervention. Fidelity issues apply to this intervention but should be tested separately’*
- *‘May be valuable in the context of the particular study, but not essential for fidelity reporting per se’*
- *‘Fidelity doesn't necessarily flow from competence’*
- *‘Not essential’*
- *‘While its important for interventions where providers play a key role, it is not always relevant’*
- *‘Not 'core' to the reporting of intervention fidelity’*
- *‘I feel there are too many problems to overcome and in considering essential criteria I don't think this is necessary (that may change in the future)’*
- *‘Better to assess actual delivery (behaviour) rather than competence, which is more of a determinant of competence’*
- *‘May be difficult to assess and may restrain generalisability’*
- *‘Good luck getting this info funded’*
- *‘Perhaps assessing training (eg did providers learn what they were supposed to after training) is a more feasible way to measure this’*
- *‘This is rather essential to good delivery, though difficult/costly to assess in clinician delivered interventions. If proficiency to deliver the intervention is assessed, the methods for doing so would need to be reported’*
- *‘This is a depends answer. If you are assessing a technique then yes, if you are looking at an overall result / outcome from a package of interventions, then no’*
- *‘This is essential in complex interventions’*
- *‘If specified in the protocol’*
- *‘Often an important consideration’*
- *‘About improving practice’*
- *‘Yes, I think this is the essence of fidelity’*

| | Not important | | | | | Very important | | | |
|---|---|---|---|---|---|---|---|---|---|
| | 1 | 2 | 3 | 4 | 5 | 6 | 7 | 8 | 9 |
| ITEM 28 | | | | | | | | | |

Please provide any comments or suggestions you have about ITEM 28.

**ITEM 29: Report provider competence and proficiency** to deliver the intervention (e.g., comprehension of the intervention protocol, demonstration of relevant skills required to deliver the intervention, etc).

Ratings from Round 2:

| **Median (IQR)** | % of panellists rating this item as **1-3** | % of panellists rating this item as **4-6** | % of panellists rating this item as **7-9** |
|---|---|---|---|
| 5 (4.5, 8) | 23.6 | 30.9 | 45.5 |

Verbatim comments from Round 2:

- *'Fidelity doesn't necessarily flow from competence'*
- *'This is not important aspect as this is a sub component of assessment of training for providers'*
- *'This is one that overlaps between intervention methods and fidelity'*
- *'Training plan and whether it is undertaken by all is sufficient as how to we really know whether some is component and proficient?'*
- *'Better to assess actual delivery (behaviour) rather than competence, which is more of a determinant of competence'*
- *'Not essential'*
- *'Often an eligibility criteria and not relevant'*
- *'Very difficult to measure. Also, changes over time, so would need to measured at multiple time points'*
- *'Depends on whether this is specified in the protocol'*
- *'This may be too subjective and difficult to implement into pragmatic trials'*
- *'If the study is intervention specific: a surgical technique, a manual therapy skill, then yes. If it is about a package of care or something like CFT, then no'*
- *'Perhaps assessing training (eg did providers learn what they were supposed to after training) is a more feasible way to measure this'*

- *‘This is more important than item 28 but completion of training is considered sufficient for assessing competency. It would be more useful to have a report than a description of methods though I am unsure how essential this is. That said, a provider lacking in knowledge and skills is unlikely to deliver the intervention as intended’*
- *‘If proficiency to deliver the intervention is assessed, then reporting the results of such assessments would be essential’*
- *‘About improving practice’*
- *‘This i feel is important’*

| | Not important | | | | | Very important | | | |
|---|---|---|---|---|---|---|---|---|---|
| | 1 | 2 | 3 | 4 | 5 | 6 | 7 | 8 | 9 |
| ITEM 29 | | | | | | | | | |

Please provide any comments or suggestions you have about ITEM 29.

**ITEM 30:** Describe an **intervention provider training plan** including training content, amount, and methods used to deliver the training (e.g, training manuals, didactic sessions, role modelling, supervision with feedback).

Ratings from Round 2:

| **Median (IQR)** | % of panellists rating this item as **1-3** | % of panellists rating this item as **4-6** | % of panellists rating this item as **7-9** |
|---|---|---|---|
| 7 (4.5, 8) | 21.8 | 21.8 | 56.4 |

Verbatim comments from Round 2:

- *‘This is not directly about fidelity assessment’*
- *‘This can be reported in the paper, but not needed in the tool...the more I think about it I dont think it affect fidelity’*
- *‘This should be in the intervention protocol’*

- *‘This is an intervention and should be treated as such’*
- *‘Important to scale up intervention but not essential to assess fidelity’*
- *‘Great to have but not essential fidelity reporting. I think fidelity reporting is more whether you got there than how you got there. How you got there is informative and supports fidelity but is separate’*
- *‘Useful for identifying why fidelity levels achieved, but not fidelity per se’*
- *‘I think these are strategies that are likely to improve fidelity but may not be quite the same as fidelity’*
- *‘It is important to BRIEFLY do this - detailed training materials would not be essential in my view, owing to the need for brevity in fidelity reporting’*
- *‘About improving practice’*
- *‘If specified in the protocol’*
- *‘Again, this is more important than the previous two items and I feel this should be a minimum requirement. Again, difficult with a single intervention provider but still important to state’*
- *‘This seems more relevant for intervention development.... the evaluation of whether this happens in practice would be more related to fidelity’*
- *‘If it is specified in the intervention, then it is relevant to its fidelity’*

| | Not important | | | | | Very important | | | |
|---|---|---|---|---|---|---|---|---|---|
| | 1 | 2 | 3 | 4 | 5 | 6 | 7 | 8 | 9 |
| ITEM 30 | | | | | | | | | |

Please provide any comments or suggestions you have about ITEM 30.

**ITEM 32:** Report the level of training **received by intervention providers** (e.g., frequency of training, attendance to training sessions).

Ratings from Round 2:

| **Median (IQR)** | % of panellists rating this item as **1-3** | % of panellists rating this item as **4-6** | % of panellists rating this item as **7-9** |
|---|---|---|---|

| | | | |
|---|---|---|---|
| 5 (5, 7) | 10.9 | 52.7 | 36.4 |

Verbatim comments from Round 2:

- *'To me, this may be less fidelity and more intervention delivery - more relevant to fidelity may be: was it according to protocol (item 31)'*
- *'This is not part of knowing if the intervention was delivered as planned it about understanding why it was or was not delivered as planned Arguably needs a separate study of reporting fidelity of training of therapists delivering non-surgical, non-drug interventions within RCTs'*
- *'Less important though a potential consideration. Depends on study'*
- *'If specified in the protocol'*
- *'As above - relevant in many circumstances'*
- *'Too much detail?'*
- *'Depend on whether training is a component or strategy'*
- *'If it is specified in the intervention, then it is relevant to its fidelity'*
- *'If specified in the protocol'*
- *'More emphasize should be given to provider training'*

| | Not important | | | | | Very important | | | |
|---|---|---|---|---|---|---|---|---|---|
| | 1 | 2 | 3 | 4 | 5 | 6 | 7 | 8 | 9 |
| ITEM 32 | | | | | | | | | |

Please provide any comments or suggestions you have about ITEM 32.

**ITEM 33:** Report whether **booster (i.e., additional) training sessions** were offered and delivered to intervention providers.

Ratings from Round 2:

| **Median (IQR)** | % of panellists rating this item as **1-3** | % of panellists rating this item as **4-6** | % of panellists rating this item as **7-9** |
|---|---|---|---|
| 6 (5, 7) | 14.5 | 36.4 | 49.1 |

Verbatim comments from Round 2:

- *‘To me, this seems less fidelity and more intervention delivery’*
- *‘Part of describing the overall training plan, doesn't need to be standalone item’*
- *‘Good to know. Informative. Not essential for fidelity measure and reporting. Optimising fidelity and reporting fidelity are two different things’*
- *‘Refresher trainings are essential for providers but I think it is less relevant for the guidelines’*
- *‘Again should be specified in the protocol and reported as delivered in the fidelity reporting’*
- *‘This could be included within item 30 in brackets i.e. (including booster training)’*
- *‘This is about therapist training & support not if intervention was delivered as planned. Part of process evaluation’*
- *‘About improving practice’*
- *‘Relevant to explain if this was needed and why’*
- *‘This is hardly ever recorded and I think this is important as we often make comments about maintenance and generalisation and the need for booster and refresher is often raised. Even to say it did not occur’*
- *‘Given that this is important in skill acquisition, yes, especially in a new skill. So now, you are asking for this to be noted with new skills only, if included’*
- *‘Important to scale up intervention’*
- *‘If it is specified in the intervention, then it is relevant to its fidelity. But if not specified in the intervention, not relevant to fidelity. Provision (or not) more likely a possible moderator when faced with upscaling an intervention or delivering an intervention more than once or over a long period’*

| | Not important | | | | | Very important | | | |
|---|---|---|---|---|---|---|---|---|---|
| | 1 | 2 | 3 | 4 | 5 | 6 | 7 | 8 | 9 |
| ITEM 33 | | | | | | | | | |

Please provide any comments or suggestions you have about ITEM 33.

**ITEM 34:** Report whether the **planned number of intervention providers** to deliver the intervention within the trial was achieved.

Ratings from Round 2:

| **Median (IQR)** | % of panellists rating this item as **1-3** | % of panellists rating this item as **4-6** | % of panellists rating this item as **7-9** |
|---|---|---|---|
| 5 (2.5, 7) | 43.6 | 29.1 | 27.3 |

Verbatim comments from Round 2:

- *'This is less relevant for fidelity'*
- *'Feels more relevant for implementation that fidelity'*
- *'Not essential to fidelity'*
- *'About improving practice'*
- *'Report number of intervention providers more important than planned number and whether this was achieved. That's more feasibility'*
- *'Nice to have rather than essential'*
- *'Process evaluation'*
- *'Is an organizational issue'*
- *'This would be self evident. ie within a trial you would report the number of providers. For fidelity you would also report the planned number (this was a previous item)'*
- *'This is quite important as difference if 1 or two highly skilled people deliver or manage to train a wider group of e.g. nurses as later more likely to be relevant for implementation'*

| | Not important | | | | | | Very important | | |
|---|---|---|---|---|---|---|---|---|---|
| | 1 | 2 | 3 | 4 | 5 | 6 | 7 | 8 | 9 |
| ITEM 34 | | | | | | | | | |

Please provide any comments or suggestions you have about ITEM 34.

#### ITEM 35

**ITEM 35:** When applicable, describe **contextual factors** assessed or observed by the research team that may have influenced intervention fidelity assessment or results (e.g., intervention complexity, intervention provider caseload, time spent by providers on small talk vs time delivering the intervention as intended, etc).

Ratings from Round 2:

| **Median (IQR)** | % of panellists rating this item as **1-3** | % of panellists rating this item as **4-6** | % of panellists rating this item as **7-9** |
|---|---|---|---|
| 7 (4.5, 8) | 23.6 | 20.0 | 56.4 |

Verbatim comments from Round 2:

- *'Not core'*
- *'Moderator of IF rather than IF itself'*
- *'This belongs to process evaluation, not treatment fidelity'*
- *'I guess this is more process evaluation'*
- *'Useful point of discussion particularly when fidelity is variable or low. Should not be required for all studies'*
- *'Useful additional info - but not core to fidelity'*
- *'I think this is important, but is often captured more in the health economics'*
- *'Not essential to fidelity'*
- *'More of a practice based factor rather than reporting of fidelity'*
- *'Difficult to determine in most circumstances'*
- *'Should be reported elsewhere (e.g., under study limitations or discussion), but not necessarily a core fidelity guideline reporting item'*
- *'The key is to describe and this could be done in a sentence or a paragraph. More detail about what exactly this means or what is expected for reporting'*
- *'Contextual factors reflect important predictors of fidelity, important for trial reporting but not essential to fidelity reporting'*

- *‘I believe this to be important information for the interpretation of fidelity assessment results, but not essential in the reporting of fidelity itself. Having said that, we could recommend that authors report any additional information they may have that helps to contextualise the observed levels of fidelity. borderline essential’*
- *‘Identifies hard core barriers and facilitators’*
- *‘Yes, I think this fits with the mixed methods reporting and has implications for replication of the intervention’*
- *‘This is important to interpret / explain fidelity results. Perhaps it could fit under requesting authors to report context/interpretation/explanation of their fidelity outcomes’*
- *‘This feels like it is very important for interpretation of fidelity reporting so yes I guess important’*

| | Not important | | | | | Very important | | | |
|---|---|---|---|---|---|---|---|---|---|
| | 1 | 2 | 3 | 4 | 5 | 6 | 7 | 8 | 9 |
| ITEM 35 | | | | | | | | | |

Please provide any comments or suggestions you have about ITEM 35.

**ITEM 36-42**

Please rate the importance of including the items below in a reporting guideline for intervention fidelity in non-drug, non-surgical trials.

Note: Some items are closely worded but their main substance is unique (highlighted in **bold**). Comments from Round 2 are reported verbatim, with some repeated across items as provided by the panellists.

**ITEM 36:** Describe any changes made to the intervention fidelity assessments based on findings/insights gained from the **intervention development process (e.g., piloting the intervention)**.

Ratings from Round 2:

| Median (IQR) | % of panellists rating this item as **1-3** | % of panellists rating this item as **4-6** | % of panellists rating this item as **7-9** |
|---|---|---|---|
| 5 (2, 7) | 36.4 | 36.4 | 27.3 |

Verbatim comments from Round 2:

- *‘Not core’*
- *‘Nice to have but no essential’*
- *‘Not applicable’*
- *‘Useful but not necessary’*
- *‘Would be part of a paper describing the development of the fidelity measurement, but too detailed for a outcome paper’*
- *‘Piloting should be completed prior. Only revisions that need to be reported are revisions made after trial has started. Intervention development otherwise, is method’*
- *‘This is an evaluation result’*
- *‘Only relevant where applicable - would leave this to the authors' discretion’*
- *‘I think this fits into intervention development and/or the fidelity protocol and does not need a separate standard’*
- *‘As long as this is happening prior to the final protocol, it seems less critical to report on this checklist’*
- *‘Might be important, but might also be more about study methods than ultimate fidelity measures. Probably not critical under almost all circumstances’*
- *‘Very essential to report as measures should can be improved after pilot’*
- *‘Should be reported’*

| | Not important | | | | | Very important | | | |
|---|---|---|---|---|---|---|---|---|---|
| | 1 | 2 | 3 | 4 | 5 | 6 | 7 | 8 | 9 |
| ITEM 36 | | | | | | | | | |

Please provide any comments or suggestions you have about ITEM 36.

**ITEM 37:** Specify whether the intervention fidelity **measures/tools were piloted**.

Ratings from Round 2:

| **Median (IQR)** | % of panellists rating this item as **1-3** | % of panellists rating this item as **4-6** | % of panellists rating this item as **7-9** |
|---|---|---|---|
| 4 (3, 5) | 40.0 | 45.5 | 14.5 |

Verbatim comments from Round 2:

- *‘Ideal but not important’*
- *‘Not critical’*
- *‘Not core’*
- *‘This does not seem core to me, as it has more to do with tool development’*
- *‘Piloting is method. Important trial reporting but not fidelity reporting’*
- *‘This is about measurement development, not about fidelity assessment’*
- *‘Not important for fidelity assessment reporting guidelines’*
- *‘This aspect is about doing good research. Is applicable for any instrument that is used in the study’*
- *‘I do think this is part of development of fidelity measures not outcome measures, but doesn't seem highest priority given where the field is currently’*
- *‘Would be nice’*

| | Not important | | | | | Very important | | | |
|---|---|---|---|---|---|---|---|---|---|
| | 1 | 2 | 3 | 4 | 5 | 6 | 7 | 8 | 9 |
| ITEM 37 | | | | | | | | | |

Please provide any comments or suggestions you have about ITEM 37.

**ITEM 38:** Describe how intervention fidelity **measures/tools were developed**. If existing measures/tools are used, specify them and provide their source.

Ratings from Round 2:

| **Median (IQR)** | % of panellists rating this item as **1-3** | % of panellists rating this item as **4-6** | % of panellists rating this item as **7-9** |
|---|---|---|---|
| 5 (3, 7) | 34.5 | 25.5 | 40.0 |

Verbatim comments from Round 2:

- *'This is important but not essential to include'*
- *'Does not belong in reporting guidelines'*
- *'This does not seem core to me, as it has more to do with tool development'*
- *'Belongs in a separate paper, not main paper reporting on fidelity results'*
- *'Fidelity reporting does not need to describe how the tools were developed, however, it does need to describe what tools/measures were used specifically, including their source'*
- *'Describing the actual fidelity measurement, including if measures are new, adapted, validated, is sufficient'*
- *'As stated, this is about doing correct research'*
- *'Important to understanding the resulting assessments of IF'*
- *'Absolutely for replicability'*
- *'Good to provide base references to tools used'*
- *'Essential for future research build-up'*
- *'Important to help avoid interpretation bias'*

| | Not important | | | | | Very important | | | |
|---|---|---|---|---|---|---|---|---|---|
| | 1 | 2 | 3 | 4 | 5 | 6 | 7 | 8 | 9 |
| ITEM 38 | | | | | | | | | |

Please provide any comments or suggestions you have about ITEM 38.

**ITEM 39:** Specify how the intervention fidelity **measures/tools align with the intervention content and activities**.

Ratings from Round 2:

| **Median (IQR)** | % of panellists rating this item as **1-3** | % of panellists rating this item as **4-6** | % of panellists rating this item as **7-9** |
|---|---|---|---|
| 6 (4, 7) | 12.7 | 50.9 | 36.4 |

Verbatim comments from Round 2:

- *'Not critical'*
- *'I dont see this as "essential" to fidelity reporting - more a nice to have'*
- *'Seems pretty straightforward and self-explanatory - no need for additional reporting'*
- *'This should be self-evident and does not need to be included as a separate reporting item'*
- *'Useful but not essential. I agree with another person that these are sometimes self evident and do not need reporting'*
- *'Should be addressed somehow. Could be simply through the inclusion of the fidelity measurement system as part of Supplementary Materials via OSF or in the publication'*
- *'I am not sure if describing how the intervention fidelity measures/tools align with the intervention content and activities should be part of fidelity measurement in a trial'*
- *'Fidelity should measure what the intervention is about'*
- *'Important to understanding the resulting assessments of IF, as item 38'*
- *'Generally important'*
- *'I think this is important as it is key to the theoretical underpinning of the fidelity protocol. We need to know if what is being measured is actually measuring what is relevant'*

| | Not important | | | | | Very important | | | |
|---|---|---|---|---|---|---|---|---|---|
| | 1 | 2 | 3 | 4 | 5 | 6 | 7 | 8 | 9 |
| ITEM 39 | | | | | | | | | |

Please provide any comments or suggestions you have about ITEM 39.

**ITEM 40: Describe each item** in the intervention fidelity measure/tool and the **process of their selection or development.**

Ratings from Round 2:

| **Median (IQR)** | % of panellists rating this item as **1-3** | % of panellists rating this item as **4-6** | % of panellists rating this item as **7-9** |
|---|---|---|---|
| 4 (3, 5) | 45.5 | 41.8 | 12.7 |

Verbatim comments from Round 2:

- *‘Not critical’*
- *‘Ideal, but not important’*
- *‘This feels like overkill and would be done elsewhere like in a fidelity paper but reporting in a trial it is not necessary’*
- *‘A level of detail that isn't appropriate for most studies’*
- *‘Too much detail, would be at the expense of reporting other more important items’*
- *‘Not necessary in detail for each item’*
- *‘Developing a measurement tool is a different study. A study in itself’*
- *‘I'm not sure they have to have reported it but maybe there is something about providing reference to measure development’*
- *Again, not highest priority given all the other items on this checklist’*
- *‘Agree that the amount of information provided would depend on the focus of the manuscript’*
- *‘Process of selection is not as important as availability of the items’*
- *‘For replicability- there needs to be logic behind and evidence if possible’*

| | Not important | | | | | Very important | | | |
|---|---|---|---|---|---|---|---|---|---|
| | 1 | 2 | 3 | 4 | 5 | 6 | 7 | 8 | 9 |
| ITEM 40 | | | | | | | | | |

Please provide any comments or suggestions you have about ITEM 40.

**ITEM 41:** Specify the **perceived usability of the fidelity measures/tools** (e.g., how straightforward they are to use).

Ratings from Round 2:

| **Median (IQR)** | % of panellists rating this item as **1-3** | % of panellists rating this item as **4-6** | % of panellists rating this item as **7-9** |
|---|---|---|---|
| 3 (2, 4) | 61.8 | 29.1 | 9.1 |

Verbatim comments from Round 2:

- *'This does not seem core to me, as it has more to do with tool development'*
- *'Not relevant'*
- *'Does not belong in reporting guidelines'*
- *'People will have different views'*
- *'Unnecessary. Just describe the tool used, including clear and reproducible operational definitions'*
- *'Belongs in separate work'*
- *'Only relevant to papers about the tool development - not for a trial using the tool'*
- *'It is common for any measures'*
- *'Might be useful, but not essential'*
- *'Yes, report in one line with information about other aspects of fidelity measurement tools'*

| | Not important | | | | | | | | Very important |
|---|---|---|---|---|---|---|---|---|---|
| | 1 | 2 | 3 | 4 | 5 | 6 | 7 | 8 | 9 |
| ITEM 41 | | | | | | | | | |

Please provide any comments or suggestions you have about ITEM 41.

**ITEM 42:** Describe the **strengths and limitations** of the intervention fidelity assessment methods used.

Ratings from Round 2:

| **Median (IQR)** | % of panellists rating this item as **1-3** | % of panellists rating this item as **4-6** | % of panellists rating this item as **7-9** |
|---|---|---|---|
| 5 (3, 6) | 32.7 | 45.5 | 21.8 |

Verbatim comments from Round 2:

- *'Nice to have – not essential'*
- *'Relevant in a feasibility context, but not essential to understand fidelity'*
- *'Might substantially further our understanding of how fidelity affects intervention outcomes. So, would be nice, but probably not essential at this point'*
- *'Belongs in manuscript or thesis'*
- *'Not the focus of most studies'*
- *'No essential only for transparency'*
- *'Same conclusion. You need good tools to measure fidelity, but developing these stools will need to be done before a trial and will be guided by the content and procedures of the intervention'*
- *'All strengths and limitations should be reported. Learnings from researcher feedback after implementing the intervention may provide valuable insights into fidelity and possible reasons for outcome'*

| | Not important | | | | | | Very important | | |
|---|---|---|---|---|---|---|---|---|---|
| | 1 | 2 | 3 | 4 | 5 | 6 | 7 | 8 | 9 |
| ITEM 42 | | | | | | | | | |

Please provide any comments or suggestions you have about ITEM 42.

#### ITEM 43-46

Please rate the importance of including the items below in a reporting guideline for intervention fidelity in non-drug, non-surgical trials.

Note: Some items are closely worded but their main substance is unique (highlighted in **bold**). Comments from Round 2 are reported verbatim, with some repeated across items as provided by the panellists.

**ITEM 43:** If **different weights were assigned to items** in an intervention fidelity measure/tool that has not been externally validated, report how these weights were determined (e.g., theory, empirical evidence, sensitivity analysis) and the rationale for this approach.

Ratings from Round 2:

| **Median (IQR)** | % of panellists rating this item as **1-3** | % of panellists rating this item as **4-6** | % of panellists rating this item as **7-9** |
|---|---|---|---|
| 5 (4, 6) | 21.8 | 60.0 | 18.2 |

Verbatim comments from Round 2:

- *'Too detailed and should be part of a separate validation of said fidelity tool (not "core")'*
- *'Important but not critical'*
- *'I agree with the commends from round 1; and practically speaking, this will not apply in a lot of cases as this level of sophistication in fidelity assessment has, generally, not yet been achieved'*
- *'I'm rating this low as it's not core and actually could limit advancement of science by advocating non validated tools(although validated tools in this area are hard to find it is true)'*
- *'More related to building a measure I would have thought'*
- *'It's a lot of detail - hard to know how granular to go with any of this'*
- *'Development of tool should be explain with detail'*

- *‘Does not need weighting. Just reporting’*
- *‘If relevant, then it is important to know how weighting was applied’*
- *‘Only if weights used’*
- *‘This belongs to psychometric studies assessing and exploring methods to measure treatment fidelity’*
- *‘If it is done, it needs to be explained or there needs to be a reference to another paper where it is explained’*
- *‘This is important for replication’*
- *‘Where relevant’*
- *‘Tno relevant only for rigor and transparency’*

| | Not important | | | | | Very important | | | |
|---|---|---|---|---|---|---|---|---|---|
| | 1 | 2 | 3 | 4 | 5 | 6 | 7 | 8 | 9 |
| ITEM 43 | | | | | | | | | |

Please provide any comments or suggestions you have about ITEM 43.

**ITEM 44: Specify the psychometric properties** of the intervention fidelity measures/tools (e.g., reliability, validity, and sensitivity to change).

Ratings from Round 2:

| **Median (IQR)** | % of panellists rating this item as **1-3** | % of panellists rating this item as **4-6** | % of panellists rating this item as **7-9** |
|---|---|---|---|
| 5 (3, 5.5) | 32.7 | 47.3 | 20.0 |

Verbatim comments from Round 2:

- *‘At this stage not core’*
- *‘Good to have but not essential’*
- *‘Most measures do not have psychometric analyses as they have been created specifically for a trial. Yes, it may be good practice but not important’*

- *‘Important but will vary depending on method used to assess fidelity (e.g., self-report, observation)’*
- *‘I have suggested that thorough fidelity work in trials requires a study within a study which is often not possible. Brief reporting regarding fidelity methods will be required in a reporting guideline but many of the details like psychometrics of the fidelity measurement tools will need to be rolled up into one reporting item’*
- *‘Often descriptive and not a standardised instrument, so likely not very useful in practice’*
- *‘When we have a standardised tool, yes. Until then, no. Just report the method used’*
- *‘While it promotes transparency and rigour in fidelity evaluation, it is not universally applicable or relevant’*
- *‘You may have generally formulated instrument items for which this can be done. But also very intervention specific questions’*
- *‘In some fidelity assessment of online-based intervention, measures are objective and psychometric properties are irrelevant’*
- *‘While important, we should acknowledge that developing and validating fidelity measures may be too resource intensive to be practical, and sometimes also doesn't seem necessary. For example, if you count number of attended sessions by means of a checklist or register, you probably don't have to validate that’*
- *‘Should be done if psychometric properties are known, but this would be highly unusual’*

| | Not important | | | | | Very important | | | |
|---|---|---|---|---|---|---|---|---|---|
| | 1 | 2 | 3 | 4 | 5 | 6 | 7 | 8 | 9 |
| ITEM 44 | | | | | | | | | |

Please provide any comments or suggestions you have about ITEM 44.

**ITEM 45:** Specify the benchmarks used to **interpret the psychometrics** of the intervention fidelity measures/tools. For example, report what constitutes an acceptable level of inter-rater reliability.

Ratings from Round 2:

| **Median (IQR)** | % of panellists rating this item as **1-3** | % of panellists rating this item as **4-6** | % of panellists rating this item as **7-9** |
|---|---|---|---|
| 4 (3, 6) | 45.5 | 34.5 | 20.0 |

Verbatim comments from Round 2:

- *‘Not essential for core fidelity reporting’*
- *‘Only if repeating a fidelity measure or using a validated tool’*
- *‘Too detailed for fidelity work unless it is a separate fidelity study’*
- *‘This is "nice to have" additional detail - this is not essential in my view’*
- *‘Likely not available or relevant in most situations’*
- *‘I think we can agree that measuring fidelity requires appropriate instruments (valid and reliable). Authors should mention this in their articles. Maybe one can standardize some concepts of fidelity and the way they should be measured. Just for inter-intervention comparison. This is a task in itself’*
- *‘Use standardised measure when exists. Walton has started this with low, mod, high %’*
- *‘Important but will vary depending on method used to assess fidelity (e.g., self-report, observation)’*
- *‘Psychometrics are so rarely known that this would not be helpful at this time’*
- *‘While it promotes transparency and rigour in fidelity evaluation, it is not universally applicable or relevant’*
- *‘It is a general practice for any tool development process’*

| | Not important | | | | | Very important | | | |
|---|---|---|---|---|---|---|---|---|---|
| | 1 | 2 | 3 | 4 | 5 | 6 | 7 | 8 | 9 |
| ITEM 45 | | | | | | | | | |

Please provide any comments or suggestions you have about ITEM 45.

**ITEM 46:** Specify an a priori threshold for **acceptable level of intervention fidelity** or cut-offs.

Ratings from Round 2:

| **Median (IQR)** | % of panellists rating this item as **1-3** | % of panellists rating this item as **4-6** | % of panellists rating this item as **7-9** |
|---|---|---|---|
| 7 (5, 8) | 9.1 | 38.2 | 52.7 |

Verbatim comments from Round 2:

- *'Not helpful'*
- *'We don't yet have evidence-based thresholds, so these would be largely made up and useless. Once an evidence-based threshold is known, though, it should be used'*
- *'I believe this to be part of a study's interpretation (and maybe a useful item for most feasibility studies), but not essential to understand fidelity itself'*
- *'Dichotomising fidelity into being acceptable/not is overly simplistic - it also loses important nuance'*
- *'Not necessary. Just report fidelity identified. Let reader decide acceptability'*
- *'I think it is more useful to understand context and have meaningful interpretation of how fidelity/lack thereof impacted the trial as opposed to a dichotomous cut-off, which may end up being quite arbitrary and meaningless'*
- *'I can't visualise how this will be done meaningfully given we're not even fully agreed on what intervention fidelity means yet'*
- *'Not sure one can really do this, seems somewhat arbitrary'*
- *'Think it is too hard and unscientific to determine a black and white 'cut off''*
- *'Don't think this is typically feasible given the ad hoc / study specific way fidelity is often measured. Fidelity assessment is not yet this sophisticated in most cases'*
- *'This is difficult if the intervention is currently being tested - may not be known what components of the intervention were or were not effective - this is part of what the initial fidelity measurements help us to understand. If it is an established intervention with evidence that it influences outcomes then think this is possible but for pilot studies may be trickier. Will also depend on all individual interventions'*

- *'Needs to be reported, but the actual data needs to be more specific than just above/below threshold'*
- *'This is more important that item 45'*
- *'This is relevant for understanding the applicability of the results'*
- *'Important consideration in many studies for per protocol analyses'*
- *'I rated this item as very important because specifying an a priori threshold ensures that fidelity is assessed objectively and systematically. It provides a clear benchmark for evaluating whether the intervention was delivered as intended'*
- *'where appropriate pre-specify OR have this as an objective - collect fidelity data in order to help determine cut off (for example in pilot feasibility RCTs.....). Fidelity guidance might need to be separated out for pilots versus full trials.....'*
- *'Is needed. You have an ambition, so make clear what that is. It helps to identify what was revealed as expected and what was not. You need this information'*
- *'I think this is critical'*
- *'I think this is essential and although often arbitrary should at least be reported to aid interpretation'*

| | Not important | | | | | Very important | | | |
|---|---|---|---|---|---|---|---|---|---|
| | 1 | 2 | 3 | 4 | 5 | 6 | 7 | 8 | 9 |
| ITEM 46 | | | | | | | | | |

Please provide any comments or suggestions you have about ITEM 46.

**ITEM 47-50**

**ITEM 50:** Report intervention fidelity **in terms of session fidelity** (i.e., fidelity to intervention core components within sessions). *Session is defined as a distinct unit of time during which the intervention is delivered. Synonyms include 'visit', 'consultation', 'meeting', 'event', etc.

Ratings from Round 2:

| **Median (IQR)** | % of panellists rating this item as **1-3** | % of panellists rating this item as **4-6** | % of panellists rating this item as **7-9** |
|---|---|---|---|
| 6 (5, 8) | 3.6 | 58.2 | 38.2 |

Verbatim comments from Round 2:

- *‘Important but could be optional or if core then researchers required to pre-specify if all or some components are reported on’*
- *‘The relevance of this item depends on the type of the intervention’*
- *‘Summary reporting here would be sufficient and helpful for avoiding too much small detail in reporting’*
- *‘It depends on the nature of the intervention. I believe guidelines should specify that a level of analysis must be included, but they should not dictate what this analysis should be’*
- *‘Valuable if applicable to the intervention, but also pretty intensive and might not be highest priority’*
- *‘I think this becomes difficult to distinguish (from item 49) and therefore may overcomplicate reporting’*
- *‘Critical when session fidelity predicts outcomes in that session, but would be overkill if fidelity was near perfect in all sessions’*
- *‘Researchers need to report this. Reporting is distinct from assessment. Depends on the intervention. Some interventions are very flexible. Then you should find out what happened doing an extra effort. If providers or participants can only follow one strict process, thus less flexibility, finishing the intervention implies that all aspects were covered. In both cases, you report dose to explain the response’*

| | Not important | | | | | Very important | | | |
|---|---|---|---|---|---|---|---|---|---|
| | 1 | 2 | 3 | 4 | 5 | 6 | 7 | 8 | 9 |
| ITEM 50 | | | | | | | | | |

Please provide any comments or suggestions you have about ITEM 50.

#### ITEM 51

**ITEM 51:** Describe a **specific intervention fidelity assessment and monitoring protocol** prior to trial analysis.

Ratings from Round 2:

| **Median (IQR)** | % of panellists rating this item as **1-3** | % of panellists rating this item as **4-6** | % of panellists rating this item as **7-9** |
|---|---|---|---|
| 7 (5, 8) | 3.6 | 40.0 | 56.4 |

Verbatim comments from Round 2:

- *‘Might be useful to do, but not necessary to report. In pilot or inductive research, measure may need to change from initially developed to final use’*
- *‘Relevant to trial design not intervention fidelity design’*
- *‘This is important but not critical in my view’*
- *‘This is more reporting on optimising fidelity than fidelity itself’*
- *‘Might be useful but not necessarily realistic as plans change, or feasible due to study burden’*
- *‘I think v important for efficacy trials, but maybe less so for implementation’*
- *‘Depends on the study’*
- *‘This should ideally be reported but often unrealistic given time and resource constraints and the need to balance fidelity with context, which is difficult to report in an a priori protocol’*
- *‘These procedures should be developed a priori’*
- *‘I cannot imagine any reason why there should not be a fidelity assessment protocol. Otherwise we might just be making it up as we go along. We could discuss when this should be finalised. Unlikely to be feasible for most studies before start of the project and leaving this until just before main trial analysis is far too late. So perhaps before the end of the intervention delivery phase of the trial. So reporting guideline becomes 'Completed fidelity protocol agreed before intervention delivery completion with date of completion' Or 'Date fidelity assessment and monitoring protocol finalised'*
- *‘Essential’*

- *‘Describing a specific fidelity assessment and monitoring protocol ensures that fidelity evaluation is planned systematically before the trial begins, avoiding ad hoc or retrospective assessments that can introduce bias or inconsistencies’*
- *‘As mentioned before, an ideal certainly, possibly essential for good practice’*
- *‘I believe this is critical. While I understand the challenges involved in conducting trials, the work still needs to be done. I feel that far too many trials are published with limitations due to oversights in fidelity, which ultimately undermines their value’*

| | Not important | | | | | Very important | | | |
|---|---|---|---|---|---|---|---|---|---|
| | 1 | 2 | 3 | 4 | 5 | 6 | 7 | 8 | 9 |
| ITEM 51 | | | | | | | | | |

Please provide any comments or suggestions you have about ITEM 51.

**ITEM 52-53**

**ITEM 52:** Specify when **intervention fidelity data were analysed**, such as concurrently with the intervention or after the intervention period was completed.

Ratings from Round 2:

| **Median (IQR)** | % of panellists rating this item as **1-3** | % of panellists rating this item as **4-6** | % of panellists rating this item as **7-9** |
|---|---|---|---|
| 7 (4, 7) | 23.6 | 25.5 | 50.9 |

Verbatim comments from Round 2:

- *‘I don’t think this time frame changes the results’*
- *‘Only if important for understanding study process (like change in study procedure due to fidelity measures) or fidelity measurement (like change in measurement system midway through study)’*

- *‘This leads into a wider question as to whether any fidelity assessment is feeding back into intervention delivery on the trial. That is I think quality assurance and monitoring rather than fidelity. SO, yes this is important but need clarity on what is done with any data analysed during the trail’*
- *‘Not relevant unless leading to substantive changes to intervention delivery’*
- *‘It is about the study design and the implication of a design choice’*
- *‘Study methods not fidelity’*
- *‘Rarely relevant’*
- *‘In some interventions, it is not possible to assess fidelity at different points during the trial’*
- *‘This would be essential IF, based on concurrent assessment, adjustments to the intervention were made’*
- *‘In some interventions, it is not possible to assess fidelity at different points during the trial’*
- *‘Important for understanding any fidelity assessment’*
- *‘It is very important. If intervention fidelity was assessed concurrently, it is also important to report any changes that arose from this evaluation’*
- *‘Very important as if analysed later it may affect the level of fidelity (e.g., drift)’*
- *‘I think it's probably important when the fidelity analysis was done in relation to trial analysis as this is where risk of bias is greatest’*
- *‘To account for recall bias’*
- *‘An important part of describing how fidelity was measured’*
- *‘Specifying the timing of fidelity analysis adds transparency to the study’s methodology, particularly in cases where concurrent analysis is used to adjust or refine the intervention’*
- *‘This is essential, as it relates to whether trialists adopted an active or passive approach towards treatment fidelity’*

| | Not important | | | | | Very important | | | |
|---|---|---|---|---|---|---|---|---|---|
| | 1 | 2 | 3 | 4 | 5 | 6 | 7 | 8 | 9 |
| ITEM 52 | | | | | | | | | |

Please provide any comments or suggestions you have about ITEM 52.

**ITEM 53:** Specify the timepoints of the intervention where **intervention fidelity data was collected** (e.g., week 1, 6, and 12).

Ratings from Round 2:

| **Median (IQR)** | % of panellists rating this item as **1-3** | % of panellists rating this item as **4-6** | % of panellists rating this item as **7-9** |
|---|---|---|---|
| 7 (6, 8) | 14.5 | 14.5 | **70.9** |

Verbatim comments from Round 2:

- *'Important for understanding any fidelity assessment'*
- *'In some interventions, it is not possible to assess fidelity at different points during the trial'*
- *'Again, would not change the data / results so not essential'*
- *'Specifying the timing of fidelity analysis adds transparency to the study's methodology, particularly in cases where concurrent analysis is used to adjust or refine the intervention'*
- *'This is about proper study design'*
- *'Not relevant'*
- *'Essential as fidelity vary with time specially for enactment'*
- *'Important to know that fidelity measures were not all collected in a single timepoint during multiple timepoint studies'*

| | Not important | | | | | | Very important | | |
|---|---|---|---|---|---|---|---|---|---|
| | 1 | 2 | 3 | 4 | 5 | 6 | 7 | 8 | 9 |
| ITEM 53 | | | | | | | | | |

Please provide any comments or suggestions you have about ITEM 53.

**ITEM 54-55**

Please rate the importance of including the items below in a reporting guideline for intervention fidelity in non-drug, non-surgical trials.

Note: Some items are closely worded but their main substance is unique (highlighted in **bold**). Comments from Round 2 are reported verbatim, with some repeated across items as provided by the panellists.

**ITEM 54:** Describe how intervention fidelity **assessors were trained** to use the intervention fidelity measures/tools and conduct the fidelity assessments.

Ratings from Round 2:

| **Median (IQR)** | % of panellists rating this item as **1-3** | % of panellists rating this item as **4-6** | % of panellists rating this item as **7-9** |
|---|---|---|---|
| 5 (4.5, 7) | 18.2 | 50.9 | 30.9 |

Verbatim comments from Round 2:

- *‘This is more a nice to have rather than an "essential" reporting item’*
- *‘Not essential can be use subject expert with minimum training’*
- *‘Important for transparency transparency the report but not for fidelity assessment’*
- *‘Yes, training of assessors need to be described. But, training is an intervention within an intervention study. Same fidelity issues apply’*
- *‘Very useful for complex or observational fidelity measures. Maybe less so for other systems’*
- *‘Important and probably underreported, but not sure this fits under core fidelity reporting requirements as assessment is often done by the research team itself, which should be trained as part of standard operating procedures and guidelines’*
- *‘Important for more complex procedures but not all’*
- *‘Most non-drug, non-surgical healthcare interventions are complex so intervention deliverer training is pretty essential for understanding fidelity of delivery’*

| | Not important | | | | | Very important | | | |
|---|---|---|---|---|---|---|---|---|---|
| | 1 | 2 | 3 | 4 | 5 | 6 | 7 | 8 | 9 |
| ITEM 54 | | | | | | | | | |

Please provide any comments or suggestions you have about ITEM 54.

**ITEM 55:** Specify if there was any requirement for the **selection of intervention fidelity assessors** (e.g., be a health registered professional).

Ratings from Round 2:

| **Median (IQR)** | % of panellists rating this item as **1-3** | % of panellists rating this item as **4-6** | % of panellists rating this item as **7-9** |
|---|---|---|---|
| 5 (3, 6) | 34.5 | 47.3 | 18.2 |

Verbatim comments from Round 2:

- *‘This is more a nice to have rather than an "essential" reporting item’*
- *‘Training of fidelity assessors and fidelity of the assessment itself is probably more important than the selection of potential assessors’*
- *‘As stated, a training is an intervention in itself. Fidelity issues for an intervention apply’*
- *‘Who the assessors were is important to know’*
- *‘More important to know their role and that they were trained to measure fidelity accurately. Since assessors may not be one homogenous group’*
- *‘Think this is probably too much detail for a reporting checklist’*
- *‘Maybe useful, but titles are probably not predictive of actual skills in assessing fidelity’*
- *‘May be relevant in some circumstances but not the majority where no special training is needed’*
- *‘Relevant for understanding delivery competence’*

- *'This is tricky, but I agree that outlining the procedure for training assessors and ensuring inter-rater reliability is more important. To be honest, I think there's probably a role for AI in this!'*
- *'I'd like to see this in the protocol but not necessarily post-hoc reporting. Which brings to mind the question of when you are thinking the guidelines will be used. I'd assumed for post trial evaluation but I guess it could be both pre and post'*

| | Not important | | | | | Very important | | | |
|---|---|---|---|---|---|---|---|---|---|
| | 1 | 2 | 3 | 4 | 5 | 6 | 7 | 8 | 9 |
| ITEM 55 | | | | | | | | | |

Please provide any comments or suggestions you have about ITEM 55.

**ITEM 56**

**ITEM 56:** Report a **clear intervention manual or detailed intervention protocol**.

Ratings from Round 2:

| **Median (IQR)** | % of panellists rating this item as **1-3** | % of panellists rating this item as **4-6** | % of panellists rating this item as **7-9** |
|---|---|---|---|
| 8 (6, 8.5) | 7.3 | 25.5 | 67.3 |

Verbatim comments from Round 2:

- *'Very important, but not fidelity. Part of general reporting'*
- *'Part of intervention reporting, not fidelity'*
- *'While these are highly important for reporting guidelines for intervention, not highly important for reporting guidelines for intervention fidelity. For scientific vigour'*

- *‘This may be more closely related to general intervention reporting than fidelity reporting, to me’*
- *‘Part of trial reporting, not part of fidelity reporting’*
- *‘This is in Tidier etc, so a fidelity guide needs to cross reference to this’*
- *‘Important for intervention development and for fidelity evaluators to have access to when designing fidelity studies - therefore rated lower as not necessarily key for the reporting guidelines for fidelity’*
- *‘This is transparency’*
- *‘The presence of absence of a manual or protocol - this is important for fidelity’*
- *‘This is standard consideration for non-drug trials’*
- *‘Feels essential to fidelity assessment’*
- *‘Important to contextualise fidelity. Vague protocol - high fidelity doesn't align’*
- *‘Very important fidelity assessment should based on the manual’*
- *‘Core’*
- *‘This item is essential to assess fidelity’*

| | Not important | | | | | Very important | | | |
|---|---|---|---|---|---|---|---|---|---|
| | 1 | 2 | 3 | 4 | 5 | 6 | 7 | 8 | 9 |
| ITEM 56 | | | | | | | | | |

Please provide any comments or suggestions you have about ITEM 56.

**ITEM 58**

**ITEM 58:** Report whether the **cultural and linguistic aspects of the intervention** were delivered as planned, if applicable.

Ratings from Round 2:

| **Median (IQR)** | % of panellists rating this item as **1-3** | % of panellists rating this item as **4-6** | % of panellists rating this item as **7-9** |
|---|---|---|---|
| 5 (3, 7) | 27.3 | 45.5 | 27.3 |

Verbatim comments from Round 2:

- *‘Is about intervention development and pilot testing. Depend on the aim of the project’*
- *‘Relatively less important as more important given to adherence and competence in terms of delivery fidelity’*
- *‘This seems self-evident when targeting specific user groups - to be reported in the general study description but not fidelity reporting’*
- *‘Not related to fidelity assessment’*
- *‘Important, but not strictly fidelity’*
- *‘Important, but I don't think it is necessary to highlight it separately, delivery of all intervention aspects should simply be assessed’*
- *‘Doesn't fit with fidelity reporting’*
- *‘Not part of treatment fidelity. May influence treatment fidelity, but that is part of a process evaluation (which is beyond and broader than treatment fidelity alone)’*
- *‘Part of the intervention components/underpinning logic model already no?’*
- *‘The relevance of this item depends on the type of intervention and the context in which the trial is conducted’*
- *‘This is a tricky one as depends what the cultural and linguistic aspects are. If they are components then agree this should be included in reports of components and delivery as planned or not’*
- *‘Where appropriate’*
- *‘If these are core elements of the intervention delivery, then fidelity assessment is important for these aspects’*
- *‘Only if it was part of the trial RQs’*
- *‘If these are core components of the intervention, they would be captured in the fidelity measure at large. It is more important that the measure capture core intervention components than specific cultural components (if those components are not core, they may end up "padding" the measure and masking errors on core components)’*
- *‘If the intervention is well specified this should be incorporated in other questions - not sure it needs to be pulled out specifically although suspect it would help people design interventions better’*

- *'All aspects of the intervention need to be reported on with respect to whether they were delivered as planned. Cultural and linguistic aspects should be reported as part of the intervention and measured as part of intervention delivery, receipt and enactment but not as a specific item of fidelity. They should be specific elements of the intervention itself'*

| | Not important | | | | | | Very important | | |
|---|---|---|---|---|---|---|---|---|---|
| | 1 | 2 | 3 | 4 | 5 | 6 | 7 | 8 | 9 |
| ITEM 58 | | | | | | | | | |

Please provide any comments or suggestions you have about ITEM 58.

**ITEM 59**

**ITEM 59:** Specify the **frameworks and guidance** used to inform intervention fidelity assessments.

Ratings from Round 2:

| **Median (IQR)** | % of panellists rating this item as **1-3** | % of panellists rating this item as **4-6** | % of panellists rating this item as **7-9** |
|---|---|---|---|
| 7 (5, 8) | 7.3 | 40.0 | 52.7 |

Verbatim comments from Round 2:

- *'Might be useful, but is not essential'*
- *'This is not unimportant, as fidelity assessment should be based in a framework, but this does seem less "core" to me, and may result in "name dropping", instead of meaningful, in-depth, and intervention-specific descriptions of how fidelity was assessed'*
- *'Not essential but to be considered'*

- *‘Need to specify how fidelity was assessed. This needs to be standardised. In the meantime, if a framework was used it should be specified but difficult to specify in the reporting guideline that this must be reported, as a framework may not have been used. When there is a standardised measure, this should be used’*
- *‘We need to know there is some sort of theoretical underpinning for what was done’*
- *‘This must be important to understand what is meant by fidelity in any study. We're not at a point where we all mean the same thing by fidelity so until that point this is needed’*
- *‘This is essential for justifying one's' approach to fidelity assessment for reviewers to see...and it requires one to lines in a paper so it is easy to do and include’*

| | Not important | | | | | Very important | | | |
|---|---|---|---|---|---|---|---|---|---|
| | 1 | 2 | 3 | 4 | 5 | 6 | 7 | 8 | 9 |
| ITEM 59 | | | | | | | | | |

Please provide any comments or suggestions you have about ITEM 59.

**ITEM 61**

**ITEM 61:** If not all intervention fidelity domains were assessed, describe the rationale for **excluding those domains**.

Ratings from Round 2:

| **Median (IQR)** | % of panellists rating this item as **1-3** | % of panellists rating this item as **4-6** | % of panellists rating this item as **7-9** |
|---|---|---|---|
| 7 (5, 8) | 5.5 | 34.5 | 60.0 |

Verbatim comments from Round 2:

- *‘Probably not needed in reporting guideline. Really up tot he researchers to decide how they address this issue according to context’*
- *‘Helpful but not essential’*
- *‘I think this is researcher specific unless we agree on a specific tool’*
- *‘Given the goal is a small set of CORE intervention fidelity domains and items for use routinely, this item does not feel core’*
- *‘This is only if there is consensus and clarity regarding what the domains actually are/should be’*
- *‘Important as a part of describing fidelity assessment - ideally all should be assessed and if not, this should be acknowledged’*
- *‘Important to know what measures were or were not used, particularly if the fidelity measure used varies systematically across arms or trial phases’*
- *‘For good practice yes!’*
- *‘Seems obvious to report this. But is not only applying to fidelity’*
- *‘This needs to be encouraged (ise of all domains)*
- *‘All domains should be assessed’*
- *‘Important consideration for the rationale for fidelity assessments’*
- *‘Important to clear report and transparency’*
- *‘Important for understanding any fidelity assessment’*

| | Not important | | | | | Very important | | | |
|---|---|---|---|---|---|---|---|---|---|
| | 1 | 2 | 3 | 4 | 5 | 6 | 7 | 8 | 9 |
| ITEM 61 | | | | | | | | | |

Please provide any comments or suggestions you have about ITEM 61.

**ITEM 62**

**ITEM 62:** Describe how **stakeholders** were involved in the development of the intervention fidelity measures/tools.

Ratings from Round 2:

| **Median (IQR)** | % of panellists rating this item as **1-3** | % of panellists rating this item as **4-6** | % of panellists rating this item as **7-9** |
|---|---|---|---|
| 4 (2.5, 5) | 41.8 | 45.5 | 12.7 |

Verbatim comments from Round 2:

- *'Important but not a core element for reporting intervention fidelity'*
- *'This is important , but not related to fidelity, sounds more related to generalisability and potential for scalability'*
- *'Is more related to the development process of an intervention. Stakeholder involvement maybe essential during every process of a trial from development to final evaluation'*
- *'This would be relevant to a study about tool development, but not for a trial that used the measure'*
- *'Not essential and not relevant most times as the measures are directly built based on the intervention protocol'*
- *'Only for transparency but not for fidelity assessment'*
- *'It would be helpful to know this but not essential'*
- *'Yes especially since the increasing focus on systems and codesign of strategies'*
- *'Anyway stakeholders involvement report in every research process'*

| | Not important | | | | | Very important | | | |
|---|---|---|---|---|---|---|---|---|---|
| | 1 | 2 | 3 | 4 | 5 | 6 | 7 | 8 | 9 |
| ITEM 62 | | | | | | | | | |

Please provide any comments or suggestions you have about ITEM 62.

**ITEM 63**

**ITEM 63:** When applicable, justify the **number of response categories** in the intervention fidelity measure/tool, for example, why a 3-point scale (delivered, partially delivered, not delivered) was chosen over other scales.

Ratings from Round 2:

| **Median (IQR)** | % of panellists rating this item as **1-3** | % of panellists rating this item as **4-6** | % of panellists rating this item as **7-9** |
|---|---|---|---|
| 3 (2, 4.5) | 58.2 | 34.5 | 7.3 |

Verbatim comments from Round 2:

- *'Not relevant for the purpose of a fidelity reporting guideline'*
- *'Nope'*
- *'Seems to specific'*
- *'No matters'*
- *'Beginning to feel we need guidelines for developing valid/reliable fidelity measures and this would go there but not necessarily in a core fidelity reporting guideline'*
- *'Useful but not essential - helpful in the appendix. The fact it is stated is more important than the rationale'*
- *'Need to be careful about this reporting guideline not becoming excessively detailed and difficult to follow'*
- *'Not core'*
- *'Nice but not important'*

| | Not important | | | | | Very important | | | |
|---|---|---|---|---|---|---|---|---|---|
| | 1 | 2 | 3 | 4 | 5 | 6 | 7 | 8 | 9 |
| ITEM 63 | | | | | | | | | |

Please provide any comments or suggestions you have about ITEM 63.

**ITEM 64**

**ITEM 64:** Describe how **data from multiple intervention fidelity measurement methods**, when applicable, are integrated and triangulated to provide comprehensive fidelity results. For example, describe how quantitative data (e.g., from fidelity measures/tools) are combined with qualitative data (e.g., self-reports).

Ratings from Round 2:

| **Median (IQR)** | % of panellists rating this item as **1-3** | % of panellists rating this item as **4-6** | % of panellists rating this item as **7-9** |
|---|---|---|---|
| 6 (5, 8) | 9.1 | 47.2 | 43.6 |

Verbatim comments from Round 2:

- *'I don't think this effect fidelity'*
- *'If applicable'*
- *'Rarely relevant'*
- *'This should be an inherent part of describing how fidelity will be measured - and therefore may not need to be its own item'*
- *'Mixed methods. Not specific for fidelity. This is about normal research'*
- *'Critical only when there are multiple measures of fidelity that are primarily reported as global scores'*
- *'Important if you are trying to come up with an over fidelity "score" but I rarely see this and seems challenging for various reasons'*
- *'I think fidelity reporting can be quantitative. Was the intervention delivered as intended, received as intended and enacted as intended. Parameters can include time-period and recency of reporting to allow for recall bias. Qualitative data is important in the interpretation of results. Why the fidelity measure outcome and how it could be improved or factors contributing to it's strength'*
- *'This reflects the fidelity analysis plan and seems like something that would be important to do with any mixed methods study / data; so if fidelity data were coming from various sources, commenting on integration/triangulation would be necessary'*

| | Not important | | | | | Very important | | | |
|---|---|---|---|---|---|---|---|---|---|
| | 1 | 2 | 3 | 4 | 5 | 6 | 7 | 8 | 9 |
| ITEM 64 | | | | | | | | | |

Please provide any comments or suggestions you have about ITEM 64.

**ITEM 65**

**ITEM 65:** Where applicable, specify the **type of analysis conducted** to obtain intervention fidelity results (e.g., descriptive, ANOVA, qualitative thematic analysis).

Ratings from Round 2:

| **Median (IQR)** | % of panellists rating this item as **1-3** | % of panellists rating this item as **4-6** | % of panellists rating this item as **7-9** |
|---|---|---|---|
| 6 (5, 8) | 10.9 | 40.0 | 49.1 |

Verbatim comments from Round 2:

- *'Not fidelity specific'*
- *'This should be in a statistical analysis plan - not specific to fidelity assessment'*
- *'No critical'*
- *'Agree not essential as assessments won't all be amenable to this type of analysis'*
- *'This will mostly be quite descriptive but in some cases an important consideration'*
- *'This is part of fidelity analysis plan'*
- *'Important, but not really about fidelity reporting'*
- *'Optional but if planned needs to be reported'*
- *'YES including quali - e.g. thematic analysis - coding used'*
- *'If quantitative – yes'*
- *'Very important when fidelity data are quantitative in nature'*

- *‘Specifying methods for assessment is important (though, as noted, this approach will not be relevant to all possible assessments)’*

| | Not important | | | | | | Very important | | |
|---|---|---|---|---|---|---|---|---|---|
| | 1 | 2 | 3 | 4 | 5 | 6 | 7 | 8 | 9 |
| ITEM 65 | | | | | | | | | |

Please provide any comments or suggestions you have about ITEM 65.

**ITEM 66**

**ITEM 66:** If applicable, describe the **rationale for any sensitivity/additional analysis** informed by intervention fidelity data (e.g., intention-to-treat versus removing low fidelity cases, mediation analysis based on fidelity to specific intervention components).

Ratings from Round 2:

| **Median (IQR)** | % of panellists rating this item as **1-3** | % of panellists rating this item as **4-6** | % of panellists rating this item as **7-9** |
|---|---|---|---|
| 5 (2, 7) | 30.9 | 36.4 | 32.7 |

Verbatim comments from Round 2:

- *‘Useful for trial reporting, but not intervention fidelity reporting’*
- *‘Study methods not fidelity’*
- *‘If applicable yes, but this is sort of obvious to include’*
- *‘Useful, and related to fidelity reporting (obviously), but not actually part of reporting the fidelity. (This is an outcome of fidelity measures, not the measures themselves)’*
- *‘Nearly always underpowered, so would omit moderator analysis requirements’*
- *‘Important but think this should come more in a SAP’*

- *‘This would be covered by the more general trial reporting relevant to what analyses were undertaken’*
- *‘Only if applicable based on a sound rationale in specific cases’*
- *‘Only important if fidelity results are additionally analyzed, which is not necessarily the case’*
- *‘This should certainly be reported, as significantly changes the interpretation of findings’*
- *‘Sensitivity analyses should be clearly planned a priori’*
- *‘This rationale is important’*

| | Not important | | | | | | Very important | | |
|---|---|---|---|---|---|---|---|---|---|
| | 1 | 2 | 3 | 4 | 5 | 6 | 7 | 8 | 9 |
| ITEM 66 | | | | | | | | | |

Please provide any comments or suggestions you have about ITEM 66.

**ITEM 67**

**ITEM 67:** If applicable, specify the rate of **missing intervention fidelity data*** and how it was managed. *Missing data refers to instances where intervention fidelity data that was planned to be collected was not actually obtained (e.g., participants or providers did not return fidelity checklists).

Ratings from Round 2:

| **Median (IQR)** | % of panellists rating this item as **1-3** | % of panellists rating this item as **4-6** | % of panellists rating this item as **7-9** |
|---|---|---|---|
| 7 (5.5, 8) | 7.3 | 32.7 | 60.0 |

Verbatim comments from Round 2:

- *‘Not ‘core’ info’*

- *‘For general reporting’*
- *‘I would say that it is important to show what fidelity is based upon (what % of data) and feel this level of detail is not particularly necessary’*
- *‘Depend on how you read this: Missing data although you attempted to measure data (data collection failure and its impact for assessing fidelity) or data you wanted to measure, but could not and the implications of not having that info. I now guess the first is intended here. You always have to report the validity/reliability of any assessment’*
- *‘Could be addressed by reporting the frequency of obtained fidelity measures, which shouldn't be too cumbersome’*
- *‘Not always important, but the 'if applicable' makes this high priority’*
- *‘Only required in case of a significant lack of fidelity data (the threshold of which would also need to be defined)’*
- *‘This is important but potentially a bit complicated to collect hence not core’*
- *‘Easy to do and important’*
- *‘Important concept’*

| | Not important | | | | | Very important | | | |
|---|---|---|---|---|---|---|---|---|---|
| | 1 | 2 | 3 | 4 | 5 | 6 | 7 | 8 | 9 |
| ITEM 67 | | | | | | | | | |

Please provide any comments or suggestions you have about ITEM 67.

**ITEM 68-69**

Please rate the importance of including the items below in a reporting guideline for intervention fidelity in non-drug, non-surgical trials.

Note: Some items are closely worded but their main substance is unique (highlighted in **bold**). Comments from Round 2 are reported verbatim, with some repeated across items as provided by the panellists.

**ITEM 68:** Specify what is the **sample of the intervention fidelity assessments** and how it was determined (e.g., whole trial sample, subset sample).

Ratings from Round 2:

| **Median (IQR)** | % of panellists rating this item as **1-3** | % of panellists rating this item as **4-6** | % of panellists rating this item as **7-9** |
|---|---|---|---|
| 7 (5, 8) | 16.4 | 21.8 | 61.8 |

Verbatim comments from Round 2:

- *‘May further our understanding of how fidelity measures are being captured, but probably is not critical’*
- *‘Only if applicable: if I assess all participants, the sample size is given by the sample size calculation for the trial, I don't need to explain that again for fidelity’*
- *‘I am assuming that the sample size was determined based on a power analysis to achieve statistical significance. If possible’*
- *‘Depends on what you measure and your goal’*
- *‘Seems important to correctly interpret reported fidelity assessments’*
- *‘Critical to know about the sample’*
- *‘Helps develop confidence in the outcome of the fidelity assessment’*
- *‘This item is relevant for understanding the origin and representativeness of fidelity data’*
- *‘Important to understand the reliability and validity of the assessment, and any possible bias affecting it’*

| | Not important | | | | | Very important | | | |
|---|---|---|---|---|---|---|---|---|---|
| | 1 | 2 | 3 | 4 | 5 | 6 | 7 | 8 | 9 |
| ITEM 68 | | | | | | | | | |

Please provide any comments or suggestions you have about ITEM 68.

**ITEM 69:** Describe any actions taken to minimise **selection bias when sampling a sub-set** of data points for intervention fidelity analysis.

Ratings from Round 2:

| **Median (IQR)** | % of panellists rating this item as **1-3** | % of panellists rating this item as **4-6** | % of panellists rating this item as **7-9** |
|---|---|---|---|
| 6 (5, 8) | 16.4 | 38.2 | 45.5 |

Verbatim comments from Round 2:

- *‘This is a "nice to have" rather than an essential reporting item for me’*
- *‘This is about conducting research according to the rules of good practice. Fidelity is no exception’*
- *‘Not a relevant detail for many studies but important if subset selected’*
- *‘If I understand correctly, a subset should be random, so that would be enough’*
- *‘Important and to be considered but maybe not in core fidelity reporting guidelines’*
- *‘This assumes random sampling is always appropriate and not sure it is’*
- *‘Important if using a subsample’*
- *‘The strategies for fidelity sampling should be briefly described’*
- *‘Critical to know about the sample’*
- *‘Important to understand the reliability and validity of the assessment, and any possible bias affecting it’*
- *‘This item is relevant for understanding the origin and representativeness of fidelity data’*

| | Not important | | | | | Very important | | | |
|---|---|---|---|---|---|---|---|---|---|
| | 1 | 2 | 3 | 4 | 5 | 6 | 7 | 8 | 9 |
| ITEM 69 | | | | | | | | | |

Please provide any comments or suggestions you have about ITEM 69.

**ITEM 70-72**

Please rate the importance of including the items below in a reporting guideline for intervention fidelity in non-drug, non-surgical trials.

Note: Some items are closely worded but their main substance is unique (highlighted in **bold**). Comments from Round 2 are reported verbatim, with some repeated across items as provided by the panellists.

**ITEM 70:** Specify the **number** of intervention fidelity assessors and indicate if they performed fidelity evaluations **independently** (without being influenced by one another).

Ratings from Round 2:

| **Median (IQR)** | % of panellists rating this item as **1-3** | % of panellists rating this item as **4-6** | % of panellists rating this item as **7-9** |
|---|---|---|---|
| 7 (6, 8) | 12.7 | 18.2 | 69.1 |

Verbatim comments from Round 2:

- *'Probably more important in highly specific study designs where this assessment should be rigorously documented'*
- *'Useful when there are multiple assessors and there seems to be variation among the assessors—not critical for all studies'*
- *'This is doing good research, Not typical for fidelity'*
- *'Important for understanding assessment'*
- *'Just as important as inter-rater assessments in quali studies and since fidelity is often quali and quant, then yes'*
- *'Important for transparency and being able to make a judgement about the validity of the fidelity assessment'*
- *'Pretty critical I would say, especially for reducing bias'*
- *'Reporting the number of assessors and their independence enhances transparency and allows future researchers to replicate rigorous fidelity assessment practices'*
- *'This is core'*

| | Not important | | | | | | | | Very important |
|---|---|---|---|---|---|---|---|---|---|
| | 1 | 2 | 3 | 4 | 5 | 6 | 7 | 8 | 9 |
| ITEM 70 | | | | | | | | | |

Please provide any comments or suggestions you have about ITEM 70.

**ITEM 71:** Specify the **level of blinding**, if any, of intervention fidelity assessors at the time of intervention fidelity assessments (e.g., blinded to intervention outcomes or intervention provider identity).

Ratings from Round 2:

| **Median (IQR)** | % of panellists rating this item as **1-3** | % of panellists rating this item as **4-6** | % of panellists rating this item as **7-9** |
|---|---|---|---|
| 6 (5, 7) | 9.1 | 43.6 | 47.3 |

Verbatim comments from Round 2:

- *'Good to report, but not critical'*
- *'Pretty critical I would say, especially for reducing bias'*
- *'This is challenging in some contexts'*
- *'It is useful to know how it was assessed, by who, and to what extent they were blinded, but blinding shouldn't be necessary and this may not be feasible in many non-drug/surgical trials'*
- *'Useful to know whether or not they were blinded, but need not be blinded'*
- *'This one is tricky - to measure fidelity, you need to know what was expected to be delivered - so you need to know whether it is the intervention arm or the control arm you are assessing. As such, while blinding sounds scientifically better, I am not convinced that is the case in this scenario'*
- *'This is probably just too hard to set as a generic reporting guideline'*
- *'Useful to know if blinded, but should not set the standard at being blinded'*
- *'This is essential'*

| | Not important | | | | | | Very important | | |
|---|---|---|---|---|---|---|---|---|---|
| | 1 | 2 | 3 | 4 | 5 | 6 | 7 | 8 | 9 |
| ITEM 71 | | | | | | | | | |

Please provide any comments or suggestions you have about ITEM 71.

#### SECTION 3 - Add Input

(Optional) If you have any further feedback, please share it here:

Powered by Qualtrics
