## Appendix 2 for "Establishing consensus on domains and items for Reporting intervention Fidelity in Non-Drug, non-surgical trials: the ReFiND Delphi study"

**ReFiND Consent**

### Welcome to the ReFiND Delphi Study Round 2

Dear ReFiND Collaborators,

Thank you for completing the ReFiND Delphi Round 1. We appreciate your time and contributions.

Your feeback contributed significantly to shaping Round 2. We have carefully reviewed all your comments and made adjustments to the survey structure, domains, and items accordingly.

Below, you will find important clarifications and guidance for the Round 2. Please review this information carefully before proceeding with the survey (tick the boxes once you have read the information).

**1. How were the Delphi domains and items developed?**

Domains and items presented in the Round 1 were extracted from a scoping review of existing recommendations on intervention fidelity (currently under peer review). We identified 73 documents (of various design types) that provided 81 recommendations on intervention fidelity. We then refined the domains and items identified (for reporting clarity purposes) and included them in the Round 1 survey.

Original recommendations from the literature were rarely developed with consensus methods and sometimes blend aspects of general trial or intervention reporting with intervention fidelity. Consequently, you may have noticed that some survey items seem broader than intervention fidelity. We decided not to exclude these items solely based on our own judgment. Since these items were reported in the literature as part of recommendations on intervention fidelity, we included them in the survey to ensure that the decision—whether to include or exclude these items— is made through your ratings (i.e., international consensus), rather than the judgment of a select group of authors. Our instruction is that if you perceive an item as more related to trial or general intervention reporting (rather than intervention fidelity), it should be voted out as it falls outside the scope of the ReFiND.

The specific sources of the domains and items are not displayed in the survey to avoid influencing your ratings, which should be based on the relevance of each domain/item and your expertise.

☐ I have read the information above.

**2. Relationship between ReFiND and other reporting guidelines**

ReFiND will serve as a standalone reporting guideline for intervention fidelity. We will encourage researchers to use CONSORT for general trial reporting, TIDieR for general intervention reporting, and ReFiND for intervention fidelity reporting. TIDieR authors suggest using TIDieR alongside CONSORT for general intervention reporting items, and we will similarly recommend that researchers using TIDieR refer to ReFiND for intervention fidelity reporting items.

While ReFiND focuses on trial reporting, the agreed domains and items may also apply to intervention protocol reporting. During the consensus meeting, we will discuss their relevance for protocols, and if consensus is reached, we will encourage their incorporation into protocol reporting (specifically for intervention fidelity), alongside SPIRIT. For now, please rate the domains and items based on their relevance for intervention fidelity reporting, not where they will be published.

☐ I have read the information above.

**3. Difference between reporting and practice guidelines**

ReFiND is a reporting guideline. Reporting guidelines focus on what should be reported in trials and/or how it should be reported. In contrast, practice guidelines focus on how to carry out specific actions. Therefore, practice ('how-to') guidelines are outside the scope of this guideline. If you feel that an item in the survey resembles a practice guideline rather than a reporting guideline, you are encouraged to give it a low rating.
Examples:

- Describe how intervention fidelity was assessed. (Reporting guideline)
- Assess intervention fidelity using validated measures. (Practice guideline)
- Report the extent to which an intervention was delivered as planned. (Reporting guideline)
- Ensure an intervention is delivered as planned. (Practice guideline)

☐ I have read the information above.

**4. Applicability to different trials and interventions**

We received a few suggestions to include a ‘if relevant’ option to domains/items, which is a valid consideration. However, adding an 'if relevant' option to all domains/items at this stage might suggest that every domain and item is important, provided it is applicable. We may consider adding this feature later, once we have agreed on the core domains and items. For now, please consider the following:

- We understand that is almost impossible an item to be applicable to all types of trials and non-drug, non-surgical interventions. However, some items will be more applicable than others. We ask you to use your judgement to decide whether it is worth to include certain domains and items based on their perceived applicability. For example, some items about intervention providers may not be applicable to self-management digital interventions. However, if we consider the range of other types of interventions, it can be still worthwhile to include some items that will not be intuitively applied to those interventions. As with other reporting guidelines, once we have agreed on a core set, we will encourage authors to use only those items that are applicable to their specific context and disregard those that are not.

☐ I have read the information above.

**5. Word/space limitations**

Some comments highlighted concerns about word and space limitations in trial reporting, which we acknowledge and appreciate. For this Delphi survey, we kindly ask that the rating of **essential** domains and items for intervention fidelity reporting focus on their importance, rather than on these limitations. Recommendations on how and where to best report items deemed essential will be addressed after Round 3.

☐ I have read the information above.

**Changes to the survey based on Round 1 feedback**

- We have adjusted the wording, merged certain items, and split others, resulting in a total of 72 items for Round 2.
- A summary of ratings and comments from Round 1 is presented along with the domains and items in Round 2. This summary is not provided for new items or those extensively changed in response to Round 1 feedback (e.g., when the substance of the domain/item was modified or a significant component was added).
- In Round 2, the full list of survey items is available at the bottom of each survey page for cross-checking if needed.

#### Instructions - ROUND 2

This Round 2 survey will take approximately 45 to 75 minutes to complete, though the duration may vary. The purpose of this study is to achieve consensus on a core set of domains and items to be included in a reporting guideline for intervention fidelity. For this study, *intervention fidelity* refers to the extent to which an intervention is implemented as planned in the trial protocol.

The survey consists of two sections:

**SECTION 1 - Rating Domains**
In this section, we will present broad domains of intervention fidelity. These domains represent overarching categories or areas related to intervention fidelity, identified through a scoping review. We will ask you to rate the importance of each domain based on its importance to fidelity evaluation and reporting.

**SECTION 2 - Rating Individual Items**
In this section, we will ask you to rate the importance of specific, individual intervention fidelity recommendation items for inclusion in the reporting guideline.

Ratings will be made using a 9-point scale (1 = not important to 9 = very important), similar to Round 1 (further details on the next page).

A key principle of the Delphi process is that participants rate domains and items based on

feedback from previous rounds. Therefore, all domains and items, including those that met the Round 1 inclusion criteria (>70% ratings 7-9), will be re-evaluated in Round 2.
Each domain and item includes a summary of Round 1 ratings and comments—please review this information before providing your ratings and use the free-text boxes to explain your rationale (optional).

You can pause the survey at any time by closing your browser and resume later, as long as you use the same browser where you started. Please note, however, that you may not be able to return to previous pages once you have submitted your responses.

This second Delphi Round will remain open until **19 December 2024**. We will contact you for the third round in late January 2025. The success of this reporting guideline in driving positive changes in fidelity and trial reporting relies on your participation, so we kindly ask you to complete all rounds. Thank you again for your collaboration.

Please rate the importance of the domains below for intervention fidelity evaluation and/or reporting:

**Domain 1: Intervention Design**
Description: This domain covers key elements of the intervention design pertinent for intervention fidelity. It may assess how well the conceptual and/or theoretical models underlying the intervention are reflected in the actual intervention execution within a trial. For example, if an intervention is based on a health behaviour change model, it assesses whether the model's components (e.g., motivation enhancement, self-efficacy building) are integrated into the intervention provider training, intervention delivery, intervention fidelity measures, etc.

Ratings from Round 1:

| **Median (IQR)** | % of panellists rating this item as **1-3** | % of panellists rating this item as **4-6** | % of panellists rating this item as **7-9** |
|---|---|---|---|
| 8 (IQR 7, 9) | 1.6 | 14.8 | **83.6** |

Comments on the ratings from panellists in Round 1:

- *'This is important but not essential, in the reporting of intervention fidelity as its about what was planned conceptually, and what is being measured in the trial - not therefore actually reporting intervention fidelity'*
- *'I believe this is important but not critical, based on the definition of intervention fidelity. A theory may not be fully operationalised within the protocol or training. However, intervention fidelity is defined as the extent to which an intervention aligns with its protocol, rather than the underlying theory'*
- *'Important for understanding the key components required to measure fidelity'*
- *'I cannot see how you can evaluate fidelity without considering the theoretical underpinnings of an intervention'*
- *'Conceptual model helps to establish a clear, structured framework for how an intervention should be delivered. It also provide explanation about the mechanisms driving behaviour change'*

The following questions use a slider. You can select your response by dragging the slider or clicking near your desired number.

If you do not want to use your mouse to navigate the survey, use the Tab key to move forward through questions and the Arrow keys to select options.

| | Not important | | | | | | Very important | | |
|---|---|---|---|---|---|---|---|---|---|
| | 1 | 2 | 3 | 4 | 5 | 6 | 7 | 8 | 9 |
| D1: Intervention Design | | | | | | | | | |

Please provide any comments or suggestions you have about Domain 1.

**Domain 2: Provider Training**
Description: This domain focuses on how intervention providers were trained to deliver the intervention with adequate fidelity. It may involve assessing and reporting whether intervention providers received the required training as outlined in the protocol and their competence and proficiency to deliver the intervention.

Ratings from Round 1:

| **Median (IQR)** | % of panellists rating this item as **1-3** | % of panellists rating this item as **4-6** | % of panellists rating this item as **7-9** |
|---|---|---|---|
| 8 (IQR 7, 9) | 1.6 | 21.3 | **77.0** |

Comments on the ratings from panellists in Round 1:

- *‘This is important but not essential, in the reporting of intervention fidelity as its about provider selection and training and perhaps performance assessment - not therefore actually reporting intervention fidelity’*
- *‘This domain is important for implementation  than intervention fidelity’*
- *‘I think training the providers also fits within Domain 3 (intervention delivery). However, I believe it’s important to have a standalone domain for provider training, as it is often overlooked’*
- *‘I believe this is important, but not critical. First, it does not apply equally to all interventions, as interventions may vary depending on the need for training (e.g., digital interventions). Second, assessing provider training overlaps with the concept of delivery’*
- *‘Training is critical because this is the point where 'drift' from the protocol standards may occur’*
- *‘Unless one knows if the provider had the required skills at the end of training it is impossible to see what can be attributed to what’*

- *'It would be important to understand, assess and clearly define what would be acceptable performance standards in terms of training. It is also important to have different perspectives depending on the level of the intervention and unit of inference (e.g. inference at the cluster level or at the individual level)'*
- *'Also important to assess how well providers have been trained (i.e. not just 'have they received training yes/no', but was the training in sufficient depth and making sure the training doesn't assume a provider to already have a baseline knowledge of intervention components specified in the protocol)'*

| | Not important | | | | | | Very important | | |
|---|---|---|---|---|---|---|---|---|---|
| | 1 | 2 | 3 | 4 | 5 | 6 | 7 | 8 | 9 |
| D2: Provider Training | | | | | | | | | |

Please provide any comments or suggestions you have about Domain 2.

**Domain 3: Intervention Delivery by Providers**
Description: This domain focuses on how intervention providers delivered the planned intervention. It may assess the extent to which intervention providers delivered the intervention as planned, including adherence to its core components, content, and dose, as well as avoidance of non-protocolised components.

Ratings from Round 1:

| **Median (IQR)** | % of panellists rating this item as **1-3** | % of panellists rating this item as **4-6** | % of panellists rating this item as **7-9** |
|---|---|---|---|
| 8 (IQR 8, 9) | 0.0 | 3.3 | **96.7** |

Comments on the ratings from panellists in Round 1:

- *'This is critical to the reporting of intervention fidelity in a RCT paper'*
- *'This is the core area to be evaluated as an intervention fidelity'*

- *'Recent systematic reviews on fidelity assessments illustrate that this  is the most common fidelity domain assessed in behavior interventions. compare to others, this domain can be assessed using several methods in low cost'*
- *'While the delivery by providers strengthens the effectiveness and reliability of interventions, the rigid adherence to protocols can pose barriers that limit the potential for necessary adaptations. Balancing protocol fidelity with the flexibility to adjust to participant needs is essential for optimizing both provider delivery and intervention outcomes'*

| | Not important | | | | | | Very important | | |
|---|---|---|---|---|---|---|---|---|---|
| | 1 | 2 | 3 | 4 | 5 | 6 | 7 | 8 | 9 |
| D3: Intervention Delivery by Providers | | | | | | | | | |

Please provide any comments or suggestions you have about Domain 3.

**Domain 4: Intervention Receipt by Participants**
Description: This domain focuses on two concepts: how intervention participants (a) received (actual receipt) and (b) understood the intervention (understanding).

(4a) Actual receipt: It may assess the extent to which participants received the intervention as planned. For example, in a group intervention where health professionals deliver weekly lectures on diabetes management, it assesses whether participants attended lectures (i.e., received the intervention).

(4b) Understanding: It may assess whether participants understood the instructions, concepts, or content of the intervention as outlined by the intervention's principles. This may include checking whether they can accurately repeat what was taught/delivered.

ATTENTION: Based on feedback from Round 1, this domain has two components (4a and 4b), which are rated independently. If you do NOT agree that Domain 4, 'Intervention Receipt by Participants', should be included in the reporting guideline, you should give a low score to both components (4a and 4b). If either component meets the threshold for inclusion consensus (>70% of ratings 7-9), Domain 4 will be recommended for inclusion in the guideline.

Ratings from Round 1:

Not applicable because this domain has changed significantly from the round 1.

Comments on the ratings from panellists in Round 1:

Not applicable because this domain has changed significantly from the round 1.

| | Not important | | | | | | Very important | | |
|---|---|---|---|---|---|---|---|---|---|
| | 1 | 2 | 3 | 4 | 5 | 6 | 7 | 8 | 9 |
| 4a: Actual receipt | | | | | | | | | |
| 4b: Understanding | | | | | | | | | |

Please provide any comments or suggestions you have about Domain 4.

**Domain 5: Intervention Enactment by Participants**
Description: This domain focuses on how intervention participants enacted the intervention. It may assess the extent to which participants integrate what they learned from the intervention into their routine behaviours. For example, it may assess whether intervention participants who received a lecture on preparing low-carbohydrate meals for diabetes management incorporated these preparations into their daily routines.

Ratings from Round 1:

| **Median (IQR)** | % of panellists rating this item as **1-3** | % of panellists rating this item as **4-6** | % of panellists rating this item as **7-9** |
|---|---|---|---|
| 7 (IQR 6, 8) | 6.6 | 29.5 | 63.9 |

Comments on the ratings from panellists in Round 1:

- *‘I know some frameworks include enactment but I personally do not think this is part of fidelity. This is an outcome of the intervention. And it is hard to differentiate this from participant adherence. In most cases, to me, they are the same. So, it is relevant for the trial but not part of fidelity. I would leave this out’*
- *‘Again this is a study outcome, not a process assessment. Fidelity is how well the study was delivered as designed - the process of intervention delivery. What the participants "do" with that education or training is the study outcome, not the study process’*
- *‘Part of NIH-BCC framework, but not 100% convinced - this feels more like a consequence of fidelity than fidelity itself’*
- *‘I think somehow we need a way to acknowledge how this may often overlap with study outcomes....as in if the main outcomes of the study are behavioural, then they might already assess enactment’*
- *‘same feedback as for the previous domain - this is not as important as the previous domain, and mixes aspects of intervention fidelity with participant adherence. Adherence to the allocated treatment is a different domain than intervention fidelity. Exploring whether participants enact the intervention, or embed in into routines, is a whole additional array of complexity - that most trials will not be able to explore well nor report meaningfully’*
- *‘Delivery, receipt and enactment are all crucial for fidelity measurement and reporting as they help us understand in detail the mechanism of impact and where fidelity may break down (eg b/c it was not conveyed to deliverers or delivered properly, it was not understood by intervention targets, or it was delivered and understood but not enacted). I rate enactment as more important than the delivery and receipt items above is we often have limited resources for fidelity assessment and enactment is ultimately what's needed for establishing fidelity.’*

- *‘The approach of evaluating how trial participants enact interventions presents both significant strengths and notable weaknesses. One of the key strengths lies in its ability to provide practical insights into real-world behavior changes. By focusing on how participants integrate what they learn into their daily routines, researchers gain a deeper understanding of the long-term impact of the intervention. This focus on behavioral integration not only highlights the effectiveness of the intervention but also allows for tailored feedback on which aspects resonate most with participants. However, this approach is not without its challenges. One major weakness is the potential for self-reporting bias, as participants may overstate their adherence to the intervention or misrepresent their behavior changes. Additionally, the narrow focus on individual enactment may overlook broader contextual factors that can influence outcomes, such as social support or environmental barriers. Furthermore, measuring enactment and adherence to specific recommendations can be complex, often requiring extensive and potentially cumbersome data collection methods. In summary, while evaluating participants' enactment of interventions offers valuable insights into behavioral change, it also presents challenges that need to be addressed to ensure accurate and comprehensive assessments’*

| | Not important | | | | | | Very important | | |
|---|---|---|---|---|---|---|---|---|---|
| | 1 | 2 | 3 | 4 | 5 | 6 | 7 | 8 | 9 |
| D5: Intervention Enactment by Participants | | | | | | | | | |

Please provide any comments or suggestions you have about Domain 5.

**Domain 6: Moderators of intervention fidelity and outcomes**
Description: This domain focuses on aspects that may moderate intervention fidelity and the relationship between intervention fidelity and trial outcomes. It may assess whether contextual elements, such as the provider-participant relationship or organisational factors (e.g., provider caseload), impact intervention fidelity. It may also assess whether intervention fidelity itself influences trial outcomes, and how

the outcomes of an intervention may be interpreted in the context of its fidelity results.

Ratings from Round 1:

| **Median (IQR)** | % of panellists rating this item as **1-3** | % of panellists rating this item as **4-6** | % of panellists rating this item as **7-9** |
|---|---|---|---|
| 7 (IQR 5, 9) | 11.5 | 32.8 | 55.7 |

Comments on the ratings from panellists in Round 1:

- *‘This is important, but is distinct from fidelity’*
- *‘The concept of "moderators" may not always be clear and straightforward’*
- *‘I think of provider-participant relationship and other 'non-specific factors' as being a part of intervention fidelity and should be considered carefully in the intervention design. That feels distinct from moderators. The context or setting of the intervention should also be specified in the design and therefor included in fidelity. If you are thinking about true moderators of trial outcomes, that seems more related to study design than intervention fidelity’*
- *‘While it can be relavant in , I think its relavant vary similar to what i described in the 'Provider Selection and Training' section. While it can be relevant, in many cases it can be difficult to capture and if planning for an large scale delivery in the future, it may give some information, how rigid the intervention components are and encourage designers to think about the generaliability of intervention’*
- *‘I do not think this should belong to the fidelity reporting guideline. This is part of secondary analyses of trials, similar to moderators of treatment effects’*
- *‘Typically analyses of these moderators are very underpowered, so not sure how important this is practically even if it is theoretically’*
- *‘I feel like there isn't yet enough consensus about what the moderators of fidelity are to include this, as in I think it would be hard for authors to know what they are being asked to report on, but I do think it's important to include’*
- *‘Extremely important as indicated higher, especially in hybrid implementation trials to differentiate between intervention and/or implementation effectiveness’*

| | Not important | | | | | | Very important | | |
|---|---|---|---|---|---|---|---|---|---|
| | 1 | 2 | 3 | 4 | 5 | 6 | 7 | 8 | 9 |
| D6: Moderators of intervention fidelity and outcomes | | | | | | | | | |

Please provide any comments or suggestions you have about Domain 6.

**Domain 7: Intervention fidelity measurement methods**
Description: This domain focuses on describing the methods used to assess and/or monitor intervention fidelity across its agreed domains. It may include reporting on the development of intervention fidelity measures, including evaluation of their psychometric properties, when applicable. It may also feature what constitutes acceptable levels of intervention fidelity, who the intervention fidelity assessors are and the sample size for intervention fidelity assessments.

Ratings from Round 1:

| **Median (IQR)** | % of panellists rating this item as **1-3** | % of panellists rating this item as **4-6** | % of panellists rating this item as **7-9** |
|---|---|---|---|
| 8 (IQR 7, 9) | 0.0 | 6.6 | **93.4** |

Comments on the ratings from panellists in Round 1:

- *‘Important consideration for reporting, but not a domain of fidelity’*
- *‘This domain is important primarily for intervention fidelity reporting. Since you are asking about this in addition to fidelity evaluation, I have marked it high’*
- *‘This is essential’*
- *‘This feels core to me, ie the trial must report how it defines and measures intervention fidelity and justify that’*
- *‘This is an important aspects of intervention fidelity in order not to misinform the intervention designers and policy makers that the success/failure of an intervention is due to or not due to its fidelity’*

- *‘This component is critical. Without confidence in the methods used to assess fidelity, we cannot determine whether the intervention was delivered as intended’*

| | Not important | | | | | | | Very important | |
|---|---|---|---|---|---|---|---|---|---|
| | 1 | 2 | 3 | 4 | 5 | 6 | 7 | 8 | 9 |
| D7: Intervention fidelity measurement methods | | | | | | | | | |

**ITEM 1: Describe a method** to assess the extent to which providers delivered the intervention as planned in the intervention protocol (i.e., fidelity of intervention delivery).

Ratings from Round 1:

| **Median (IQR)** | % of panellists rating this item as **1-3** | % of panellists rating this item as **4-6** | % of panellists rating this item as **7-9** |
|---|---|---|---|
| 9 (8, 9) | 0.0 | 6.6 | **93.4** |

Comments on the ratings from panellists in Round 1:

None provided.

| | Not important | | | | | | Very important | | |
|---|---|---|---|---|---|---|---|---|---|
| | 1 | 2 | 3 | 4 | 5 | 6 | 7 | 8 | 9 |
| ITEM 1 | | | | | | | | | |

Please provide any comments or suggestions you have about ITEM 1.

**ITEM 2:** Describe the **intervention content** (i.e., the 'what') planned to be delivered in the trial and report whether it was actually delivered as planned.

Ratings from Round 1:

| **Median (IQR)** | % of panellists rating this item as **1-3** | % of panellists rating this item as **4-6** | % of panellists rating this item as **7-9** |
|---|---|---|---|
| 9 (8, 9) | 1.6 | 3.3 | **95.1** |

Comments on the ratings from panellists in Round 1:

- *'Seems critical'*

| | Not important | | | | | Very important | | | |
|---|---|---|---|---|---|---|---|---|---|
| | 1 | 2 | 3 | 4 | 5 | 6 | 7 | 8 | 9 |
| ITEM 2 | | | | | | | | | |

Please provide any comments or suggestions you have about ITEM 2.

**ITEM 3:** Specify the planned **intervention dose** for each intervention arm, including the comparison/control intervention arm, and report whether it was actually delivered as planned.

Ratings from Round 1:

| **Median (IQR)** | % of panellists rating this item as **1-3** | % of panellists rating this item as **4-6** | % of panellists rating this item as **7-9** |
|---|---|---|---|
| 9 (8, 9) | 0.0 | 4.9 | **95.1** |

Comments on the ratings from panellists in Round 1:

- *'This is really important. It is relevant to report the fidelity in the comparison/control arm as well'*
- *'This should be the minimal requirement for a fidelity assessment'*

| | Not important | | | | | Very important | | | |
|---|---|---|---|---|---|---|---|---|---|
| | 1 | 2 | 3 | 4 | 5 | 6 | 7 | 8 | 9 |
| ITEM 3 | | | | | | | | | |

Please provide any comments or suggestions you have about ITEM 3.

**ITEM 4:** Describe the planned **intervention processes** (e.g., delivery mode—in-person/digital, setting—clinic/home/school, format—individual/group) and indicate whether they were implemented as planned.

Ratings from Round 1:

| **Median (IQR)** | % of panellists rating this item as **1-3** | % of panellists rating this item as **4-6** | % of panellists rating this item as **7-9** |
|---|---|---|---|
| 9 (8, 9) | 1.6 | 6.6 | **91.8** |

Comments on the ratings from panellists in Round 1:

- *'Not fidelity per se? This is more form rather than function'*
- *'I think this will be included under intervention description'*
- *'Clearly describing the intervention is essential for providing a clear account of fidelity'*
- *'Important info for replication studies'*

| | Not important | | | | | | Very important | | |
|---|---|---|---|---|---|---|---|---|---|
| | 1 | 2 | 3 | 4 | 5 | 6 | 7 | 8 | 9 |
| ITEM 4 | | | | | | | | | |

Please provide any comments or suggestions you have about ITEM 4.

**ITEM 5:** Report the degree to which the intervention delivered to intervention participants is **congruent with its underlying theory or logic model**.

Ratings from Round 1:

| **Median (IQR)** | % of panellists rating this item as **1-3** | % of panellists rating this item as **4-6** | % of panellists rating this item as **7-9** |
|---|---|---|---|
| 7 (5, 9) | 9.8 | 27.9 | 62.3 |

Comments on the ratings from panellists in Round 1:

- *'I am conflicted by this one. I feel it is good to do this but am reluctant to have this as a core requirement. Complex interventions may have manifold pathways of intervention effect for which it may be unproductive to have a complete mapping against the theory as opposed to mapping against what are considered to be the key mechanistic pathway(s)'*
- *'The theoretical or logic model should be part of a satellite intervention, not the full trial reporting'*
- *'Cannot really assess fidelity without this (fidelity to what?)'*
- *'The underpinning theory/logic model is so often not reported, so this is really important, and without this there could be knock-on impacts on fidelity (e.g., without this we don't know what the 'essential components' are, and therefore whayt providers should be trained in). It also aids the development of fidelity checklists.'*

| | Not important | | | | | Very important | | | |
|---|---|---|---|---|---|---|---|---|---|
| | 1 | 2 | 3 | 4 | 5 | 6 | 7 | 8 | 9 |
| ITEM 5 | | | | | | | | | |

Please provide any comments or suggestions you have about ITEM 5.

**ITEM 6:** Describe a **method to assess** whether unintended components are delivered (e.g., components that are unnecessary or unhelpful).

Ratings from Round 1:

| **Median (IQR)** | % of panellists rating this item as **1-3** | % of panellists rating this item as **4-6** | % of panellists rating this item as **7-9** |
|---|---|---|---|
| 7 (5, 9) | 8.2 | 31.1 | 60.7 |

Comments on the ratings from panellists in Round 1:

- *'The delivery of key components, to me, is more important than the fact that there might be 'unnecessary' (let's call them 'not key') components being delivered as well'*
- *'Important - but depends on the intervention design explicitly stating what prohibited components may be? This is quite hard to assess in practice e.g. in situations where providers are using their own clinical judgement too to adapt interventions (what criteria would be used to evaluate whether these were prohibited?'*
- *'This is useful when possible but should not be a requirement'*
- *'To measure fidelity, you first need a clear description of the intervention and its processes including proscribed components'*
- *'Yes! Conflicting advice is a big issue impacting adherence'*
- *'This is an essential aspect of intervention fidelity'*

| | Not important | | | | | Very important | | | |
|---|---|---|---|---|---|---|---|---|---|
| | 1 | 2 | 3 | 4 | 5 | 6 | 7 | 8 | 9 |
| ITEM 6 | | | | | | | | | |

Please provide any comments or suggestions you have about ITEM 6.

**ITEM 7: Report whether** unintended components (e.g., components that are unnecessary or unhelpful) were delivered.

Ratings from Round 1:

| **Median (IQR)** | % of panellists rating this item as **1-3** | % of panellists rating this item as **4-6** | % of panellists rating this item as **7-9** |
|---|---|---|---|
| 7 (5, 9) | 8.2 | 34.4 | 57.4 |

Comments on the ratings from panellists in Round 1:

- *‘The delivery of key components, to me, is more important than the fact that there might be 'unnecessary' (let's call them 'not key') components being delivered as well’*
- *‘Important - but depends on the intervention design explicitly stating what prohibited components may be? This is quite hard to assess in practice e.g. in situations where providers are using their own clinical judgement too to adapt interventions (what criteria would be used to evaluate whether these were prohibited?’*
- *‘This is useful when possible but should not be a requirement’*
- *‘To measure fidelity, you first need a clear description of the intervention and its processes including proscribed components’*
- *‘Yes! Conflicting advice is a big issue impacting adherence’*
- *‘This is an essential aspect of intervention fidelity'*

| | Not important | | | | | | Very important | | |
|---|---|---|---|---|---|---|---|---|---|
| | 1 | 2 | 3 | 4 | 5 | 6 | 7 | 8 | 9 |
| ITEM 7 | | | | | | | | | |

Please provide any comments or suggestions you have about ITEM 7.

**ITEM 8:** Specify how delivery contamination **between intervention arms** was avoided.

Ratings from Round 1:

| **Median (IQR)** | % of panellists rating this item as **1-3** | % of panellists rating this item as **4-6** | % of panellists rating this item as **7-9** |
|---|---|---|---|
| 7 (5, 9) | 16.4 | 16.4 | 67.2 |

Comments on the ratings from panellists in Round 1:

- *'This is important for trial design, but not intervention fidelity measurement'*
- *'This is part of trial design, not treatment fidelity.*
- *'Not fidelity specific'*
- *'It is important to ensure the intended outcome can be attributed to the intervention'*
- *'Important for preventing contamination and thereby preserving fidelity'*

| | Not important | | | | | Very important | | | |
|---|---|---|---|---|---|---|---|---|---|
| | 1 | 2 | 3 | 4 | 5 | 6 | 7 | 8 | 9 |
| ITEM 8 | | | | | | | | | |

Please provide any comments or suggestions you have about ITEM 8.

**ITEM 9-18**

Please rate the importance of including the items below in a reporting guideline for intervention fidelity in non-drug, non-surgical trials.

Note: Some items are closely worded but their main substance is unique (highlighted in **bold**). Comments from Round 1 are reported verbatim, with some repeated across items as provided by the panellists.

**ITEM 9: Describe a method** to assess the degree to which participants **received** the intervention components as planned (e.g., being exposed to the intervention).

Ratings from Round 1:

Not applicable because this item has changed significantly from the round 1.

Comments on the ratings from panellists in Round 1:

Not applicable because this item has changed significantly from the round 1.

| | Not important | | | | | Very important | | | |
|---|---|---|---|---|---|---|---|---|---|
| | 1 | 2 | 3 | 4 | 5 | 6 | 7 | 8 | 9 |
| ITEM 9 | | | | | | | | | |

Please provide any comments or suggestions you have about ITEM 9.

**ITEM 10: Report** the degree to which participants **received** the planned intervention **content**.

Ratings from Round 1:

Not applicable because this item has changed significantly from the round 1.

Comments on the ratings from panellists in Round 1:

Not applicable because this item has changed significantly from the round 1.

| | Not important | | | | | Very important | | | |
|---|---|---|---|---|---|---|---|---|---|
| | 1 | 2 | 3 | 4 | 5 | 6 | 7 | 8 | 9 |
| ITEM 10 | | | | | | | | | |

Please provide any comments or suggestions you have about ITEM 10.

**ITEM 11: Report** the degree to which participants **received** the planned intervention **dose**.

Ratings from Round 1:

Not applicable because this item has changed significantly from the round 1.

Comments on the ratings from panellists in Round 1:

Not applicable because this item has changed significantly from the round 1.

| | Not important | | | | | Very important | | | |
|---|---|---|---|---|---|---|---|---|---|
| | 1 | 2 | 3 | 4 | 5 | 6 | 7 | 8 | 9 |
| ITEM 11 | | | | | | | | | |

Please provide any comments or suggestions you have about ITEM 11.

**ITEM 12: Describe a method** to assess the degree to which participants **understood/comprehended** the intervention components as planned.

Ratings from Round 1:

Not applicable because this item has changed significantly from the round 1.

Comments on the ratings from panellists in Round 1:

Not applicable because this item has changed significantly from the round 1.

| | Not important | | | | | Very important | | | |
|---|---|---|---|---|---|---|---|---|---|
| | 1 | 2 | 3 | 4 | 5 | 6 | 7 | 8 | 9 |
| ITEM 12 | | | | | | | | | |

Please provide any comments or suggestions you have about ITEM 12.

**ITEM 13: Report** the degree to which participants **understood/comprehended** the intervention components as planned.

Ratings from Round 1:

Not applicable because this item has changed significantly from the round 1.

Comments on the ratings from panellists in Round 1:

Not applicable because this item has changed significantly from the round 1.

| | Not important | | | | | Very important | | | |
|---|---|---|---|---|---|---|---|---|---|
| | 1 | 2 | 3 | 4 | 5 | 6 | 7 | 8 | 9 |
| ITEM 13 | | | | | | | | | |

Please provide any comments or suggestions you have about ITEM 13.

**ITEM 14: Describe a method** to assess the degree to which intervention participants **apply the intervention in real-life settings** (also referred to as 'enactment').

Ratings from Round 1:

| **Median (IQR)** | % of panellists rating this item as **1-3** | % of panellists rating this item as **4-6** | % of panellists rating this item as **7-9** |
|---|---|---|---|
| 6 (4, 8) | 21.3 | 36.1 | 42.6 |

Comments on the ratings from panellists in Round 1:

- *'This is study outcome, not fidelity'*

- *'Whether a participant applies the intervention principles to their settings is an outcome of the intervention. Not part of fidelity'*
- *'A moderator really'*
- *'What is delivered is important. What is enacted is less controllable and not so relevant for say a pragmatic trial'*
- *'This item may be relevant to some interventions, but not all, so maybe this item should not be part of the core components of treatment fidelity'*

| | Not important | | | | | Very important | | | |
|---|---|---|---|---|---|---|---|---|---|
| | 1 | 2 | 3 | 4 | 5 | 6 | 7 | 8 | 9 |
| ITEM 14 | | | | | | | | | |

Please provide any comments or suggestions you have about ITEM 14.

**ITEM 15: Report** how well intervention participants **enacted** the intervention **content**, meaning the degree to which they **applied it in real-life settings.**

Ratings from Round 1:

Not applicable because this item has changed significantly from the round 1.

Comments on the ratings from panellists in Round 1:

Not applicable because this item has changed significantly from the round 1.

| | Not important | | | | | Very important | | | |
|---|---|---|---|---|---|---|---|---|---|
| | 1 | 2 | 3 | 4 | 5 | 6 | 7 | 8 | 9 |
| ITEM 15 | | | | | | | | | |

Please provide any comments or suggestions you have about ITEM 15.

**ITEM 16: Report** how well intervention participants **enacted** the intervention **dose**, meaning the degree to which they **applied it in real-life settings**.

Ratings from Round 1:

Not applicable because this item has changed significantly from the round 1.

Comments on the ratings from panellists in Round 1:

Not applicable because this item has changed significantly from the round 1.

| | Not important | | | | | Very important | | | |
|---|---|---|---|---|---|---|---|---|---|
| | 1 | 2 | 3 | 4 | 5 | 6 | 7 | 8 | 9 |
| ITEM 16 | | | | | | | | | |

Please provide any comments or suggestions you have about ITEM 16.

**ITEM 17: Describe a method to assess participant adherence** to the intervention (i.e., degree to which the participant **follows the intervention as instructed/required**).

Ratings from Round 1:

Not applicable because this item has changed significantly from the round 1.

Comments on the ratings from panellists in Round 1:

Not applicable because this item has changed significantly from the round 1.

| | Not important | | | | | Very important | | | |
|---|---|---|---|---|---|---|---|---|---|
| | 1 | 2 | 3 | 4 | 5 | 6 | 7 | 8 | 9 |
| ITEM 17 | | | | | | | | | |

Please provide any comments or suggestions you have about ITEM 17.

**ITEM 18: Report participant adherence** to the intervention (i.e., degree to which the participant **follows the intervention as instructed/required**).

Ratings from Round 1:

Not applicable because this item has changed significantly from the round 1.

Comments on the ratings from panellists in Round 1:

Not applicable because this item has changed significantly from the round 1.

| | Not important | | | | | Very important | | | |
|---|---|---|---|---|---|---|---|---|---|
| | 1 | 2 | 3 | 4 | 5 | 6 | 7 | 8 | 9 |
| ITEM 18 | | | | | | | | | |

Please provide any comments or suggestions you have about ITEM 18.

**ITEM 19-22**

Please rate the importance of including the items below in a reporting guideline for intervention fidelity in non-drug, non-surgical trials.

Note: Some items are closely worded but their main substance is unique (highlighted in **bold**). Comments from Round 1 are reported verbatim, with some repeated across items as provided by the panellists.

**ITEM 19:** Describe a **method to assess** the extent to which intervention participants **accept** the intervention (e.g., their perceptions, beliefs, attitudes, and intentions to use the intervention).

Ratings from Round 1:

| **Median (IQR)** | % of panellists rating this item as **1-3** | % of panellists rating this item as **4-6** | % of panellists rating this item as **7-9** |
|---|---|---|---|
| 6 (3, 8) | 29.5 | 27.9 | 42.6 |

Comments on the ratings from panellists in Round 1:

- *'Will influence fidelity, but probably not part of it, though related to receipt so I gave it a 2'*
- *'I think acceptability is distinct from fidelity – linked outcome but distinct. It can fit under a moderator of fidelity. But it is a separate implementation outcome so wouldn't suggest it is an essential item in a fidelity reporting guideline per se'*
- *this is not intervention fidelity but rather intervention acceptability - a different domain/construct and should be part of the work-up of RCTs but not part of reporting guidance for intervention fidelity'*
- *'Buy-in is required. Implementation strategies need to be informed by this knowledge so they can be tailored to the end-users needs and overcome barriers to Implementation'*
- *'Important but is it critical to fidelity?'*

| | Not important | | | | | Very important | | | |
|---|---|---|---|---|---|---|---|---|---|
| | 1 | 2 | 3 | 4 | 5 | 6 | 7 | 8 | 9 |
| ITEM 19 | | | | | | | | | |

Please provide any comments or suggestions you have about ITEM 19.

**ITEM 20: Report** the extent to which intervention participants **accept** the intervention (e.g., their perceptions, beliefs, attitudes, and intentions to use the intervention).

Ratings from Round 1:

| **Median (IQR)** | % of panellists rating this item as **1-3** | % of panellists rating this item as **4-6** | % of panellists rating this item as **7-9** |
|---|---|---|---|
| 6 (3, 8) | 29.5 | 21.3 | 49.2 |

Comments on the ratings from panellists in Round 1:

- *'Will influence fidelity, but probably not part of it, though related to receipt so I gave it a 2'*
- *'I think acceptability is distinct from fidelity – linked outcome but distinct. It can fit under a moderator of fidelity. But it is a separate implementation outcome so wouldn't suggest it is an essential item in a fidelity reporting guideline per se'*
- *this is not intervention fidelity but rather intervention acceptability - a different domain/construct and should be part of the work-up of RCTs but not part of reporting guidance for intervention fidelity'*
- *'Buy-in is required. Implementation strategies need to be informed by this knowledge so they can be tailored to the end-users needs and overcome barriers to Implementation'*
- *'As a moderator - and like many moderators - it can change over time'*
- *'Important but is it critical to fidelity?'*

| | Not important | | | | | Very important | | | |
|---|---|---|---|---|---|---|---|---|---|
| | 1 | 2 | 3 | 4 | 5 | 6 | 7 | 8 | 9 |
| ITEM 20 | | | | | | | | | |

Please provide any comments or suggestions you have about ITEM 20.

**ITEM 21:** Report intervention **participants' characteristics** (e.g., age, gender, comorbid conditions, personality traits) that can influence their engagement with the intervention.

Ratings from Round 1:

| **Median (IQR)** | % of panellists rating this item as **1-3** | % of panellists rating this item as **4-6** | % of panellists rating this item as **7-9** |
|---|---|---|---|
| 6 (4, 8) | 24.6 | 34.4 | 41.0 |

Comments on the ratings from panellists in Round 1:

- *‘This is not part of fidelity. General reporting’*
- *‘This refers much more to the result analysis than to the fidelity report’*
- *‘Intervention should take account of these - again, these might act as a moderator of outcomes rather than being related to fidelity’*
- *‘Important as trials often recruit the more 'engaged' participants - helps us to know the extent to which the intervention could be scaled up in future’*
- *‘Important for differentiating between different levels of fidelity and engagement’*
- *This is important to see the contextual factors which facilitate/hinder the fidelity'*

| | Not important | | | | | Very important | | | |
|---|---|---|---|---|---|---|---|---|---|
| | 1 | 2 | 3 | 4 | 5 | 6 | 7 | 8 | 9 |
| ITEM 21 | | | | | | | | | |

Please provide any comments or suggestions you have about ITEM 21.

**ITEM 22:** Report **intervention participants’ satisfaction** with the intervention.

Ratings from Round 1:

| **Median (IQR)** | % of panellists | % of panellists | % of panellists |
|---|---|---|---|

| | rating this item as **1-3** | rating this item as **4-6** | rating this item as **7-9** |
|---|---|---|---|
| 6 (3, 7) | 34.3 | 31.1 | 34.4 |

Comments on the ratings from panellists in Round 1:

- *‘This is not intervention fidelity but rather intervention satisfaction - a different domain/construct and should be part of the outcomes in RCTs but not part of reporting guidance for intervention fidelity’*
- *‘This is a trial outcome, not fidelity assessment. Satisfaction, a very difficult variable to deal with’*
- *‘This is good to do, but adjacent to fidelity’*
- *‘Yet satisfaction is a domain having many items in it. Since it is not easy to assess satisfaction by a single item’*
- *‘Unhappy participants equals a risk to fidelity’*

| | Not important | | | | | Very important | | | |
|---|---|---|---|---|---|---|---|---|---|
| | 1 | 2 | 3 | 4 | 5 | 6 | 7 | 8 | 9 |
| ITEM 22 | | | | | | | | | |

Please provide any comments or suggestions you have about ITEM 22.

**ITEM 23-26**

Please rate the importance of including the items below in a reporting guideline for intervention fidelity in non-drug, non-surgical trials.

Note: Some items are closely worded but their main substance is unique (highlighted in **bold**). Comments from Round 1 are reported verbatim, with some repeated across items as provided by the panellists.

**ITEM 23:** Describe, from the trial outset, the **core components** of the intervention that are essential and must be delivered.

Ratings from Round 1:

| **Median (IQR)** | % of panellists rating this item as **1-3** | % of panellists rating this item as **4-6** | % of panellists rating this item as **7-9** |
|---|---|---|---|
| 9 (7, 9) | 6.6 | 8.2 | **85.2** |

Comments on the ratings from panellists in Round 1:

- *'This is really intervention description, but is a start point for fidelity assessment'*
- *'To measure fidelity, you first need a clear description of the intervention and its processes'*

| | Not important | | | | | | Very important | | |
|---|---|---|---|---|---|---|---|---|---|
| | 1 | 2 | 3 | 4 | 5 | 6 | 7 | 8 | 9 |
| ITEM 23 | | | | | | | | | |

Please provide any comments or suggestions you have about ITEM 23.

**ITEM 24:** Describe, from the trial outset, the **non-core components** where delivery is flexible.

Ratings from Round 1:

| **Median (IQR)** | % of panellists rating this item as **1-3** | % of panellists rating this item as **4-6** | % of panellists rating this item as **7-9** |
|---|---|---|---|
| 8 (7, 9) | 4.9 | 18.0 | **77.0** |

Comments on the ratings from panellists in Round 1:

- *‘This is important, but is part of the method description, not part of fidelity. (The part related to fidelity would be how the fidelity measure was built to address the adaptations.)’*
- *‘This is really intervention description, but is a start point for fidelity assessment’*
- *‘To measure fidelity, you first need a clear description of the intervention and its processes’*

| | Not important | | | | | Very important | | | |
|---|---|---|---|---|---|---|---|---|---|
| | 1 | 2 | 3 | 4 | 5 | 6 | 7 | 8 | 9 |
| ITEM 24 | | | | | | | | | |

Please provide any comments or suggestions you have about ITEM 24.

**ITEM 25:** Report whether **adaptations made** to the intervention are consistent with the flexibility defined in the protocol, if applicable.

Ratings from Round 1:

Not applicable because this item has changed significantly from the round 1.

Comments on the ratings from panellists in Round 1:

Not applicable because this item has changed significantly from the round 1.

| | Not important | | | | | Very important | | | |
|---|---|---|---|---|---|---|---|---|---|
| | 1 | 2 | 3 | 4 | 5 | 6 | 7 | 8 | 9 |
| ITEM 25 | | | | | | | | | |

Please provide any comments or suggestions you have about ITEM 25.

**ITEM 26:** When applicable, report how **intervention fidelity deviations** were addressed, including whether strategies to improve fidelity were used (e.g., increased provider supervision).

Ratings from Round 1:

| **Median (IQR)** | % of panellists rating this item as **1-3** | % of panellists rating this item as **4-6** | % of panellists rating this item as **7-9** |
|---|---|---|---|
| 7 (6, 9) | 3.3 | 23.0 | **73.8** |

Comments on the ratings from panellists in Round 1:

- *'This one is a bit trickier- might not always be appropriate to 'intervene' to deal with fidelity deviations, particularly depending on trial and its design, as sometimes this can be seen as changing/iterating the intervention as the trial progresses. And there are mixed views on this. So I have put as a mid-rating not because I don't think it is important, but because appreciate there are likely mixed views on this'*
- *'Fidelity deviations are part of usual care for any treatment - so pragmatic trials will be less concerned about this, than explanatory trials'*
- *'Essential. Including if they were not dealt with'*
- *'Important for replication studies'*

| | Not important | | | | | | Very important | | |
|---|---|---|---|---|---|---|---|---|---|
| | 1 | 2 | 3 | 4 | 5 | 6 | 7 | 8 | 9 |
| ITEM 26 | | | | | | | | | |

Please provide any comments or suggestions you have about ITEM 26.

**ITEM 27-34**

Please rate the importance of including the items below in a reporting guideline for intervention fidelity in non-drug, non-surgical trials.

Note: Some items are closely worded but their main substance is unique (highlighted in **bold**). Comments from Round 1 are reported verbatim, with some repeated across items as provided by the panellists.

**ITEM 27:** Report the extent to which the **actual intervention providers in the trial met the provider selection criteria or expected characteristics** described in the protocol (e.g., training, experience, qualifications).

Ratings from Round 1:

Not applicable because this item has changed significantly from the round 1.

Comments on the ratings from panellists in Round 1:

Not applicable because this item has changed significantly from the round 1.

| | Not important | | | | | Very important | | | |
|---|---|---|---|---|---|---|---|---|---|
| | 1 | 2 | 3 | 4 | 5 | 6 | 7 | 8 | 9 |
| ITEM 27 | | | | | | | | | |

Please provide any comments or suggestions you have about ITEM 27.

**ITEM 28: Describe a method** to assess **provider competence and proficiency** to deliver the intervention (e.g., assessment of knowledge and skills).

Ratings from Round 1:

| **Median (IQR)** | % of panellists rating this item as **1-3** | % of panellists rating this item as **4-6** | % of panellists rating this item as **7-9** |
|---|---|---|---|
| 7 (5, 8) | 6.6 | 36.1 | 57.4 |

Comments on the ratings from panellists in Round 1:

- *'This is a tricky one, as high fidelity doesn't necessarily = more skilful, as think it depends on the quality of the intervention components that the provider is asked to deliver too. If this can be judged in other ways than fidelity then think this could be interesting'*
- *'This is not a fidelity assessment'*
- *'Competence assessments are controversial in RCTs, particularly pragmatic embedded RCTs where the interventions are designed to be delivered by healthcare providers that would typically offer the care to patients. Explanatory trials may need more information on these aspects than pragmatic trials'*
- *'May be particularly difficult to assess'*
- *'Important, with a focus on using existing valid tools, if applicable. We have too many non-evaluated methods'*

| | Not important | | | | | Very important | | | |
|---|---|---|---|---|---|---|---|---|---|
| | 1 | 2 | 3 | 4 | 5 | 6 | 7 | 8 | 9 |
| ITEM 28 | | | | | | | | | |

Please provide any comments or suggestions you have about ITEM 28.

**ITEM 29: Report provider competence and proficiency** to deliver the intervention (e.g., comprehension of the intervention components, demonstration of relevant skills required to deliver the intervention, etc).

Ratings from Round 1:

| **Median (IQR)** | % of panellists rating this item as **1-3** | % of panellists rating this item as **4-6** | % of panellists rating this item as **7-9** |
|---|---|---|---|
| 7 (6, 8) | 4.9 | 29.5 | 65.6 |

Comments on the ratings from panellists in Round 1:

- *'This is a tricky one, as high fidelity doesn't necessarily = more skilful, as think it depends on the quality of the intervention components that the provider is asked to deliver too. If this can be judged in other ways than fidelity then think this could be interesting'*
- *'This is not a fidelity assessment'*
- *'Competence assessments are controversial in RCTs, particularly pragmatic embedded RCTs where the interventions are designed to be delivered by healthcare providers that would typically offer the care to patients. Explanatory trials may need more information on these aspects than pragmatic trials'*
- *'May be particularly difficult to assess'*

- *'Important, with a focus on using existing valid tools, if applicable. We have too many non-evaluated methods'*

| | Not important | | | | | Very important | | | |
|---|---|---|---|---|---|---|---|---|---|
| | 1 | 2 | 3 | 4 | 5 | 6 | 7 | 8 | 9 |
| ITEM 29 | | | | | | | | | |

Please provide any comments or suggestions you have about ITEM 29.

**ITEM 30:** Describe an **intervention provider training plan** including training content, amount, and methods used to deliver the training (e.g, training manuals, didactic sessions, role modelling, supervision with feedback).

Ratings from Round 1:

| **Median (IQR)** | % of panellists rating this item as **1-3** | % of panellists rating this item as **4-6** | % of panellists rating this item as **7-9** |
|---|---|---|---|
| 7 (6, 9) | 3.3 | 24.6 | **72.1** |

Comments on the ratings from panellists in Round 1:

- *'This is not focused on intervention fidelity'*
- *'Useful for identifying why fidelity levels achieved, but not fidelity per se'*
- *'Important for strengthening and assessing fidelity (of delivery)'*
- *'This is just good research practice to ensure quality of delivery and to identify where improvement could be made - especially for adaptation prior to scaleup or modification during implementation'*

| | Not important | | | | | Very important | | | |
|---|---|---|---|---|---|---|---|---|---|
| | 1 | 2 | 3 | 4 | 5 | 6 | 7 | 8 | 9 |
| ITEM 30 | | | | | | | | | |

Please provide any comments or suggestions you have about ITEM 30.

**ITEM 31:** Report whether intervention **provider training was delivered as planned**, including content, amount, and methods of training, as well as the reasons for any discrepancy.

Ratings from Round 1:

| **Median (IQR)** | % of panellists rating this item as **1-3** | % of panellists rating this item as **4-6** | % of panellists rating this item as **7-9** |
|---|---|---|---|
| 8 (7, 9) | 1.6 | 19.7 | **78.7** |

Comments on the ratings from panellists in Round 1:

- *'Only important for certain kinds of studies'*
- *'Depends on mode of delivery of intervention'*
- *'This seems key. There are so many treatment variations that even defining 'exercise' is valuable'*

- *'Essential'*

| | Not important | | | | | Very important | | | |
|---|---|---|---|---|---|---|---|---|---|
| | 1 | 2 | 3 | 4 | 5 | 6 | 7 | 8 | 9 |
| ITEM 31 | | | | | | | | | |

Please provide any comments or suggestions you have about ITEM 31.

**ITEM 32:** Report the level of training **received by intervention providers** (e.g., frequency of training, attendance to training sessions).

Ratings from Round 1:

| **Median (IQR)** | % of panellists rating this item as **1-3** | % of panellists rating this item as **4-6** | % of panellists rating this item as **7-9** |
|---|---|---|---|
| 7 (6, 8) | 6.6 | 39.3 | 54.1 |

Comments on the ratings from panellists in Round 1:

- *'It is important to ensure that the training was performed, but it is not necessary to provide this detailed information'*
- *'Less important than learning, unless the study addresses these variables as part of mediator for DV based on existing hypotheses'*

| | Not important | | | | | Very important | | | |
|---|---|---|---|---|---|---|---|---|---|
| | 1 | 2 | 3 | 4 | 5 | 6 | 7 | 8 | 9 |
| ITEM 32 | | | | | | | | | |

Please provide any comments or suggestions you have about ITEM 32.

**ITEM 33:** Report whether **booster (i.e., additional) training sessions** were offered and delivered to intervention providers.

Ratings from Round 1:

| **Median (IQR)** | % of panellists rating this item as **1-3** | % of panellists rating this item as **4-6** | % of panellists rating this item as **7-9** |
|---|---|---|---|
| 7 (6, 8) | 4.9 | 27.9 | 67.2 |

Comments on the ratings from panellists in Round 1:

- *'I think this would be more important for scaleup - but depends on the intervention duration;*
- *'This is relevant for intervention fidelity, particularly if these sessions were offered/provided during the course of the intervention delivery'*

| | Not important | | | | | Very important | | | |
|---|---|---|---|---|---|---|---|---|---|
| | 1 | 2 | 3 | 4 | 5 | 6 | 7 | 8 | 9 |
| ITEM 33 | | | | | | | | | |

Please provide any comments or suggestions you have about ITEM 33.

**ITEM 34:** Report whether the **planned number of intervention providers** to deliver the intervention within the trial was achieved.

Ratings from Round 1:

Not applicable because this item has changed significantly from the round 1.

Comments on the ratings from panellists in Round 1:

Not applicable because this item has changed significantly from the round 1.

| | Not important | | | | | Very important | | | |
|---|---|---|---|---|---|---|---|---|---|
| | 1 | 2 | 3 | 4 | 5 | 6 | 7 | 8 | 9 |
| ITEM 34 | | | | | | | | | |

Please provide any comments or suggestions you have about ITEM 34.

**ITEM 35**

**ITEM 35:** When applicable, describe **contextual factors** assessed or observed by the research team that may have influenced intervention fidelity assessment or results (e.g., intervention complexity, intervention provider caseload, time spent by providers on small talk vs time delivering the intervention as intended, etc).

Ratings from Round 1:

Not applicable because this item has changed significantly from the round 1.

Comments on the ratings from panellists in Round 1:

Not applicable because this item has changed significantly from the round 1.

| | Not important | | | | | Very important | | | |
|---|---|---|---|---|---|---|---|---|---|
| | 1 | 2 | 3 | 4 | 5 | 6 | 7 | 8 | 9 |
| ITEM 35 | | | | | | | | | |

Please provide any comments or suggestions you have about ITEM 35.

#### ITEM 36-42

Please rate the importance of including the items below in a reporting guideline for intervention fidelity in non-drug, non-surgical trials.

Note: Some items are closely worded but their main substance is unique (highlighted in **bold**). Comments from Round 1 are reported verbatim, with some repeated across items as provided by the panellists.

**ITEM 36:** Describe any revisions to intervention fidelity assessments based on findings from the **intervention development process (e.g., piloting the intervention)**.

Ratings from Round 1:

Not applicable because this item has changed significantly from the round 1.

Comments on the ratings from panellists in Round 1:

Not applicable because this item has changed significantly from the round 1.

| | Not important | | | | | Very important | | |
|---|---|---|---|---|---|---|---|---|
| | 1 | 2 | 3 | 4 | 5 | 6 | 7 | 8 | 9 |
| ITEM 36 | | | | | | | | | |

Please provide any comments or suggestions you have about ITEM 36.

**ITEM 37:** Specify whether the intervention fidelity **measures/tools were piloted**.

Ratings from Round 1:

| **Median (IQR)** | % of panellists rating this item as **1-3** | % of panellists rating this item as **4-6** | % of panellists rating this item as **7-9** |
|---|---|---|---|
| 6 (4, 7) | 18.0 | 47.5 | 34.4 |

Comments on the ratings from panellists in Round 1:

- *'This refers to development of an outcome measure and should not be included in the reporting guideline. That belongs to a satellite manuscript'*
- *'If the fidelity measure is straightforward or has been validated in previous studies, not important'*
- *'Important but not critical for understanding whether the intervention was delivered'*

| | Not important | | | | | Very important | | | |
|---|---|---|---|---|---|---|---|---|---|
| | 1 | 2 | 3 | 4 | 5 | 6 | 7 | 8 | 9 |
| ITEM 37 | | | | | | | | | |

Please provide any comments or suggestions you have about ITEM 37.

**ITEM 38:** Describe how intervention fidelity **measures/tools were developed**. If existing measures/tools are used, specify them and provide their source.

Ratings from Round 1:

Not applicable because this item has changed significantly from the round 1.

Comments on the ratings from panellists in Round 1:

Not applicable because this item has changed significantly from the round 1.

| | Not important | | | | | Very important | | | |
|---|---|---|---|---|---|---|---|---|---|
| | 1 | 2 | 3 | 4 | 5 | 6 | 7 | 8 | 9 |
| ITEM 38 | | | | | | | | | |

Please provide any comments or suggestions you have about ITEM 38.

**ITEM 39:** Specify how the intervention fidelity **measures/tools align with the intervention content and activities**.

Ratings from Round 1:

| **Median (IQR)** | % of panellists rating this item as **1-3** | % of panellists rating this item as **4-6** | % of panellists rating this item as **7-9** |
|---|---|---|---|
| 7 (5, 8) | 14.8 | 29.5 | 55.7 |

Comments on the ratings from panellists in Round 1:

- *'Quite often the connections are self evident and do not require a detailed justification. I would view this as being something authors can add if they want rather than make it a core requirement'*
- *'Fidelity should align with the intervention content and activities engaged in during the intervention'*
- *'Align with intervention content, yes'*

| | Not important | | | | | Very important | | | |
|---|---|---|---|---|---|---|---|---|---|
| | 1 | 2 | 3 | 4 | 5 | 6 | 7 | 8 | 9 |
| ITEM 39 | | | | | | | | | |

Please provide any comments or suggestions you have about ITEM 39.

**ITEM 40: Describe each item** in the intervention fidelity measure/tool and the **process of their selection or development.**

Ratings from Round 1:

| **Median (IQR)** | % of panellists rating this item as **1-3** | % of panellists rating this item as **4-6** | % of panellists rating this item as **7-9** |
|---|---|---|---|
| 6 (3, 7) | 31.1 | 39.3 | 29.5 |

Comments on the ratings from panellists in Round 1:

- *'Should be covered when discussing the theoretical underpinning of the tools used. Only if these are made from scratch, this can be of importance'*
- *'Belongs to a satellite publication'*
- *'Not needed for each individual item'*
- *'For replicability'*
- *'I think it's important to give a sense of how fidelity measures were developed. I think the amount of information provided on this will depend on the overall focus of the paper e.g. methods vs findings paper'*

| | Not important | | | | | Very important | | | |
|---|---|---|---|---|---|---|---|---|---|
| | 1 | 2 | 3 | 4 | 5 | 6 | 7 | 8 | 9 |
| ITEM 40 | | | | | | | | | |

Please provide any comments or suggestions you have about ITEM 40.

**ITEM 41:** Specify the **perceived usability of the fidelity measures/tools** (e.g., how straightforward they are to use or implement).

Ratings from Round 1:

| **Median (IQR)** | % of panellists rating this item as **1-3** | % of panellists rating this item as **4-6** | % of panellists rating this item as **7-9** |
|---|---|---|---|
| 5 (3, 7) | 31.1 | 32.8 | 36.1 |

Comments on the ratings from panellists in Round 1:

- *'These issues are useful to know but not essential for reporting'*
- *'Not relevant'*
- *'This is adjunct work and a nice to have – not essential in the trial report to me'*
- *'Somewhat subjective'*

| | Not important | | | | | | | | Very important |
|---|---|---|---|---|---|---|---|---|---|
| | 1 | 2 | 3 | 4 | 5 | 6 | 7 | 8 | 9 |
| ITEM 41 | | | | | | | | | |

Please provide any comments or suggestions you have about ITEM 41.

**ITEM 42:** Describe the **strengths and limitations** of the intervention fidelity assessment methods used.

Ratings from Round 1:

| **Median (IQR)** | % of panellists rating this item as **1-3** | % of panellists rating this item as **4-6** | % of panellists rating this item as **7-9** |
|---|---|---|---|
| 6 (4, 8) | 19.7 | 32.8 | 47.5 |

Comments on the ratings from panellists in Round 1:

- *'Nice to have – not essential'*
- *'This should be apparent to informed reader'*
- *'This is part of the discussion of the trial manuscript, not the reporting guideline'*
- *'I think it is important to have this reflection'*
- *'It is crucial to thoroughly reflect on both strengths and limitations'*

| | Not important | | | | | Very important | | | |
|---|---|---|---|---|---|---|---|---|---|
| | 1 | 2 | 3 | 4 | 5 | 6 | 7 | 8 | 9 |
| ITEM 42 | | | | | | | | | |

Please provide any comments or suggestions you have about ITEM 42.

**ITEM 43-46**

Please rate the importance of including the items below in a reporting guideline for intervention fidelity in non-drug, non-surgical trials.

Note: Some items are closely worded but their main substance is unique (highlighted in **bold**). Comments from Round 1 are reported verbatim, with some repeated across items as provided by the panellists.

**ITEM 43:** If **different weights were assigned to items** in an intervention fidelity measure/tool that has not been externally validated, report how these weights were determined (e.g., theory, empirical evidence, sensitivity analysis) and the rationale for this approach.

Ratings from Round 1:

| **Median (IQR)** | % of panellists rating this item as **1-3** | % of panellists rating this item as **4-6** | % of panellists rating this item as **7-9** |
|---|---|---|---|
| 6 (5, 8) | 13.1 | 42.6 | 44.3 |

Comments on the ratings from panellists in Round 1:

- *'It may be useful, but I do not think such information needs to be included in the core item set'*
- *'This belongs to psychometric studies assessing and exploring methods to measure treatment fidelity'*
- *'May help identify efficacious components and which fidelity components are essential'*

| | Not important | | | | | Very important | | | |
|---|---|---|---|---|---|---|---|---|---|
| | 1 | 2 | 3 | 4 | 5 | 6 | 7 | 8 | 9 |
| ITEM 43 | | | | | | | | | |

Please provide any comments or suggestions you have about ITEM 43.

**ITEM 44: Specify the psychometric properties** of the intervention fidelity measures/tools (e.g., reliability, validity, and sensitivity to change).

Ratings from Round 1:

| **Median (IQR)** | % of panellists rating this item as **1-3** | % of panellists rating this item as **4-6** | % of panellists rating this item as **7-9** |
|---|---|---|---|
| 7 (5, 8) | 11.5 | 36.1 | 52.5 |

Comments on the ratings from panellists in Round 1:

- *'I honestly don't see merit in this as this is resource intensive and unlikely to be helpful across different trials'*
- *'Ideally – but not always'*
- *'Many of these measures don't have psychometric properties, they are quite simply descriptive statistics'*

- *‘Often fidelity assessments rely on face validity and are not a standardized instrument (e.g., number of sessions attended)’*
- *‘For scientific rigor – good practice’*

| | Not important | | | | | Very important | | | |
|---|---|---|---|---|---|---|---|---|---|
| | 1 | 2 | 3 | 4 | 5 | 6 | 7 | 8 | 9 |
| ITEM 44 | | | | | | | | | |

Please provide any comments or suggestions you have about ITEM 44.

**ITEM 45:** Specify the benchmarks used to **interpret the psychometrics** of the intervention fidelity measures/tools. For example, report what constitutes an acceptable level of inter-rater reliability.

Ratings from Round 1:

| **Median (IQR)** | % of panellists rating this item as **1-3** | % of panellists rating this item as **4-6** | % of panellists rating this item as **7-9** |
|---|---|---|---|
| 6 (3, 7) | 29.5 | 24.6 | 45.9 |

Comments on the ratings from panellists in Round 1:

- *‘Again relevant, but this is to me adjunct work that could be reported alongside a trial - not directly in the trial report’*
- *‘It is important that fidelity measures are comparable (currently comparing across studies is v problematic for this reason) - but info on e.g. inter-rater reliability is fine, even if benchmarks not indicated’*
- *‘This should not be part of treatment fidelity reporting guideline. That refers to the development of the tool or measurement used in a trial to assess treatment fidelity. In this case, this item should not be reported on a satellite study’*

- *‘These thresholds are often arbitrary and relatively meaningless. Eg. if a threshold is set at 80%, dichotomising everything above 80% as good and everything below as bad is not as helpful as just stating what the actual percentage was or distribution of percentages were’*
- *‘This component is critical. Without confidence in the methods used to assess fidelity, we cannot determine whether the intervention was delivered as intended’*

| | Not important | | | | | | Very important | | |
|---|---|---|---|---|---|---|---|---|---|
| | 1 | 2 | 3 | 4 | 5 | 6 | 7 | 8 | 9 |
| ITEM 45 | | | | | | | | | |

Please provide any comments or suggestions you have about ITEM 45.

**ITEM 46:** Specify an a priori threshold for **acceptable level of intervention fidelity** or cut-offs.

Ratings from Round 1:

| **Median (IQR)** | % of panellists rating this item as **1-3** | % of panellists rating this item as **4-6** | % of panellists rating this item as **7-9** |
|---|---|---|---|
| 7 (5, 8) | 14.8 | 19.7 | 65.6 |

Comments on the ratings from panellists in Round 1:

- *‘I think it is more important to understand the context/influencing factors on trial outcomes than to have a dichotomous results for acceptable level of fidelity’*
- *‘As with many research endeavours, I think we should encourage colleagues to interpret findings based on their complexity and without adopting a dichotomous perspective. Interventions are complex and so should be the interpretation of the findings’*
- *‘Thresholds are often arbitrary and meaningless’*

- *'I think this is very important but equally I recognise there is a challenge in that there are not many reported threshold criteria in the literature, and lack of consensus. So it is a bit of a chicken and egg thing! Hard to report due to lack of consensus thresholds, but hard to develop these without evidence and interpretation around thresholds being reported in the literature'*
- *'Of course- but no-one really knows what that is yet...'*
- *'Important, so 'core"*
- *'It is a must to have acceptable cutoff value for an intervention fidelity. This is one of the main limitations while measuring intervention fidelity, to declare high. Medium or low'*

| | Not important | | | | | Very important | | | |
|---|---|---|---|---|---|---|---|---|---|
| | 1 | 2 | 3 | 4 | 5 | 6 | 7 | 8 | 9 |
| ITEM 46 | | | | | | | | | |

Please provide any comments or suggestions you have about ITEM 46.

**ITEM 47-50**

Please rate the importance of including the items below in a reporting guideline for intervention fidelity in non-drug, non-surgical trials.

Note: Some items are closely worded but their main substance is unique (highlighted in **bold**). Comments from Round 1 are reported verbatim, with some repeated across items as provided by the panellists.

**ITEM 47:** Report intervention fidelity **results for all study arms**, including the control intervention arm.

Ratings from Round 1:

| **Median (IQR)** | % of panellists rating this item as **1-3** | % of panellists rating this item as **4-6** | % of panellists rating this item as **7-9** |
|---|---|---|---|

| | | | |
|---|---|---|---|
| 8 (7, 9) | 9.8 | 9.8 | **80.3** |

Comments on the ratings from panellists in Round 1:

- *'Very useful for head to head comparisons, possibly also for social control conditions, but perhaps less so for comparisons with a "usual care" comparison'*
- *'Important for trial design but not intervention-specific fidelity'*
- *'This is very important. Too often it is assumed that controls receive no intervention and of course this may not be the case'*
- *'Sometimes there is no assessment of fidelity in a control intervention, but apart from that example I think fidelity needs to be assessed and reported in all 'relevant' arms'*
- *'I like that the control group is focused upon!'*
- *'This is essential'*

| | Not important | | | | | Very important | | | |
|---|---|---|---|---|---|---|---|---|---|
| | 1 | 2 | 3 | 4 | 5 | 6 | 7 | 8 | 9 |
| ITEM 47 | | | | | | | | | |

Please provide any comments or suggestions you have about ITEM 47.

**ITEM 48:** Report intervention fidelity **in terms of overall fidelity of delivery** (i.e., fidelity to intervention core components across sessions combined). *Session is defined as a distinct unit of time during which the intervention is delivered. Synonyms include 'visit', 'consultation', 'meeting', 'event', etc.

Ratings from Round 1:

| **Median (IQR)** | % of panellists rating this item as **1-3** | % of panellists rating this item as **4-6** | % of panellists rating this item as **7-9** |
|---|---|---|---|
| 8 (7, 9) | 1.6 | 13.1 | **85.2** |

Comments on the ratings from panellists in Round 1:

- *'I don't believe its appropriate to recommend that this approach to assessing fidelity is the ONE to recommend. It also data heavy which feels unrealistic. It might be possible in some RCTs'*
- *'The overall fidelity may not be informative for intervention designers/implementers'*
- *'I rated this as important as an overall fidelity rating is often all we can obtain and we often only have 'room' for one fidelity variable in analysis of fidelity and trial outcomes'*
- *'This is particularly important if there are no obvious 'essential' components (although the identification of so-called 'essential' components might also be open to criticism, e.g. how identified)'*

| | Not important | | | | | | Very important | | |
|---|---|---|---|---|---|---|---|---|---|
| | 1 | 2 | 3 | 4 | 5 | 6 | 7 | 8 | 9 |
| ITEM 48 | | | | | | | | | |

Please provide any comments or suggestions you have about ITEM 48.

**ITEM 49:** Report intervention fidelity **in terms of component fidelity** (i.e., fidelity to each intervention core component across sessions).

Ratings from Round 1:

| **Median (IQR)** | % of panellists rating this item as **1-3** | % of panellists rating this item as **4-6** | % of panellists rating this item as **7-9** |
|---|---|---|---|
| 8 (7, 9) | 1.6 | 19.7 | **78.7** |

Comments on the ratings from panellists in Round 1:

- *'I don't believe its appropriate to recommend that this approach to assessing fidelity is the ONE to recommend. It also data heavy which feels unrealistic. It might be possible in some RCTs. But no RCT will subgroup the analysis into the patients for whom each and every component of treatment was delivered to fidelity - feels unrealistic'*
- *'Very importance but one of the barriers to reporting overall fidelity of delivery is that it can be a comprehensive and resource-intensive process. Qualitative studies, such as ethnographic field studies, is often require: -)'*
- *'if possible this can be useful for understanding details of the mechanisms of impact of an intervention ... but it is not always feasible'*
- *'This is important to help build upon interventions/training so should be widely encouraged'*
- *'Detailed reporting on component fidelity helps guide future implementations by highlighting which components are essential to replicate precisely'*

- *'Important as will identify any core components that are either not being delivered/delivered well, and then identify whether these need to be better specified in the protocol, or providers need more in-depth training in specific components'*

| | Not important | | | | | Very important | | | |
|---|---|---|---|---|---|---|---|---|---|
| | 1 | 2 | 3 | 4 | 5 | 6 | 7 | 8 | 9 |
| ITEM 49 | | | | | | | | | |

Please provide any comments or suggestions you have about ITEM 49.

**ITEM 50:** Report intervention fidelity **in terms of session fidelity** (i.e., fidelity to intervention core components within sessions). *Session is defined as a distinct unit of time during which the intervention is delivered. Synonyms include 'visit', 'consultation', 'meeting', 'event', etc.

Ratings from Round 1:

| **Median (IQR)** | % of panellists | % of panellists | % of panellists |
|---|---|---|---|

| | | | |
|---|---|---|---|
| | rating this item as **1-3** | rating this item as **4-6** | rating this item as **7-9** |
| 7 (5, 8) | 9.8 | 36.1 | 54.1 |

Comments on the ratings from panellists in Round 1:

- *'Again depends on nature of intervention, not all interventions will have multiple sessions- may also be other dimensions that might come up for later items, but things like fidelity over time, over sites, over providers etc'*
- *'Only possible if resources are available, hence maybe less important than reporting fidelity across sessions (if any)'*
- *'Like anything (most of the time anyway), the more data the better, for understanding;*
- *'Yes dose-response to avoid type III errors (Basch)'*

| | Not important | | | | | | | Very important | |
|---|---|---|---|---|---|---|---|---|---|
| | 1 | 2 | 3 | 4 | 5 | 6 | 7 | 8 | 9 |
| ITEM 50 | | | | | | | | | |

Please provide any comments or suggestions you have about ITEM 50.

**ITEM 51**

**ITEM 51:** Describe a **specific intervention fidelity assessment and monitoring protocol** prior to trial initiation/trial analysis.

Ratings from Round 1:

| **Median (IQR)** | % of panellists rating this item as **1-3** | % of panellists rating this item as **4-6** | % of panellists rating this item as **7-9** |
|---|---|---|---|
| 7 (6, 9) | 4.9 | 37.7 | 57.4 |

Comments on the ratings from panellists in Round 1:

- *‘This would be ideal, but it is not always possible or quite difficult in the actual research/implementation environment’*
- *‘Important to do for effectiveness trials, but feels overkill for pilot trials’*
- *‘This item is important but not essential because we don't know will happen during the implementation’*
- *‘Important but bearing in mind the need to find a balance between fidelity and contextual adaptation, which can happen in real time due to shifts in the internal / external environment and difficult to plan per protocol (e.g., impact of Covid) - hence leaving a margin for adaptation is useful’*
- *‘...recognizing that the fidelity play may need to be modified/adjusted once the trial begins (but this should be documented)’*
- *‘I believe this is critical. I also think the results of the fidelity assessment should be analysed and released before the main trial analysis or conducted by blinded, independent assessors’*

| | Not important | | | | | Very important | | | |
|---|---|---|---|---|---|---|---|---|---|
| | 1 | 2 | 3 | 4 | 5 | 6 | 7 | 8 | 9 |
| ITEM 51 | | | | | | | | | |

Please provide any comments or suggestions you have about ITEM 51.

**ITEM 52-53**

Please rate the importance of including the items below in a reporting guideline for intervention fidelity in non-drug, non-surgical trials.

Note: Some items are closely worded but their main substance is unique (highlighted in **bold**). Comments from Round 1 are reported verbatim, with some repeated across items as provided by the panellists.

**ITEM 52:** Specify when **intervention fidelity data was analysed**, such as concurrently with the intervention or after the intervention period was completed.

Ratings from Round 1:

Not applicable because this item has changed significantly from the round 1.

Comments on the ratings from panellists in Round 1:

Not applicable because this item has changed significantly from the round 1.

| | Not important | | | | | Very important | | | |
|---|---|---|---|---|---|---|---|---|---|
| | 1 | 2 | 3 | 4 | 5 | 6 | 7 | 8 | 9 |
| ITEM 52 | | | | | | | | | |

Please provide any comments or suggestions you have about ITEM 52.

**ITEM 53:** Specify the timepoints of the intervention where **intervention fidelity data was collected** (e.g., week 1, 6, and 12).

Ratings from Round 1:

Not applicable because this item has changed significantly from the round 1.

Comments on the ratings from panellists in Round 1:

Not applicable because this item has changed significantly from the round 1.

| | Not important | | | | | Very important | | | |
|---|---|---|---|---|---|---|---|---|---|
| | 1 | 2 | 3 | 4 | 5 | 6 | 7 | 8 | 9 |
| ITEM 53 | | | | | | | | | |

Please provide any comments or suggestions you have about ITEM 53.

##### ITEM 54-55

Please rate the importance of including the items below in a reporting guideline for intervention fidelity in non-drug, non-surgical trials.

Note: Some items are closely worded but their main substance is unique (highlighted in **bold**). Comments from Round 1 are reported verbatim, with some repeated across items as provided by the panellists.

**ITEM 54:** Describe how intervention fidelity **assessors were trained** to use the intervention fidelity measures/tools and conduct the fidelity assessments.

Ratings from Round 1:

| **Median (IQR)** | % of panellists rating this item as **1-3** | % of panellists rating this item as **4-6** | % of panellists rating this item as **7-9** |
|---|---|---|---|
| 7 (5, 8) | 8.2 | 37.7 | 54.1 |

Comments on the ratings from panellists in Round 1:

- *'This is important but perhaps not as important as information on the competency of the fidelity assessor at end of training'*
- *'This makes me think bc it does not seem hard, but only bc I have done it. So not sure'*
- *'This component is critical. Without confidence in the methods used to assess fidelity, we cannot determine whether the intervention was delivered as intended'*

| | Not important | | | | | Very important | | | |
|---|---|---|---|---|---|---|---|---|---|
| | 1 | 2 | 3 | 4 | 5 | 6 | 7 | 8 | 9 |
| ITEM 54 | | | | | | | | | |

Please provide any comments or suggestions you have about ITEM 54.

**ITEM 55:** Specify if there was any requirement for the **selection of intervention fidelity assessors** (e.g., be a health registered professional).

Ratings from Round 1:

| **Median (IQR)** | % of panellists rating this item as **1-3** | % of panellists rating this item as **4-6** | % of panellists rating this item as **7-9** |
|---|---|---|---|
| 6 (5, 7) | 11.5 | 42.6 | 45.9 |

Comments on the ratings from panellists in Round 1:

- *'Not essential'*
- *'Useful to know – but actually if they can assess competently more important than who they are'*
- *'Specifying requirements for the selection of fidelity assessors is important for ensuring the expertise, credibility, and consistency of the fidelity evaluations. It ultimately contributes to the integrity and reliability of the research findings'*
- *'Important, but I believe outlining the procedure for training assessors is even more crucial'*

| | Not important | | | | | | Very important | | |
|---|---|---|---|---|---|---|---|---|---|
| | 1 | 2 | 3 | 4 | 5 | 6 | 7 | 8 | 9 |
| ITEM 55 | | | | | | | | | |

Please provide any comments or suggestions you have about ITEM 55.

**ITEM 56**

**ITEM 56:** Report a **clear intervention manual or detailed intervention protocol**.

Ratings from Round 1:

| **Median (IQR)** | % of panellists rating this item as **1-3** | % of panellists rating this item as **4-6** | % of panellists rating this item as **7-9** |
|---|---|---|---|
| 9 (7, 9) | 6.6 | 9.8 | **83.6** |

Comments on the ratings from panellists in Round 1:

- *'While these are highly important for reporting guidelines for intervention, not highly important for reporting guidelines for intervention fidelity'*
- *'This also falls under general reproducibility, not just fidelity'*
- *'This is key to understand the quality and relevance of fidelity methods'*
- *'This is really important, as without this it's not clear what exactly providers should be trained in'*
- *'Important and feels 'core''*

| | Not important | | | | | Very important | | | |
|---|---|---|---|---|---|---|---|---|---|
| | 1 | 2 | 3 | 4 | 5 | 6 | 7 | 8 | 9 |
| ITEM 56 | | | | | | | | | |

Please provide any comments or suggestions you have about ITEM 56.

**ITEM 57**

**ITEM 57:** Describe what **recruitment procedures** were used to attract participants to the intervention and any barriers to maintaining their involvement.

Ratings from Round 1:

| **Median (IQR)** | % of panellists rating this item as **1-3** | % of panellists rating this item as **4-6** | % of panellists rating this item as **7-9** |
|---|---|---|---|
| 5 (3, 8) | 41.0 | 19.7 | **39.3** |

Comments on the ratings from panellists in Round 1:

- *'This is trial in general, not intervention fidelity'*
- *'This is extremely important to include, but I view it as being outside of the reporting of fidelity per se'*
- *'This is less relevant to the specific reporting of intervention fidelity'*
- *'Important, but not fidelity?'*
- *'Important for replication studies and generalizebility of the study results'*

| | Not important | | | | | | Very important | | |
|---|---|---|---|---|---|---|---|---|---|
| | 1 | 2 | 3 | 4 | 5 | 6 | 7 | 8 | 9 |
| ITEM 57 | | | | | | | | | |

Please provide any comments or suggestions you have about ITEM 57.

**ITEM 58**

**ITEM 58:** Report whether the **cultural and linguistic aspects of the intervention** were delivered as planned, if applicable.

Ratings from Round 1:

| **Median (IQR)** | % of panellists | % of panellists | % of panellists |
|---|---|---|---|

| | | | |
|---|---|---|---|
| | rating this item as **1-3** | rating this item as **4-6** | rating this item as **7-9** |
| 6 (5, 8) | 19.7 | 31.1 | 49.2 |

Comments on the ratings from panellists in Round 1:

- *'Not related to intervention fidelity'*
- *'This is study design'*
- *'An intervention must be adapted before being applied to a sample with different cultural, social, and economic characteristics. Therefore, it is not a point related to fidelity'*
- *'this is not intervention fidelity but rather Equity/Diversity/Inclusivity considerations - different domain/construct and should be part of the work-up of RCTs but not part of reporting guidance for intervention fidelity'*
- *'High importance as it will impact fidelity ultimately'*
- *'Important for tailorability and buy-in'*

| | Not important | | | | | Very important | | | |
|---|---|---|---|---|---|---|---|---|---|
| | 1 | 2 | 3 | 4 | 5 | 6 | 7 | 8 | 9 |
| ITEM 58 | | | | | | | | | |

Please provide any comments or suggestions you have about ITEM 58.

**ITEM 59**

**ITEM 59:** Specify the **frameworks and guidance** used to inform intervention fidelity assessments.

Ratings from Round 1:

| **Median (IQR)** | % of panellists rating this item as **1-3** | % of panellists rating this item as **4-6** | % of panellists rating this item as **7-9** |
|---|---|---|---|

| | | | |
|---|---|---|---|
| 7 (6, 8) | 6.6 | 36.1 | 57.4 |

Comments on the ratings from panellists in Round 1:

- *'Given the current lack of guidelines to be considered in view of the outcomes of this exercise'*
- *'Important to know what informed intervention fidelity assessments'*
- *'Specifying the frameworks and guidance used to inform fidelity assessments is essential for ensuring transparency, consistency, and rigor in the evaluation process'*

| | Not important | | | | | | | | Very important |
|---|---|---|---|---|---|---|---|---|---|
| | 1 | 2 | 3 | 4 | 5 | 6 | 7 | 8 | 9 |
| ITEM 59 | | | | | | | | | |

Please provide any comments or suggestions you have about ITEM 59.

**ITEM 60**

**ITEM 60:** Specify how **intervention fidelity is defined and conceptualised**.

Ratings from Round 1:

| **Median (IQR)** | % of panellists rating this item as **1-3** | % of panellists rating this item as **4-6** | % of panellists rating this item as **7-9** |
|---|---|---|---|
| 8 (7, 9) | 0.0 | 18.0 | **82.0** |

Comments on the ratings from panellists in Round 1:

- *'This is needed to aid interpretation'*

- *This is critical, particularly as there are mixed views about intervention fidelity'*

| | Not important | | | | | Very important | | | |
|---|---|---|---|---|---|---|---|---|---|
| | 1 | 2 | 3 | 4 | 5 | 6 | 7 | 8 | 9 |
| ITEM 60 | | | | | | | | | |

Please provide any comments or suggestions you have about ITEM 60.

**ITEM 61**

**ITEM 61:** If not all intervention fidelity domains were assessed, describe the rationale for selecting the **assessed domains**.

Ratings from Round 1:

| **Median (IQR)** | % of panellists rating this item as **1-3** | % of panellists rating this item as **4-6** | % of panellists rating this item as **7-9** |
|---|---|---|---|
| 7 (6, 8) | 4.9 | 32.8 | 62.3 |

Comments on the ratings from panellists in Round 1:

- *'If you have space. normally this is barn door obvious and might not specific comment'*
- *'Probably should assess them all'*
- *'Important, but what answer can we expect besides no time, not enough money, burden for the target groups'*
- *'Important to set scene in intro as why not including certain domains'*

| | Not important | | | | | Very important | | | |
|---|---|---|---|---|---|---|---|---|---|
| | 1 | 2 | 3 | 4 | 5 | 6 | 7 | 8 | 9 |
| ITEM 61 | | | | | | | | | |

Please provide any comments or suggestions you have about ITEM 61.

**ITEM 62**

**ITEM 62:** Describe how **stakeholders** were involved in the development of the intervention fidelity measures/tools.

Ratings from Round 1:

| **Median (IQR)** | % of panellists rating this item as **1-3** | % of panellists rating this item as **4-6** | % of panellists rating this item as **7-9** |
|---|---|---|---|
| 5 (4, 7) | 21.3 | 45.9 | 32.8 |

Comments on the ratings from panellists in Round 1:

- *‘Not essential and not relevant most times as the measures are directly built based on the intervention protocol’*
- *‘This belongs to psychometric properties of a measurement or tool. That would be a satellite study’*
- *‘I don't think this is always necessary’*
- *‘Important for method transparency’*
- *‘Stakeholders are essential from the beginning of project to ensure the needs of all parties are met and for sustainability’*

| | Not important | | | | | Very important | | | |
|---|---|---|---|---|---|---|---|---|---|
| | 1 | 2 | 3 | 4 | 5 | 6 | 7 | 8 | 9 |
| ITEM 62 | | | | | | | | | |

Please provide any comments or suggestions you have about ITEM 62.

#### ITEM 63

**ITEM 63:** When applicable, justify the **number of response categories** in the intervention fidelity measure/tool, for example, why a 3-point scale (delivered, partially delivered, not delivered) was chosen over other scales.

Ratings from Round 1:

| **Median (IQR)** | % of panellists rating this item as **1-3** | % of panellists rating this item as **4-6** | % of panellists rating this item as **7-9** |
|---|---|---|---|
| 5 (3, 7) | 26.2 | 45.9 | 27.9 |

Comments on the ratings from panellists in Round 1:

- *‘As long as a tool is used, I don't think it matter how many categories were used (thinking purely from a feasibility aspect)’*
- *‘Useful, but could be done as a parenthetical citation rather than a full description’*
- *‘Dont think this matters”*
- *'Too specific for a high-level reporting guideline'*

| | Not important | | | | | | Very important | | |
|---|---|---|---|---|---|---|---|---|---|
| | 1 | 2 | 3 | 4 | 5 | 6 | 7 | 8 | 9 |
| ITEM 63 | | | | | | | | | |

Please provide any comments or suggestions you have about ITEM 63.

**ITEM 64**

**ITEM 64:** Describe how **data from multiple intervention fidelity measurement methods**, when applicable, are integrated and triangulated to provide comprehensive fidelity results. For example, describe how quantitative data (e.g., from fidelity measures/tools) are combined with qualitative data (e.g., self-reports).

Ratings from Round 1:

| **Median (IQR)** | % of panellists rating this item as **1-3** | % of panellists rating this item as **4-6** | % of panellists rating this item as **7-9** |
|---|---|---|---|
| 7 (6, 9) | 8.2 | 31.1 | 60.7 |

Comments on the ratings from panellists in Round 1:

- *'This should be self-evident?'*
- *'Important only when there are multiple measures of fidelity, which may not be common'*
- *'If applicable'*

| | Not important | | | | | | Very important | | |
|---|---|---|---|---|---|---|---|---|---|
| | 1 | 2 | 3 | 4 | 5 | 6 | 7 | 8 | 9 |
| ITEM 64 | | | | | | | | | |

Please provide any comments or suggestions you have about ITEM 64.

**ITEM 65**

**ITEM 65:** Specify the **type of statistical analysis conducted** to obtain intervention fidelity results (e.g., descriptive, ANOVA, regression).

Ratings from Round 1:

| **Median (IQR)** | % of panellists rating this item as **1-3** | % of panellists rating this item as **4-6** | % of panellists rating this item as **7-9** |
|---|---|---|---|
| 7 (6, 9) | 3.3 | 29.5 | 67.2 |

Comments on the ratings from panellists in Round 1:

- *'Only important if fidelity results are statistically analyzed, which is not necessarily the case'*
- *'Not all assessments may be quantitative'*
- *'This is important'*
- *'Should be included in the Statistical Analysis Plan, just like all the analyses'*

| | Not important | | | | | Very important | | | |
|---|---|---|---|---|---|---|---|---|---|
| | 1 | 2 | 3 | 4 | 5 | 6 | 7 | 8 | 9 |
| ITEM 65 | | | | | | | | | |

Please provide any comments or suggestions you have about ITEM 65.

**ITEM 66**

**ITEM 66:** If applicable, describe the **rationale for any sensitivity/additional analysis** informed by intervention fidelity data (e.g., intention-to-treat versus removing low fidelity cases, mediation analysis based on fidelity to specific intervention components).

Ratings from Round 1:

Not applicable because this item has changed significantly from the round 1.

Comments on the ratings from panellists in Round 1:

Not applicable because this item has changed significantly from the round 1.

| | Not important | | | | | Very important | | | |
|---|---|---|---|---|---|---|---|---|---|
| | 1 | 2 | 3 | 4 | 5 | 6 | 7 | 8 | 9 |
| ITEM 66 | | | | | | | | | |

Please provide any comments or suggestions you have about ITEM 66.

**ITEM 67**

**ITEM 67:** If applicable, specify the rate of **missing intervention fidelity data** and how it was managed.

Ratings from Round 1:

| **Median (IQR)** | % of panellists rating this item as **1-3** | % of panellists rating this item as **4-6** | % of panellists rating this item as **7-9** |
|---|---|---|---|
| 7 (6, 8) | 6.6 | 27.9 | 65.6 |

Comments on the ratings from panellists in Round 1:

- *'Where space allows, if in a secondary publication, then yes this is fine. It feels like we are building expectations for fidelity assessment that is equal to the main trial - the main trial is hard enough to do and report'*
- *'Could be covered as part of general reporting'*
- *'Important for transparency of what findings are based on'*

| | Not important | | | | | Very important | | |
|---|---|---|---|---|---|---|---|---|
| | 1 | 2 | 3 | 4 | 5 | 6 | 7 | 8 | 9 |
| ITEM 67 | | | | | | | | | |

Please provide any comments or suggestions you have about ITEM 67.

**ITEM 68-69**

Please rate the importance of including the items below in a reporting guideline for intervention fidelity in non-drug, non-surgical trials.

Note: Some items are closely worded but their main substance is unique (highlighted in **bold**). Comments from Round 1 are reported verbatim, with some repeated across items as provided by the panellists.

**ITEM 68:** Specify what is the **sample size of the intervention fidelity assessments** and how it was determined (e.g., whole trial sample, subset sample).

Ratings from Round 1:

Not applicable because this item has changed significantly from the round 1.

Comments on the ratings from panellists in Round 1:

Not applicable because this item has changed significantly from the round 1.

| | Not important | | | | | Very important | | |
|---|---|---|---|---|---|---|---|---|
| | 1 | 2 | 3 | 4 | 5 | 6 | 7 | 8 | 9 |
| ITEM 68 | | | | | | | | | |

Please provide any comments or suggestions you have about ITEM 68.

**ITEM 69:** Describe any actions taken to minimise **selection bias when sampling a sub-set** of data points for intervention fidelity analysis.

Ratings from Round 1:

Not applicable because this item has changed significantly from the round 1.

Comments on the ratings from panellists in Round 1:

Not applicable because this item has changed significantly from the round 1.

| | Not important | | | | | Very important | | | |
|---|---|---|---|---|---|---|---|---|---|
| | 1 | 2 | 3 | 4 | 5 | 6 | 7 | 8 | 9 |
| ITEM 69 | | | | | | | | | |

Please provide any comments or suggestions you have about ITEM 69.

**ITEM 70-72**

Please rate the importance of including the items below in a reporting guideline for intervention fidelity in non-drug, non-surgical trials.

Note: Some items are closely worded but their main substance is unique (highlighted in **bold**). Comments from Round 1 are reported verbatim, with some repeated across items as provided by the panellists.

**ITEM 70:** Specify the **number** of intervention fidelity assessors and indicate if they performed their fidelity evaluations **independently**.

Ratings from Round 1:

| **Median (IQR)** | % of panellists rating this item as **1-3** | % of panellists rating this item as **4-6** | % of panellists rating this item as **7-9** |
|---|---|---|---|

| | | | |
|---|---|---|---|
| 7 (6, 8) | 6.6 | 31.1 | 62.3 |

Comments on the ratings from panellists in Round 1:

- *'Depend on assessment methods'*
- *'Fidelity is frequently subjective so know how many raters and their independence lends important information to the confidence one can have in ratings'*
- *'important for transparency'*
- *'This component is critical. Without confidence in the methods used to assess fidelity, we cannot determine whether the intervention was delivered as intended'*

| | Not important | | | | | Very important | | | |
|---|---|---|---|---|---|---|---|---|---|
| | 1 | 2 | 3 | 4 | 5 | 6 | 7 | 8 | 9 |
| ITEM 70 | | | | | | | | | |

Please provide any comments or suggestions you have about ITEM 70.

**ITEM 71:** Specify the **level of blinding** of intervention fidelity assessors at the time of intervention fidelity assessments (e.g., blinded to intervention outcomes or intervention provider identity).

Ratings from Round 1:

| **Median (IQR)** | % of panellists rating this item as **1-3** | % of panellists rating this item as **4-6** | % of panellists rating this item as **7-9** |
|---|---|---|---|
| 7 (6, 8) | 9.8 | 26.2 | 63.9 |

Comments on the ratings from panellists in Round 1:

- *‘Most RCTs of complex interventions wont be blinded, so not 'core' for reporting’*
- *‘In an ideal world everyone would be blind to these things but it often is not practical. To some extent this depends on volume of data that is being analysed. I think blinding to intervention outcomes is probably more important than provider identity (although perhaps this depends of relationship between assessor and providers). I belief fidelity assessment should preferably be blind to outcome.’*
- *‘Specifying the level of blinding of fidelity assessors is critical for ensuring the integrity of the evaluation process, but it also presents practical challenges that researchers must navigate to maintain objectivity and reduce bias in their studies’*
- *‘Useful to know whether or not they were blinded, but need not be blinded’*
- *‘This component is critical. Without confidence in the methods used to assess fidelity, we cannot determine whether the intervention was delivered as intended’*

| | Not important | | | | | Very important | | | |
|---|---|---|---|---|---|---|---|---|---|
| | 1 | 2 | 3 | 4 | 5 | 6 | 7 | 8 | 9 |
| ITEM 71 | | | | | | | | | |

Please provide any comments or suggestions you have about ITEM 71.

**ITEM 72:** Specify whether **intervention fidelity assessors were members of the research team, the intervention delivery team, or external to both**.

Ratings from Round 1:

| **Median (IQR)** | % of panellists rating this item as **1-3** | % of panellists rating this item as **4-6** | % of panellists rating this item as **7-9** |
|---|---|---|---|
| 7 (6, 8) | 6.6 | 19.7 | **73.8** |

Comments on the ratings from panellists in Round 1:

- *‘This would be best practice and should be easy to do’*
- *‘This component is critical. Without confidence in the methods used to assess fidelity, we cannot determine whether the intervention was delivered as intended’*
- *‘This is essential. It is critical to know when the person who provided the intervention was the same reporting intervention fidelity in the trial’*

| | Not important | | | | | Very important | | | |
|---|---|---|---|---|---|---|---|---|---|
| | 1 | 2 | 3 | 4 | 5 | 6 | 7 | 8 | 9 |
| ITEM 72 | | | | | | | | | |

Please provide any comments or suggestions you have about ITEM 72.

**SECTION 3 - Add Input**

(Optional) If you have any further feedback, please share it here:

Powered by Qualtrics
