## Appendix 1 for "Establishing consensus on domains and items for Reporting intervention Fidelity in Non-Drug, non-surgical trials: the ReFiND Delphi study"

#### ReFiND Consent

### Welcome to the ReFiND Delphi Study

Thank you for your interest in this study. Please read the information below before giving consent and taking the survey.

**1. Our aim**

This study is a collective effort by international researchers to develop a consensus-based guideline for reporting intervention fidelity in trials of non-drug, non-surgical interventions. It received approval from the Monash University Human Research Ethics Committee (ID 41579/2024).

For this study, intervention fidelity refers to the extent to which an intervention is implemented as planned in the trial protocol.

We recently conducted a scoping review of existing intervention fidelity guidance documents and found high inconsistency of fidelity recommendations across the field. This lack of consensus is a major barrier to the standardised reporting of intervention fidelity in trials.

The aim of this study is to achieve consensus on a core set of intervention fidelity domains and items to be reported in trials of non-drug, non-surgical interventions. Your input is crucial in shaping this important topic, which will improve the quality of reporting in these trials.

**2. Why you were invited**

You were invited to contribute to this study because you have authored at least one peer-reviewed article on intervention fidelity, research reporting methods, or process evaluation of non-drug, non-surgical interventions. We identified your contact information through your published article(s).

Can you please mark all options below that apply to you?

- [ ] I am an author of a peer-reviewed publication related to reporting methods

☐ I am an author of a peer-reviewed publication related to trials of non-drug, non-surgical interventions

☐ I am an author of a peer-reviewed publication related to the process evaluation of non-drug, non-surgical interventions

☐ I am an author of a peer-reviewed publication related to the fidelity of non-drug, non-surgical interventions

☐ Other. Please specify ____________

**3. Your contribution**

This study consists of three rounds of online surveys, each open for about three weeks. You will receive one email reminder each week during each round. The first round will remain open until **7 November 2024**. We will contact you for the second round in late November. After the third round, you may be invited to an online consensus meeting to review the findings and agree on the structure of the guideline report.

In the **first round**, the survey includes questions about your demographics and asks you to rate the importance of a set of fidelity domains and items using a 9-point scale (1 = not important to 9 = very important). Some optional open-ended questions will also be provided.

In the **second round**, you will receive a summary of the ratings and justifications from the first round, and any new items suggested in the open-ended responses. You will be asked to re-evaluate each domain and items using the same 9-point scale.

In the **third round**, you will be asked to rate domains and items that did not achieve consensus for inclusion or exclusion in the previous round.

This first survey takes about 45 to 75 minutes to complete, but it can vary. You will be asked **to participate in all three rounds of surveys**. Our results may be compromised if participants do not respond to the final two rounds. Therefore, the success of this reporting guideline in driving positive changes in fidelity and trial reporting depends on your participation.

**Co-authorship**

All participants completing all consensus processes (three survey rounds and/or consensus meeting) will have the option to **co-author the guideline statement publication** (if opted in), contingent upon meeting ICJME authorship criteria (e.g., reviewing the manuscript). Your name would be included as part of the ReFiND collaborative group authorship.

We greatly value your contribution!

We will ask for your consent to participate in this study on the next page.

#### Consent request

Please read this Explanatory Statement (click here) in full before deciding whether to participate in this research. If you would like further information about any aspect of the project, you are encouraged to contact the researchers via the email addresses provided in the Explanatory Statement.

By selecting 'Yes' to the following question, you will be giving your consent to participate in this study, and proceed to Round 1 of the Delphi survey. Please read the following statements and respond to the question.

- *I have read and understood the Explanatory Statement.*
- *I understand what my participation in this study involves, including the possible benefits and risks.*
- *I understand that the research team will have access to my email address for contact purposes, but it will be stored separately from my survey responses to maintain confidentiality.*
- *I understand that my survey responses will be de-identified through a unique ID and will be extracted and reported without any link to my personally identifiable information.*
- *I understand I can withdraw from this study at any time, but it will not be possible to withdraw data after survey responses are submitted.*
- ***I give my consent to participate in this study.***

Question: Are these statements true?

○ Yes, I give consent to participate and would like to take the survey

○ No, I do not give consent to participate and would like to stop here

*Monash University values the privacy of every individual's personal information and is committed to the protection of that information from unauthorised use and disclosure except where permitted by law. For more information about Data Protection and Privacy at Monash University please see our Data Protection and Privacy Procedure.*

*If you have any questions about how Monash University is collecting and handling your personal information, please contact our Data Protection and Privacy Office at.*

#### ReFiND Demo

##### Delphi Survey

This survey includes three sections. It is important to note the distinction between fidelity domains and individual recommendation items, which are treated separately in Sections 1 and 2.

**SECTION 1 - Rating Fidelity Domains:** In this section, we will present broad domains of fidelity. These domains represent overarching categories or areas related to intervention fidelity, which were identified through a scoping review. We will ask you to rate the importance of each domain based on its relevance to intervention fidelity evaluation and reporting. We will provide definitions, examples, and further instructions to inform your ratings.
**SECTION 2 - Rating Individual Items:** In this section, we will ask you to rate the importance of specific, individial fidelity recommendation items for inclusion in the reporting guideline for intervention fidelity.
**SECTION 3 - Additional Input:** You will have the opportunity to provide any additional input you may have about the domains, the items, the survey, the study, or anything else.

You can pause the survey at any time by closing the browser and then resume later, as long as you use the same browser where you started. However, please note you may not be able to go back to previous pages once you have submitted your responses.

We will begin by collecting demographic details before proceeding to these survey sections.

What is your name? (This information will be used to contact you in the next round. It will be stored separately from your survey responses, which will remain confidential)

Please enter your email address. (This information will be used to contact you in the next round. It will be stored separately from your survey responses, which will remain confidential)

Please confirm your email address. (This information will be used to contact you in the next round. It will be stored separately from your survey responses, which will remain confidential)

Participants completing all consensus processes (three survey rounds and/or consensus meeting) will have the option to co-author the guideline statement publication, contingent upon meeting the ICJME authorship criteria (e.g., reviewing the manuscript). Your name woud be included as part of the ReFiND collaborative group authorship.

Are you interested in co-authorship?

- ◯ Yes
- ◯ No

We would like to acknowledge participants who contributed to this research in our publication. Do you give consent to have your name included in the acknowledgments section of the publication? (This information will be stored separately from your survey responses, which will remain confidential)

- ◯ Yes, I consent to be acknowledged by name in the publication *(Please specify how your name should appear in the acknowledgments)* ____________
- ◯ No, I prefer to remain anonymous and not be acknowledged by name

Would you like to receive a summary of the results of this study once it is completed?

- ◯ Yes
- ◯ No

How old are you?

- ◯ Under 18
- ◯ 18-24 years old
- ◯ 25-34 years old
- ◯ 35-44 years old
- ◯ 45-54 years old
- ◯ 55-64 years old
- ◯ 65+ years old

What is your gender? (We ask this question to assess the gender inclusivity of this study)

- ○ Woman
- ○ Man
- ○ Non-binary / gender diverse
- ○ Prefer not to say
- ○ My gender is not listed. I identify as: ________

In which country were you born?

[dropdown]

In which country do you currently reside?

[dropdown]

What is your highest educational qualification?

- ○ Bachelor's or equivalent
- ○ Master's or equivalent
- ○ Doctorate (DPhil/PhD) or equivalent
- ○ Prefer not to say/Not applicable
- ○ Other, please specify: ________

What is your academic (or research) title/position?

- ○ Research assistant/PhD student
- ○ Postdoctoral Research Fellow
- ○ Lecturer/Assistant Professor
- ○ Senior Lecturer/Senior Research Fellow
- ○ Associate Professor/Reader/Principal Lecturer
- ○ Professor
- ○ Prefer not to say/Not applicable
- ○ Other, please specify: ________

What are your roles? Please mark all options that apply:

- ☐ Trial investigator
- ☐ Trial methodologist
- ☐ Trial assistant
- ☐ Intervention provider within a trial
- ☐ Implementation scientist
- ☐ Evidence synthesis specialist
- ☐ Statistician
- ☐ Epidemiologist
- ☐ Health economist
- ☐ Journal Editor or Associate Editor
- ☐ Journal Editorial Board member
- ☐ Policymaker
- ☐ Research funding agency member
- ☐ Research ethics committee member
- ☐ Clinician/Health or Allied Health Professional/Social worker
- ☐ Other, please specify: __________

What are your areas of research? Please mark all options that apply:

- ☐ Meta-Research/Research methodology (transparency, reproducibility, credibility)
- ☐ Clinical trials
- ☐ Implementation science
- ☐ Health Services
- ☐ Health Policy and Systems
- ☐ Health Education
- ☐ Health Ethics
- ☐ Health Informatics
- ☐ Digital Health
- ☐ Public, Environmental and Occupational Health
- ☐ Behavioural research
- ☐ Psychology

- [ ] Nursing/Midwifery
- [ ] Social Work
- [ ] Clinical Medicine
- [ ] Rehabilitation
- [ ] Sports Sciences
- [ ] Complementary and Integrative Health
- [ ] Other, please specify: ____________

In which stages and processes of intervention research do you have experience? Please mark all options that apply:

- [ ] Intervention design
- [ ] Feasibility and Pilot trials
- [ ] Explanatory or efficacy trials
  Click here for more info
- [ ] Pragmatic or effectiveness trials
  Click here for more info
- [ ] Effectiveness-implementation hybrid trials
  Click here for more info
- [ ] Implementation trials
  Click here for more info
- [ ] Other, please specify: ____________

#### SECTION 1 - Domains

##### SECTION 1 – Rating Fidelity Domains

In this section, we present fidelity domains with descriptions and examples and ask you to rate their importance for intervention fidelity evaluation and/or reporting. You will use a 9-point scale (1 = 'not important' to 9 = 'very important'). You can also provide comments (e.g., suggestions regarding wording, clarity, etc.) in the optional free-text box accompanying each domain.

**Rating 1 to 3:** indicates the domain is not important for intervention fidelity evaluation and/or reporting.
**Rating 4 to 6:** indicates the domain may be important but is not essential for intervention fidelity evaluation and/or reporting.
**Rating 7 to 9:** indicates the domain is very important for intervention fidelity evaluation and/or reporting.

**Key information to consider while rating items (Please READ CAREFULLY)**

- For this study, a **domain** refers to a specific and connected set of aspects related to intervention fidelity that should be assessed and reported in trials of non-drug, non-surgical interventions.
- We aim to achieve consensus on **domains specific to intervention fidelity**. Therefore, when rating a domain, please focus on its importance to intervention fidelity evaluation and reporting, rather than its relevance to general intervention reporting.
- For this study, **intervention fidelity** refers to the extent to which an intervention is implemented as planned in the trial protocol. This is a working definition and will be updated after the Delphi study to ensure it aligns with the domains and items achieving consensus after the third survey round. Acknowledging differing perspectives on what constitutes fidelity, we **seek your expert opinion** on the importance of each domain presented.
- We present all domains on this same page, so you can review them together. If you find a domain to be less important because it overlaps with or could be integrated into another domain, please **use the comment box to provide your rationale and suggestions**.

Please rate the importance of the domains below for intervention fidelity evaluation and/or reporting:

**Domain 1: Trial and Intervention Design**
Description: This domain covers key elements of the trial and the intervention design pertinent for intervention fidelity. It may evaluate how well the conceptual and/or theoretical models underlying the intervention are reflected in the trial's execution. For example, if an intervention is based on a health behaviour change model, it assesses whether the model's components (e.g., motivation enhancement, self-efficacy building) are integrated into trial measures, provider training, etc.

The following questions use a slider. You can select your response by dragging the slider or clicking near your desired number.

If you do not want to use your mouse to navigate the survey, use the Tab key to move forward through questions and the Arrow keys to select options.

| | Not important 1 | 2 | 3 | 4 | 5 | 6 | 7 | Very important 8 | 9 |
|---|---|---|---|---|---|---|---|---|---|
| D1: Trial and Intervention Design | | | | | | | | | |

Please provide any comments or suggestions you have about Domain 1.

**Domain 2: Provider Selection and Training**
Description: This domain focuses on the criteria and methods used for selecting and training providers to deliver the intervention with adequate fidelity. It may involve reporting on the criteria used to select providers (e.g., qualifications, experience) and evaluating whether all providers received the required training as outlined in the protocol. It may also assess whether they met pre-established performance standards (e.g., achieving at least 80% on a post-training test).

| | Not important 1 | 2 | 3 | 4 | 5 | 6 | 7 | Very important 8 | 9 |
|---|---|---|---|---|---|---|---|---|---|
| D2: Provider Selection and Training | | | | | | | | | |

Please provide any comments or suggestions you have about Domain 2.

**Domain 3: Intervention Delivery by Providers**
Description: This domain focuses on how well and competently trial providers delivered the planned intervention. It may evaluate the extent to which providers adhered to the intervention as outlined in the intervention protocol, including their adherence to core components and avoidance of non-protocolised components. It

may also examine the delivery of intervention components in different trial arms to assess whether there is clear differentiation between interventions (i.e., to address concerns about intervention contamination).

| | Not important | | | | | | Very important | |
|---|---|---|---|---|---|---|---|---|---|
| | 1 | 2 | 3 | 4 | 5 | 6 | 7 | 8 | 9 |
| D3: Intervention Delivery by Providers | | | | | | | | | |

Please provide any comments or suggestions you have about Domain 3.

**Domain 4: Intervention Receipt by Participants**
Description: This domain focuses on how trial participants received and understood the intervention. It may evaluate the extent to which participants received the intervention as planned. For example, in a group intervention where health professionals deliver weekly lectures on diabetes management, it assesses whether participants attended lectures (i.e., received the intervention). It may also examine whether participants understood the instructions, concepts, or content of the intervention as outlined by the intervention's principles. This may include checking whether they can accurately repeat or apply what was taught/delivered. Receipt can be measured both thoroughout and at the end of the intervention period.

| | Not important | | | | | | Very important | |
|---|---|---|---|---|---|---|---|---|---|
| | 1 | 2 | 3 | 4 | 5 | 6 | 7 | 8 | 9 |
| D4: Intervention Receipt by Participants | | | | | | | | | |

Please provide any comments or suggestions you have about Domain 4.

**Domain 5: Intervention Enactment by Participants**

Description: This domain focuses on how trial participants enacted the intervention. It may evaluate the extent to which participants integrate what they learned from the intervention into their routine behaviours. For example, it may assess whether participants who received a lecture on preparing low-carbohydrate meals for diabetes management incorporated these preparations into their daily routines. It may also examine whether they followed specific recommendations from the intervention, such as engaging in prescribed exercises or other activities. Enactment can be measured both throughout and at the end of the intervention period.

| | Not important | | | | | | Very important | | |
|---|---|---|---|---|---|---|---|---|---|
| | 1 | 2 | 3 | 4 | 5 | 6 | 7 | 8 | 9 |
| D5: Intervention Enactment by Participants | | | | | | | | | |

Please provide any comments or suggestions you have about Domain 5.

**Domain 6: Moderators of fidelity and outcomes**

Description: This domain focuses on aspects that may moderate intervention fidelity and the relationship between intervention fidelity and trial outcomes. It may evaluate whether contextual elements, such as the provider-participant relationship or organisational factors (e.g., provider caseload), impact intervention fidelity. It may also examine whether intervention fidelity itself influences trial outcomes, and how the outcomes of an intervention may be interpreted in the context of its fidelity results.

| | Not important | | | | | | Very important | | |
|---|---|---|---|---|---|---|---|---|---|
| | 1 | 2 | 3 | 4 | 5 | 6 | 7 | 8 | 9 |
| D6: Moderators of fidelity and outcomes | | | | | | | | | |

Please provide any comments or suggestions you have about Domain 6.

**Domain 7: Fidelity measurement methods**
Description: This domain focuses on the specific methods used to assess and monitor intervention fidelity in a trial. It may involve detailed reporting on the development of each fidelity measure, including evaluation of psychometric properties. It may also cover the thresholds used to define acceptable levels of fidelity. For example, 'delivery of more than XX% of the core components of the intervention as planned is considered acceptable fidelity'. This domain may also include the number of fidelity assessors and the sample size of intervention sessions reviewed for fidelity assessments.

| | Not important | | | | | | Very important | | |
|---|---|---|---|---|---|---|---|---|---|
| | 1 | 2 | 3 | 4 | 5 | 6 | 7 | 8 | 9 |
| D7: Fidelity measurement methods | | | | | | | | | |

Please provide any comments or suggestions you have about Domain 7.

If you any further comments or suggestions about the domains listed in this survey, please enter them here.

**SECTION 2 - Instructions - items**

**SECTION 2 – Rating Individual Fidelity Reporting Items**

In this section, we present intervention fidelity items and ask you to **please rate their importance for inclusion in the reporting guideline for intervention fidelity**. You will use a 9-point scale (1 = 'not important' to 9 = 'very important'). You can also provide comments (e.g., suggestions regarding wording, clarity, etc.) in the optional free-text box accompanying each domain.

**ITEM 1**

Please rate the importance of including this item in a reporting guideline for intervention fidelity in non-drug, non-surgical trials:

**ITEM 1: Provide a clear and thorough intervention manual or detailed intervention protocol.**

| | Not important | | | | | Very important | | | |
|---|---|---|---|---|---|---|---|---|---|
| | 1 | 2 | 3 | 4 | 5 | 6 | 7 | 8 | 9 |
| ITEM 1 | | | | | | | | | |

Please provide any comments or suggestions you have about ITEM 1.

**ITEM 2**

Please rate the importance of including this item in a reporting guideline for intervention fidelity in non-drug, non-surgical trials:

**ITEM 2: Describe the theoretical or logic model (e.g., the key concepts, activities and resources involved in the intervention's implementation) and report theoretical fidelity, that is, whether the intervention theory or logic model is adequately reflected in intervention delivery.**

| | Not important | | | | | Very important | | | |
|---|---|---|---|---|---|---|---|---|---|
| | 1 | 2 | 3 | 4 | 5 | 6 | 7 | 8 | 9 |
| ITEM 2 | | | | | | | | | |

Please provide any comments or suggestions you have about ITEM 2.

**ITEM 3**

Please rate the importance of including this item in a reporting guideline for intervention fidelity in non-drug, non-surgical trials:

**ITEM 3: Describe a method to assess the extent to which providers delivered the intervention as planned in the intervention protocol (i.e., fidelity of intervention delivery), including aspects such as intervention content, dose (e.g., frequency, duration), progression, and other processes.**

| | Not important | | | | | | Very important | | |
|---|---|---|---|---|---|---|---|---|---|
| | 1 | 2 | 3 | 4 | 5 | 6 | 7 | 8 | 9 |
| ITEM 3 | | | | | | | | | |

Please provide any comments or suggestions you have about ITEM 3.

**ITEM 4-8**

Please rate the importance of including these items in a reporting guideline for intervention fidelity in non-drug, non-surgical trials:

**ITEM 4: Describe the intervention content (i.e., the 'what') planned to be delivered in the trial and report whether it was actually delivered as planned.**

| | Not important | | | | | | Very important | | |
|---|---|---|---|---|---|---|---|---|---|
| | 1 | 2 | 3 | 4 | 5 | 6 | 7 | 8 | 9 |
| ITEM 4 | | | | | | | | | |

Please provide any comments or suggestions you have about ITEM 4.

**ITEM 5: Describe, from the trial outset, the core components of the intervention, which are essential and cannot be modified.**

| | Not important | | | | | Very important | | | |
|---|---|---|---|---|---|---|---|---|---|
| | 1 | 2 | 3 | 4 | 5 | 6 | 7 | 8 | 9 |
| ITEM 5 | | | | | | | | | |

Please provide any comments or suggestions you have about ITEM 5.

**ITEM 6: Describe, from the trial outset, the non-core components where delivery is flexible and adaptations are allowed.**

| | Not important | | | | | Very important | | | |
|---|---|---|---|---|---|---|---|---|---|
| | 1 | 2 | 3 | 4 | 5 | 6 | 7 | 8 | 9 |
| ITEM 6 | | | | | | | | | |

Please provide any comments or suggestions you have about ITEM 6.

**ITEM 7: Describe a method to assess whether prohibited components are delivered (e.g., components that are unnecessary or unhelpful).**

| | Not important | | | | | Very important | | | |
|---|---|---|---|---|---|---|---|---|---|
| | 1 | 2 | 3 | 4 | 5 | 6 | 7 | 8 | 9 |
| ITEM 7 | | | | | | | | | |

Please provide any comments or suggestions you have about ITEM 7.

**ITEM 8: Report whether prohibited components (e.g., components that are unnecessary or unhelpful) were delivered.**

| | Not important | | | | | Very important | | | |
|---|---|---|---|---|---|---|---|---|---|
| | 1 | 2 | 3 | 4 | 5 | 6 | 7 | 8 | 9 |
| ITEM 8 | | | | | | | | | |

Please provide any comments or suggestions you have about ITEM 8.

**ITEM 9**

Please rate the importance of including this item in a reporting guideline for intervention fidelity in non-drug, non-surgical trials:

**ITEM 9: Specify the planned intervention dose for each intervention arm, including the comparison/control intervention arm, and report whether it was actually delivered as planned.**

| | Not important | | | | | Very important | | | |
|---|---|---|---|---|---|---|---|---|---|
| | 1 | 2 | 3 | 4 | 5 | 6 | 7 | 8 | 9 |
| ITEM 9 | | | | | | | | | |

Please provide any comments or suggestions you have about ITEM 9.

**ITEM 10**

Please rate the importance of including this item in a reporting guideline for intervention fidelity in non-drug, non-surgical trials:

**ITEM 10: Describe if the level of interactions that providers had with participants was consistent across trial arms and among all participants within each arm.**

| | Not important | | | | | Very important | | | |
|---|---|---|---|---|---|---|---|---|---|
| | 1 | 2 | 3 | 4 | 5 | 6 | 7 | 8 | 9 |
| ITEM 10 | | | | | | | | | |

Please provide any comments or suggestions you have about ITEM 10.

**ITEM 11**

Please rate the importance of including this item in a reporting guideline for intervention fidelity in non-drug, non-surgical trials:

**ITEM 11: Specify how contamination between trial arms will be prevented, particularly if interventions and comparators are delivered by the same provider.**

| | Not important | | | | | Very important | | | |
|---|---|---|---|---|---|---|---|---|---|
| | 1 | 2 | 3 | 4 | 5 | 6 | 7 | 8 | 9 |
| ITEM 11 | | | | | | | | | |

Please provide any comments or suggestions you have about ITEM 11.

**ITEM 12**

Please rate the importance of including this item in a reporting guideline for intervention fidelity in non-drug, non-surgical trials:

**ITEM 12: Describe what recruitment procedures were used to attract participants to the intervention and what were the barriers to maintaining their involvement.**

| | Not important | | | | | Very important | | | |
|---|---|---|---|---|---|---|---|---|---|
| | 1 | 2 | 3 | 4 | 5 | 6 | 7 | 8 | 9 |
| ITEM 12 | | | | | | | | | |

Please provide any comments or suggestions you have about ITEM 12.

**ITEM 13**

Please rate the importance of including this item in a reporting guideline for intervention fidelity in non-drug, non-surgical trials:

**ITEM 13: Specify the intervention processes (i.e., the how) planned to be implemented in the trial and report whether they were actually implemented as planned. This includes the intervention mode of delivery (e.g., in person, phone), setting (e.g., school, home), format (e.g., group, individual), target participants (e.g., patients, parents).**

| | Not important | | | | | Very important | | | |
|---|---|---|---|---|---|---|---|---|---|
| | 1 | 2 | 3 | 4 | 5 | 6 | 7 | 8 | 9 |
| ITEM 13 | | | | | | | | | |

Please provide any comments or suggestions you have about ITEM 13.

**ITEM 14-15**

Please rate the importance of including these items in a reporting guideline for intervention fidelity in non-drug, non-surgical trials:

**ITEM 14: Describe a method to assess providers ability and skillfulness in delivering the intervention (i.e., 'provider competence').**

| | Not important | | | | | Very important | | | |
|---|---|---|---|---|---|---|---|---|---|
| | 1 | 2 | 3 | 4 | 5 | 6 | 7 | 8 | 9 |
| ITEM 14 | | | | | | | | | |

Please provide any comments or suggestions you have about ITEM 14.

**ITEM 15: Report providers ability and skillfulness in delivering the intervention (i.e., 'provider competence').**

| | Not important | | | | | Very important | | | |
|---|---|---|---|---|---|---|---|---|---|
| | 1 | 2 | 3 | 4 | 5 | 6 | 7 | 8 | 9 |
| ITEM 15 | | | | | | | | | |

Please provide any comments or suggestions you have about ITEM 15.

**ITEM 16**

Please rate the importance of including this item in a reporting guideline for intervention fidelity in non-drug, non-surgical trials:

**ITEM 16: Describe how multicultural factors are considered in the development and delivery of the intervention (e.g., provided in native language; protocol is consistent with the values of the target group).**

| | Not important | | | | | Very important | | | |
|---|---|---|---|---|---|---|---|---|---|
| | 1 | 2 | 3 | 4 | 5 | 6 | 7 | 8 | 9 |
| ITEM 16 | | | | | | | | | |

Please provide any comments or suggestions you have about ITEM 16.

**ITEM 17**

Please rate the importance of including this item in a reporting guideline for intervention fidelity in non-drug, non-surgical trials:

**ITEM 17: Specify how the intervention was piloted, and how the findings from the piloting process informed the fidelity assessments.**

| | Not important | | | | | Very important | | | |
|---|---|---|---|---|---|---|---|---|---|
| | 1 | 2 | 3 | 4 | 5 | 6 | 7 | 8 | 9 |
| ITEM 17 | | | | | | | | | |

Please provide any comments or suggestions you have about ITEM 17.

**ITEM 18**

Please rate the importance of including this item in a reporting guideline for intervention fidelity in non-drug, non-surgical trials:

**ITEM 18: Specify the frameworks and guidance used to inform fidelity assessments, including the development of fidelity measures.**

| | Not important | | | | | Very important | | | |
|---|---|---|---|---|---|---|---|---|---|
| | 1 | 2 | 3 | 4 | 5 | 6 | 7 | 8 | 9 |
| ITEM 18 | | | | | | | | | |

Please provide any comments or suggestions you have about ITEM 18.

**ITEM 19**

Please rate the importance of including this item in a reporting guideline for intervention fidelity in non-drug, non-surgical trials:

**ITEM 19: Describe how intervention complexity was considered when planning and conducting fidelity assessments.**

| | Not important | | | | | Very important | | | |
|---|---|---|---|---|---|---|---|---|---|
| | 1 | 2 | 3 | 4 | 5 | 6 | 7 | 8 | 9 |
| ITEM 19 | | | | | | | | | |

Please provide any comments or suggestions you have about ITEM 19.

**ITEM 20**

Please rate the importance of including this item in a reporting guideline for intervention fidelity in non-drug, non-surgical trials:

**ITEM 20: Specify how fidelity is defined and conceptualised from the trial outset.**

| | Not important | | | | | Very important | | | |
|---|---|---|---|---|---|---|---|---|---|
| | 1 | 2 | 3 | 4 | 5 | 6 | 7 | 8 | 9 |
| ITEM 20 | | | | | | | | | |

Please provide any comments or suggestions you have about ITEM 20.

**ITEM 21**

Please rate the importance of including this item in a reporting guideline for intervention fidelity in non-drug, non-surgical trials:

**ITEM 21: Describe a specific fidelity assessment and monitoring protocol prior to trial initiation/trial analysis. It should provide a clear timeline with an overview of what actions need to be undertaken and when, and a plan for dealing with potential fidelity deviations.**

| | Not important | | | | | Very important | | | |
|---|---|---|---|---|---|---|---|---|---|
| | 1 | 2 | 3 | 4 | 5 | 6 | 7 | 8 | 9 |
| ITEM 21 | | | | | | | | | |

Please provide any comments or suggestions you have about ITEM 21.

**ITEM 22**

Please rate the importance of including this item in a reporting guideline for intervention fidelity in non-drug, non-surgical trials:

**ITEM 22: If all fidelity domains were not assessed, describe the rationale for selecting the assessed domains.**

| | Not important | | | | | Very important | | | |
|---|---|---|---|---|---|---|---|---|---|
| | 1 | 2 | 3 | 4 | 5 | 6 | 7 | 8 | 9 |
| ITEM 22 | | | | | | | | | |

Please provide any comments or suggestions you have about ITEM 22.

**ITEM 23**

Please rate the importance of including this item in a reporting guideline for intervention fidelity in non-drug, non-surgical trials:

**ITEM 23: Describe the degree of flexibility permitted for different intervention aspects, such as in intervention manuals, provider training (e.g., adapting to learning styles and experience), intervention delivery (e.g., adapting to patient type and motivation), receipt (e.g., adapting to learning style and health literacy), and enactment (e.g., adapting to social and environmental context).**

| | Not important | | | | | Very important | | | |
|---|---|---|---|---|---|---|---|---|---|
| | 1 | 2 | 3 | 4 | 5 | 6 | 7 | 8 | 9 |
| ITEM 23 | | | | | | | | | |

Please provide any comments or suggestions you have about ITEM 23.

**ITEM 24**

Please rate the importance of including this item in a reporting guideline for intervention fidelity in non-drug, non-surgical trials:

**ITEM 24: Describe a method to determine whether adaptations made during intervention implementation fall within the allowed flexibility specified in the protocol or compromise fidelity.**

| | Not important | | | | | Very important | | | |
|---|---|---|---|---|---|---|---|---|---|
| | 1 | 2 | 3 | 4 | 5 | 6 | 7 | 8 | 9 |
| ITEM 24 | | | | | | | | | |

Please provide any comments or suggestions you have about ITEM 24.

**ITEM 25**

Please rate the importance of including this item in a reporting guideline for intervention fidelity in non-drug, non-surgical trials:

**ITEM 25: Report how fidelity deviations were dealt with (e.g., additional training was given to providers).**

| | Not important | | | | | Very important | | | |
|---|---|---|---|---|---|---|---|---|---|
| | 1 | 2 | 3 | 4 | 5 | 6 | 7 | 8 | 9 |
| ITEM 25 | | | | | | | | | |

Please provide any comments or suggestions you have about ITEM 25.

**ITEM 26-28**

Please rate the importance of including these items in a reporting guideline for intervention fidelity in non-drug, non-surgical trials:

**ITEM 26: At the trial outset, specify the minimum criteria for selecting trial providers. This can be based on their characteristics (e.g., motivation, flexibility, etc) and qualifications, including education, experience, and credentials.**

| | Not important 1 | 2 | 3 | 4 | 5 | Very important 6 | 7 | 8 | 9 |
|---|---|---|---|---|---|---|---|---|---|
| ITEM 26 | | | | | | | | | |

Please provide any comments or suggestions you have about ITEM 26.

**ITEM 27: When selecting trial providers, describe whether there is a good fit between the potential provider, the population (i.e., matching to key characteristics such as ethnicity) and the intervention (i.e., providers buy into the intervention theory and foundations).**

| | Not important 1 | 2 | 3 | 4 | 5 | Very important 6 | 7 | 8 | 9 |
|---|---|---|---|---|---|---|---|---|---|
| ITEM 27 | | | | | | | | | |

Please provide any comments or suggestions you have about ITEM 27.

**ITEM 28: Report providers characteristics (e.g. years of experience, gender and race/ethnicity) and qualifications (i.e., education, experience, credentials).**

| | Not important | | | | | Very important | | | |
|---|---|---|---|---|---|---|---|---|---|
| | 1 | 2 | 3 | 4 | 5 | 6 | 7 | 8 | 9 |
| ITEM 28 | | | | | | | | | |

Please provide any comments or suggestions you have about ITEM 28.

**ITEM 29-36**

Please rate the importance of including these items in a reporting guideline for intervention fidelity in non-drug, non-surgical trials:

**ITEM 29: Describe a provider training plan that details how providers will be trained, including the methods used to deliver the training (e.g., training manuals, didactic sessions, role modelling, supervision with feedback), and ensure it is driven by the treatment protocol while emphasizing the theoretical underpinnings of the intervention.**

| | Not important | | | | | Very important | | | |
|---|---|---|---|---|---|---|---|---|---|
| | 1 | 2 | 3 | 4 | 5 | 6 | 7 | 8 | 9 |
| ITEM 29 | | | | | | | | | |

Please provide any comments or suggestions you have about ITEM 29.

**ITEM 30: Report whether provider training was delivered as planned, including training content and dose, and the reasons for any discrepancy.**

| | Not important | | | | | Very important | | | |
|---|---|---|---|---|---|---|---|---|---|
| | 1 | 2 | 3 | 4 | 5 | 6 | 7 | 8 | 9 |
| ITEM 30 | | | | | | | | | |

Please provide any comments or suggestions you have about ITEM 30.

**ITEM 31: Report provider training according to a well-defined, a priori competence criterion (e.g., adherence to 80% of the intervention components).**

| | Not important | | | | | Very important | | | |
|---|---|---|---|---|---|---|---|---|---|
| | 1 | 2 | 3 | 4 | 5 | 6 | 7 | 8 | 9 |
| ITEM 31 | | | | | | | | | |

Please provide any comments or suggestions you have about ITEM 31.

**ITEM 32: Report the level of training received by each provider (e.g., frequency of training, attendance to training sessions).**

| | Not important | | | | | Very important | | | |
|---|---|---|---|---|---|---|---|---|---|
| | 1 | 2 | 3 | 4 | 5 | 6 | 7 | 8 | 9 |
| ITEM 32 | | | | | | | | | |

Please provide any comments or suggestions you have about ITEM 32.

**ITEM 33: Describe a method to measure provider skill acquisition and maintenance after training, as well as their understanding of the treatment components, including the underlying rationale and theory.**

| | Not important | | | | | Very important | | |
|---|---|---|---|---|---|---|---|---|
| | 1 | 2 | 3 | 4 | 5 | 6 | 7 | 8 | 9 |
| ITEM 33 | | | | | | | | | |

Please provide any comments or suggestions you have about ITEM 33.

**ITEM 34: Report provider skill acquisition and maintenance after training, including their understanding of the treatment components and the underlying rationale and theory.**

| | Not important | | | | | Very important | | |
|---|---|---|---|---|---|---|---|---|
| | 1 | 2 | 3 | 4 | 5 | 6 | 7 | 8 | 9 |
| ITEM 34 | | | | | | | | | |

Please provide any comments or suggestions you have about ITEM 34.

**ITEM 35: Report if booster training sessions were offered and provided to providers.**

| | Not important | | | | | Very important | | |
|---|---|---|---|---|---|---|---|---|
| | 1 | 2 | 3 | 4 | 5 | 6 | 7 | 8 | 9 |
| ITEM 35 | | | | | | | | | |

Please provide any comments or suggestions you have about ITEM 35.

**ITEM 36: Report the number of providers available to be trained in a specific intervention, and the number of providers required to deliver the specific intervention.**

| | Not important | | | | | Very important | | | |
|---|---|---|---|---|---|---|---|---|---|
| | 1 | 2 | 3 | 4 | 5 | 6 | 7 | 8 | 9 |
| ITEM 36 | | | | | | | | | |

Please provide any comments or suggestions you have about ITEM 36.

**ITEM 37**

Please rate the importance of including this item in a reporting guideline for intervention fidelity in non-drug, non-surgical trials:

**ITEM 37: Specify the provider-participant ratios, caseloads and any other responsibilities that providers had in addition to their participant caseload that may impact the amount of time they can spend on intervention delivery.**

| | Not important | | | | | Very important | | | |
|---|---|---|---|---|---|---|---|---|---|
| | 1 | 2 | 3 | 4 | 5 | 6 | 7 | 8 | 9 |
| ITEM 37 | | | | | | | | | |

Please provide any comments or suggestions you have about ITEM 37.

**ITEM 38-41**

Please rate the importance of including these items in a reporting guideline for intervention fidelity in non-drug, non-surgical trials:

**ITEM 38: Describe a method to assess the degree to which participants received and understood the information/concepts/content provided in the intervention.**

| | Not important | | | | | Very important | | | |
|---|---|---|---|---|---|---|---|---|---|
| | 1 | 2 | 3 | 4 | 5 | 6 | 7 | 8 | 9 |
| ITEM 38 | | | | | | | | | |

Please provide any comments or suggestions you have about ITEM 38.

**ITEM 39: Report the degree to which participants received and understood the information/concepts/content provided in the intervention.**

| | Not important | | | | | Very important | | | |
|---|---|---|---|---|---|---|---|---|---|
| | 1 | 2 | 3 | 4 | 5 | 6 | 7 | 8 | 9 |
| ITEM 39 | | | | | | | | | |

Please provide any comments or suggestions you have about ITEM 39.

**ITEM 40: Describe a method to assess participants' ability to apply the intervention skills to real-life settings (also called 'enactment', 'engagement' or 'participant responsiveness').**

| | Not important | | | | | Very important | | | |
|---|---|---|---|---|---|---|---|---|---|
| | 1 | 2 | 3 | 4 | 5 | 6 | 7 | 8 | 9 |
| ITEM 40 | | | | | | | | | |

Please provide any comments or suggestions you have about ITEM 40.

**ITEM 41: Report the degree to which participants applied the intervention skills into their real life (also called 'enactment', 'engagement' or 'participant responsiveness').**

| | Not important | | | | | Very important | | | |
|---|---|---|---|---|---|---|---|---|---|
| | 1 | 2 | 3 | 4 | 5 | 6 | 7 | 8 | 9 |
| ITEM 41 | | | | | | | | | |

Please provide any comments or suggestions you have about ITEM 41.

**ITEM 42**

Please rate the importance of including this item in a reporting guideline for intervention fidelity in non-drug, non-surgical trials:

**ITEM 42: Report moderators of intervention fidelity.**

| | Not important | | | | | Very important | | | |
|---|---|---|---|---|---|---|---|---|---|
| | 1 | 2 | 3 | 4 | 5 | 6 | 7 | 8 | 9 |
| ITEM 42 | | | | | | | | | |

Please provide any comments or suggestions you have about ITEM 42.

**ITEM 43-44**

Please rate the importance of including these items in a reporting guideline for intervention fidelity in non-drug, non-surgical trials:

**ITEM 43: Describe a method to assess participant acceptability before and during intervention implementation. This includes evaluating the extent to which participants accept the intervention's content and their positive perceptions, beliefs, attitudes, and intentions to use the intervention.**

| | Not important | | | | | Very important | | | |
|---|---|---|---|---|---|---|---|---|---|
| | 1 | 2 | 3 | 4 | 5 | 6 | 7 | 8 | 9 |
| ITEM 43 | | | | | | | | | |

Please provide any comments or suggestions you have about ITEM 43.

**ITEM 44: Report participant acceptability before and during intervention implementation. This includes reporting the extent to which participants accept the intervention's content and their positive perceptions, beliefs, attitudes, and intentions to use the intervention.**

| | Not important | | | | | Very important | | | |
|---|---|---|---|---|---|---|---|---|---|
| | 1 | 2 | 3 | 4 | 5 | 6 | 7 | 8 | 9 |
| ITEM 44 | | | | | | | | | |

Please provide any comments or suggestions you have about ITEM 44.

**ITEM 45**

Please rate the importance of including this item in a reporting guideline for intervention fidelity in non-drug, non-surgical trials:

**ITEM 45: Report participants' characteristics (e.g., age, gender, comorbid conditions, personality traits), preferences, and behaviours that can influence their engagement with the intervention and intervention fidelity by providers.**

| | Not important | | | | | Very important | | | |
|---|---|---|---|---|---|---|---|---|---|
| | 1 | 2 | 3 | 4 | 5 | 6 | 7 | 8 | 9 |
| ITEM 45 | | | | | | | | | |

Please provide any comments or suggestions you have about ITEM 45.

**ITEM 46**

Please rate the importance of including this item in a reporting guideline for intervention fidelity in non-drug, non-surgical trials:

**ITEM 46: Report participants' satisfaction with the intervention and incorporate into the outcome evaluation.**

| | Not important | | | | | Very important | | | |
|---|---|---|---|---|---|---|---|---|---|
| | 1 | 2 | 3 | 4 | 5 | 6 | 7 | 8 | 9 |
| ITEM 46 | | | | | | | | | |

Please provide any comments or suggestions you have about ITEM 46.

**ITEM 47-48**

Please rate the importance of including these items in a reporting guideline for intervention fidelity in non-drug, non-surgical trials:

**ITEM 47: Describe a method to assess participant adherence to the intervention.**

| | Not important | | | | | Very important | | | |
|---|---|---|---|---|---|---|---|---|---|
| | 1 | 2 | 3 | 4 | 5 | 6 | 7 | 8 | 9 |
| ITEM 47 | | | | | | | | | |

Please provide any comments or suggestions you have about ITEM 47.

**ITEM 48: Report participant adherence to the intervention.**

| | Not important | | | | | Very important | | | |
|---|---|---|---|---|---|---|---|---|---|
| | 1 | 2 | 3 | 4 | 5 | 6 | 7 | 8 | 9 |
| ITEM 48 | | | | | | | | | |

Please provide any comments or suggestions you have about ITEM 48.

**ITEM 49**

Please rate the importance of including this item in a reporting guideline for intervention fidelity in non-drug, non-surgical trials:

**ITEM 49: Specify whether the fidelity measures/tools were developed during the study or adapted from existing ones.**

| | Not important | | | | | Very important | | | |
|---|---|---|---|---|---|---|---|---|---|
| | 1 | 2 | 3 | 4 | 5 | 6 | 7 | 8 | 9 |
| ITEM 49 | | | | | | | | | |

Please provide any comments or suggestions you have about ITEM 49.

**ITEM 50**

Please rate the importance of including this item in a reporting guideline for intervention fidelity in non-drug, non-surgical trials:

**ITEM 50: Specify how the fidelity measures/tools align with the research questions, intervention content, and underlying theoretical or concept models.**

| | Not important | | | | | Very important | | | |
|---|---|---|---|---|---|---|---|---|---|
| | 1 | 2 | 3 | 4 | 5 | 6 | 7 | 8 | 9 |
| ITEM 50 | | | | | | | | | |

Please provide any comments or suggestions you have about ITEM 50.

**ITEM 51**

Please rate the importance of including this item in a reporting guideline for intervention fidelity in non-drug, non-surgical trials:

**ITEM 51: Describe how stakeholders were involved in the development of the fidelity measures/tools.**

| | Not important | | | | | Very important | | | |
|---|---|---|---|---|---|---|---|---|---|
| | 1 | 2 | 3 | 4 | 5 | 6 | 7 | 8 | 9 |
| ITEM 51 | | | | | | | | | |

Please provide any comments or suggestions you have about ITEM 51.

**ITEM 52**

Please rate the importance of including this item in a reporting guideline for intervention fidelity in non-drug, non-surgical trials:

**ITEM 52: Describe each item in the fidelity measures/tools and outline the process of their selection or development, specifying who was involved in creating or choosing them.**

| | Not important | | | | | Very important | | | |
|---|---|---|---|---|---|---|---|---|---|
| | 1 | 2 | 3 | 4 | 5 | 6 | 7 | 8 | 9 |
| ITEM 52 | | | | | | | | | |

Please provide any comments or suggestions you have about ITEM 52.

**ITEM 53**

Please rate the importance of including this item in a reporting guideline for intervention fidelity in non-drug, non-surgical trials:

**ITEM 53: Justify the number of response categories in the fidelity measure/tool, for example, why a 3-point scale (delivered, partially delivered, not delivered) was chosen over other scales.**

| | Not important | | | | | Very important | | | |
|---|---|---|---|---|---|---|---|---|---|
| | 1 | 2 | 3 | 4 | 5 | 6 | 7 | 8 | 9 |
| ITEM 53 | | | | | | | | | |

Please provide any comments or suggestions you have about ITEM 53.

**ITEM 54**

Please rate the importance of including this item in a reporting guideline for intervention fidelity in non-drug, non-surgical trials:

**ITEM 54: If different weights were assigned to items in the fidelity measure/tool, report how these weights were determined, including the method used (e.g., theory, empirical evidence, or sensitivity analysis) and the rationale for this approach.**

| | Not important | | | | | Very important | | | |
|---|---|---|---|---|---|---|---|---|---|
| | 1 | 2 | 3 | 4 | 5 | 6 | 7 | 8 | 9 |
| ITEM 54 | | | | | | | | | |

Please provide any comments or suggestions you have about ITEM 54.

**ITEM 55**

Please rate the importance of including this item in a reporting guideline for intervention fidelity in non-drug, non-surgical trials:

**ITEM 55: Describe how the fidelity measures/tools were piloted, including who conducted and the number of assessors involved.**

| | Not important | | | | | Very important | | | |
|---|---|---|---|---|---|---|---|---|---|
| | 1 | 2 | 3 | 4 | 5 | 6 | 7 | 8 | 9 |
| ITEM 55 | | | | | | | | | |

Please provide any comments or suggestions you have about ITEM 55.

**ITEM 56**

Please rate the importance of including this item in a reporting guideline for intervention fidelity in non-drug, non-surgical trials:

**ITEM 56: Specify the psychometric properties of the fidelity measures/tools, including inter-rater reliability, validity, and sensitivity to change.**

| | Not important | | | | | Very important | | | |
|---|---|---|---|---|---|---|---|---|---|
| | 1 | 2 | 3 | 4 | 5 | 6 | 7 | 8 | 9 |
| ITEM 56 | | | | | | | | | |

Please provide any comments or suggestions you have about ITEM 56.

**ITEM 57**

Please rate the importance of including this item in a reporting guideline for intervention fidelity in non-drug, non-surgical trials:

**ITEM 57: Specify the usability of the fidelity measures/tools, including their ease of use and practical application in real-world settings.**

| | Not important | | | | | Very important | | | |
|---|---|---|---|---|---|---|---|---|---|
| | 1 | 2 | 3 | 4 | 5 | 6 | 7 | 8 | 9 |
| ITEM 57 | | | | | | | | | |

Please provide any comments or suggestions you have about ITEM 57.

**ITEM 58**

Please rate the importance of including this item in a reporting guideline for intervention fidelity in non-drug, non-surgical trials:

**ITEM 58: Specify the benchmarks used to interpret the psychometrics of the fidelity measures/tools. For example, report what constitutes an acceptable level of inter-rater reliability.**

| | Not important | | | | | Very important | | | |
|---|---|---|---|---|---|---|---|---|---|
| | 1 | 2 | 3 | 4 | 5 | 6 | 7 | 8 | 9 |
| ITEM 58 | | | | | | | | | |

Please provide any comments or suggestions you have about ITEM 58.

**ITEM 59**

Please rate the importance of including this item in a reporting guideline for intervention fidelity in non-drug, non-surgical trials:

**ITEM 59: Describe how data from multiple fidelity measurement methods, if applicable, are integrated and triangulated to provide comprehensive fidelity results. For example, describe how quantitative data (e.g., from fidelity measures/tools) are combined with qualitative data (e.g., self-reports).**

| | Not important | | | | | | | Very important |
|---|---|---|---|---|---|---|---|---|
| | 1 | 2 | 3 | 4 | 5 | 6 | 7 | 8 | 9 |
| ITEM 59 | | | | | | | | | |

Please provide any comments or suggestions you have about ITEM 59.

**ITEM 60**

Please rate the importance of including this item in a reporting guideline for intervention fidelity in non-drug, non-surgical trials:

**ITEM 60: Describe the strengths and weaknesses of fidelity assessment methods used.**

| | Not important | | | | | | | Very important |
|---|---|---|---|---|---|---|---|---|
| | 1 | 2 | 3 | 4 | 5 | 6 | 7 | 8 | 9 |
| ITEM 60 | | | | | | | | | |

Please provide any comments or suggestions you have about ITEM 60.

**ITEM 61**

Please rate the importance of including this item in a reporting guideline for intervention fidelity in non-drug, non-surgical trials:

**ITEM 61: Specify the specific statistical analysis conducted to obtain fidelity results (e.g., ANOVA, regression).**

| | Not important | | | | | Very important | | | |
|---|---|---|---|---|---|---|---|---|---|
| | 1 | 2 | 3 | 4 | 5 | 6 | 7 | 8 | 9 |
| ITEM 61 | | | | | | | | | |

Please provide any comments or suggestions you have about ITEM 61.

**ITEM 62**

Please rate the importance of including this item in a reporting guideline for intervention fidelity in non-drug, non-surgical trials:

**ITEM 62: Describe the rationale for any sensitivity analysis of fidelity data (e.g., intention-to-treat versus per protocol analysis, removing low fidelity cases).**

| | Not important | | | | | Very important | | | |
|---|---|---|---|---|---|---|---|---|---|
| | 1 | 2 | 3 | 4 | 5 | 6 | 7 | 8 | 9 |
| ITEM 62 | | | | | | | | | |

Please provide any comments or suggestions you have about ITEM 62.

**ITEM 63**

Please rate the importance of including this item in a reporting guideline for intervention fidelity in non-drug, non-surgical trials:

**ITEM 63: Specify the rate of missing fidelity data and how it was managed.**

| | Not important | | | | | Very important | | |
|---|---|---|---|---|---|---|---|---|
| | 1 | 2 | 3 | 4 | 5 | 6 | 7 | 8 | 9 |
| ITEM 63 | | | | | | | | | |

Please provide any comments or suggestions you have about ITEM 63.

**ITEM 64**

Please rate the importance of including this item in a reporting guideline for intervention fidelity in non-drug, non-surgical trials:

**ITEM 64: Specify if a component analysis was conducted (e.g., identification of which intervention components are critical for producing effects).**

| | Not important | | | | | Very important | | |
|---|---|---|---|---|---|---|---|---|
| | 1 | 2 | 3 | 4 | 5 | 6 | 7 | 8 | 9 |
| ITEM 64 | | | | | | | | | |

Please provide any comments or suggestions you have about ITEM 64.

**ITEM 65**

Please rate the importance of including this item in a reporting guideline for intervention fidelity in non-drug, non-surgical trials:

**ITEM 65: Specify how the sample size for fidelity assessment was determined (e.g., all sessions, random subset of sessions). If the fidelity sample was randomly selected, explain the randomisation process.**

| | Not important | | | | | Very important | | | |
|---|---|---|---|---|---|---|---|---|---|
| | 1 | 2 | 3 | 4 | 5 | 6 | 7 | 8 | 9 |
| ITEM 65 | | | | | | | | | |

Please provide any comments or suggestions you have about ITEM 65.

**ITEM 66**

Please rate the importance of including this item in a reporting guideline for intervention fidelity in non-drug, non-surgical trials:

**ITEM 66: Describe any actions taken to minimise selection bias when selecting sessions to be rated and how the fidelity sample is representative of the whole trial sample.**

| | Not important | | | | | Very important | | | |
|---|---|---|---|---|---|---|---|---|---|
| | 1 | 2 | 3 | 4 | 5 | 6 | 7 | 8 | 9 |
| ITEM 66 | | | | | | | | | |

Please provide any comments or suggestions you have about ITEM 66.

**ITEM 67**

Please rate the importance of including this item in a reporting guideline for intervention fidelity in non-drug, non-surgical trials:

**ITEM 67: Specify if the fidelity sample represents different stages of the**

**intervention (e.g., early, middle, or late) and whether fidelity assessments covered full sessions or only specific segments.**

| | Not important | | | | | Very important | | | |
|---|---|---|---|---|---|---|---|---|---|
| | 1 | 2 | 3 | 4 | 5 | 6 | 7 | 8 | 9 |
| ITEM 67 | | | | | | | | | |

Please provide any comments or suggestions you have about ITEM 67.

**ITEM 68-71**

Please rate the importance of including these items in a reporting guideline for intervention fidelity in non-drug, non-surgical trials:

**ITEM 68: Report fidelity results for all study arms, including the control intervention arm.**

| | Not important | | | | | Very important | | | |
|---|---|---|---|---|---|---|---|---|---|
| | 1 | 2 | 3 | 4 | 5 | 6 | 7 | 8 | 9 |
| ITEM 68 | | | | | | | | | |

Please provide any comments or suggestions you have about ITEM 68.

**ITEM 69: Report intervention fidelity in terms of overall fidelity of delivery (i.e., fidelity to all intervention core components across all sessions combined).**

| | Not important | | | | | Very important | | | |
|---|---|---|---|---|---|---|---|---|---|
| | 1 | 2 | 3 | 4 | 5 | 6 | 7 | 8 | 9 |
| ITEM 69 | | | | | | | | | |

Please provide any comments or suggestions you have about ITEM 69.

**ITEM 70: Report intervention fidelity in terms of component fidelity (i.e., fidelity to each intervention core component across all sessions).**

| | Not important | | | | | Very important | | | |
|---|---|---|---|---|---|---|---|---|---|
| | 1 | 2 | 3 | 4 | 5 | 6 | 7 | 8 | 9 |
| ITEM 70 | | | | | | | | | |

Please provide any comments or suggestions you have about ITEM 70.

**ITEM 71: Report intervention fidelity in terms of session fidelity (i.e., fidelity to all intervention core components within each session).**

| | Not important | | | | | Very important | | | |
|---|---|---|---|---|---|---|---|---|---|
| | 1 | 2 | 3 | 4 | 5 | 6 | 7 | 8 | 9 |
| ITEM 71 | | | | | | | | | |

Please provide any comments or suggestions you have about ITEM 71.

**ITEM 72**

Please rate the importance of including this item in a reporting guideline for intervention fidelity in non-drug, non-surgical trials:

**ITEM 72: Specify when fidelity assessments were conducted, such as**

**concurrently with the intervention or after the intervention period was completed.**

| | Not important | | | | | Very important | | | |
|---|---|---|---|---|---|---|---|---|---|
| | 1 | 2 | 3 | 4 | 5 | 6 | 7 | 8 | 9 |
| ITEM 72 | | | | | | | | | |

Please provide any comments or suggestions you have about ITEM 72.

**ITEM 73-77**

Please rate the importance of including these items in a reporting guideline for intervention fidelity in non-drug, non-surgical trials:

**ITEM 73: Specify the number of fidelity assessors and indicate if they performed their evaluations independently.**

| | Not important | | | | | Very important | | | |
|---|---|---|---|---|---|---|---|---|---|
| | 1 | 2 | 3 | 4 | 5 | 6 | 7 | 8 | 9 |
| ITEM 73 | | | | | | | | | |

Please provide any comments or suggestions you have about ITEM 73.

**ITEM 74: Specify the level of blinding of fidelity assessors at the time of fidelity assessments (e.g., blinded to intervention outcomes, provider identity).**

| | Not important | | | | | Very important | | | |
|---|---|---|---|---|---|---|---|---|---|
| | 1 | 2 | 3 | 4 | 5 | 6 | 7 | 8 | 9 |
| ITEM 74 | | | | | | | | | |

Please provide any comments or suggestions you have about ITEM 74.

**ITEM 75: Specify whether fidelity assessors were members of the research team, the intervention delivery team, or external to both.**

| | Not important | | | | | Very important | | | |
|---|---|---|---|---|---|---|---|---|---|
| | 1 | 2 | 3 | 4 | 5 | 6 | 7 | 8 | 9 |
| ITEM 75 | | | | | | | | | |

Please provide any comments or suggestions you have about ITEM 75.

**ITEM 76: Describe how fidelity assessors were trained to use the fidelity measures/tools and conduct the fidelity assessments.**

| | Not important | | | | | Very important | | | |
|---|---|---|---|---|---|---|---|---|---|
| | 1 | 2 | 3 | 4 | 5 | 6 | 7 | 8 | 9 |
| ITEM 76 | | | | | | | | | |

Please provide any comments or suggestions you have about ITEM 76.

**ITEM 77: Specify if there was any requirement for the selection of fidelity assessors (e.g., be a health registered professional).**

| | Not important | | | | | Very important | | | |
|---|---|---|---|---|---|---|---|---|---|
| | 1 | 2 | 3 | 4 | 5 | 6 | 7 | 8 | 9 |
| ITEM 77 | | | | | | | | | |

Please provide any comments or suggestions you have about ITEM 77.

**ITEM 78**

Please rate the importance of including this item in a reporting guideline for intervention fidelity in non-drug, non-surgical trials:

**ITEM 78: Specify an a priori threshold for acceptable level of fidelity or cut-offs.**

| | Not important | | | | | Very important | | | |
|---|---|---|---|---|---|---|---|---|---|
| | 1 | 2 | 3 | 4 | 5 | 6 | 7 | 8 | 9 |
| ITEM 78 | | | | | | | | | |

Please provide any comments or suggestions you have about ITEM 78.

**ITEM 79**

Please rate the importance of including this item in a reporting guideline for intervention fidelity in non-drug, non-surgical trials:

**ITEM 79: Report any enhancement strategies employed to address low fidelity, if applicable.**

| | Not important | | | | | Very important | | | |
|---|---|---|---|---|---|---|---|---|---|
| | 1 | 2 | 3 | 4 | 5 | 6 | 7 | 8 | 9 |
| ITEM 79 | | | | | | | | | |

Please provide any comments or suggestions you have about ITEM 79.

#### SECTION 3 - Add Input

##### SECTION 3 – Additional input

Are there any other aspects of intervention fidelity not mentioned in this survey that should be reported in trials of non-drug, non-surgical interventions? (Optional)

What aspects or resources would help you in adopting an intervention fidelity reporting guideline? (Optional)

Powered by Qualtrics
